# The effects of trauma on neurobiological and psychopathological phenotypes in a transdiagnostic community sample

**DOI:** 10.64898/2025.12.09.25341925

**Authors:** Toby Constable, Jeggan Tiego, Kane Pavlovich, Nancy Ong Tran, Bree Hartshorn, Jessica Kwee, Kate Fortune, Kate Thompson, Sam Brown, Rebecca O’Neill, James McLauchlan, Maggie Eyers, Roman Kotov, Mark A. Bellgrove, Alex Fornito

## Abstract

Trauma is an established risk factor for a diverse range of psychiatric disorders. This effect on risk is widely thought to be mediated, at least partially, by the deleterious impact of trauma on brain structure and function. We tested this neurobiological mediation hypothesis in 670 adults (40% male; age 18–45 years, *M* = 29.60, *SD* = 7.67) with diverse psychiatric histories who undertook self-reported and interview-based assessment of psychopathology along with magnetic resonance imaging (MRI). We evaluated 252 path models, testing direct and indirect (i.e., mediation) effects of distinct types of trauma (sexual or interpersonal physical violence) on measures of brain structure (regional grey matter volume, GMV) and function (estimates of inter-regional functional coupling; FC) as well as dimensional and categorical psychological outcomes (empirically derived latent dimensions of trait psychopathology and lifetime diagnosis of post-traumatic stress disorder). Greater exposure to interpersonal physical violence, but not Sexual Trauma, predicted widespread differences in GMV, including lower fronto-parietal-occipital volume and higher lateral temporal and medial cortical volumes; elevated FC between default, control, visual, and attentional systems; and anticorrelated FC between default, limbic, and somatomotor networks. These patterns were generally more pronounced for individuals with an older age of first exposure and were moderated by survivor sex. We found no robust statistical evidence to support the neurobiological mediation hypothesis. Our findings indicate that trauma exerts independent effects on both neurobiology and psychopathology in ways that depend on trauma type, survivor sex, age of onset, and trauma dose, challenging the presumptive role of these neurophenotypes in trauma-related psychopathology.

## 1. Introduction

Trauma exposure poses a significant risk for the development of adverse psychological outcomes, including multiple psychiatric disorders (Carr et al., 2013; Dunn et al., 2017). Trauma also produces measurable changes in brain structure and function, which are thought to result from the neurotoxic effects of glucocorticoids released during the stress response cycle (Sapolsky et al., 1990) and an extensive remodelling of stress-response related neural architecture (Borrow et al., 2019). Such brain changes can be identified non-invasively in humans as altered regional matter volume (McCrory et al., 2011), regional brain activity (Cassiers et al., 2018), and/or inter-regional functional coupling (FC) (Breukelaar et al., 2021).

Numerous magnetic resonance imaging (MRI) studies have demonstrated that trauma is cross-sectionally associated with heterogeneous effects across the brain, with some regions showing correlated increases or decreases in activity while others exhibit volumetric expansion or contraction (Cassiers et al., 2018). Trauma exposure may exert particularly strong effects on frontal and limbic systems, which are critical for stress regulation and coping (Herzog & Schmahl, 2018; Holz et al., 2023; McCrory et al., 2011; McEwen & Gianaros, 2010). These effects are thought to heighten vulnerability to future stressors (Cassiers et al., 2018; Tomoda, Suzuki, et al., 2009). Volume increases, particularly within primary sensory cortices, may signal potentiated sensory information processing, which elevates environmental threat detection (Cassiers et al., 2018).

Despite such findings, the field has struggled to converge on a replicable neural signature of trauma exposure (Cassiers et al., 2018). Reasons for this limited progress include a reliance on small samples (typically <100), which inflates standard errors and the risk of both Type I and Type II errors (Button et al., 2013; Cao et al., 2025; Marek et al., 2022); the frequent exclusion of individuals with multiple trauma exposures in attempts to isolate specific effects (Cassiers et al., 2018), which may lead to unrepresentative samples, given evidence that multiple exposures represent the norm (Finkelhor et al., 2007; Turner et al., 2010); and the use of inadequate statistical models incapable of parsing known sources of heterogeneity, such as survivor sex (Cassiers et al., 2018). Moreover, while many studies infer that observed trauma-related neurobiological changes are clinically significant and predispose trauma survivors to adverse psychological outcomes, this neurobiological mediation hypothesis is seldom tested directly (Cassiers et al., 2018; Stevens et al., 2018). The few studies that have examined neurobiological mediation have found that the effects of aggregate measures of childhood trauma exposure on psychological resilience, depression relapse risk, and psychological well-being are partially mediated by variations in visual, prefrontal, and insular cortex volumes, and amygdala-related FC (Chen et al., 2024; Harb et al., 2024; Opel et al., 2019; Wan et al., 2022).

Although encouraging, such work has generally relied on coarse aggregations of diverse forms of adversity into single composite scores, precluding investigation of how specific forms of trauma may influence risk for distinct outcomes (Constable, Tiego, Pavlovich, Tran, et al., 2025). In addition, the existing literature has commonly used traditional diagnoses for study inclusion/exclusion or for defining psychiatric outcomes, which reduces sensitivity and statistical power through the application of arbitrary diagnostic thresholds to intrinsically dimensional psychopathological outcomes (Helmchen & Linden, 2000). Moreover, such approaches fail to account for the high levels of within-disorder heterogeneity (e.g., 636,120 symptom combinations yield a diagnosis of PTSD) (Galatzer-Levy & Bryant, 2013) and between-disorder comorbidity (e.g., 90% of people with PTSD meet criteria for another lifetime diagnosis) that are commonly observed in epidemiological samples (Kessler et al., 1995). Removal of participants based on standard hierarchical exclusion rules (i.e., presence of specific comorbid diagnoses) further limits sample representativeness (Kotov et al., 2018). Details of the trauma (e.g. age of onset and survivor sex) are also typically ignored, despite evidence that they can have a major impact on individual outcomes (Guidi et al., 2021; Keyes et al., 2012; Stevens et al., 2018).

These limitations can be addressed by adopting a hierarchical, dimensional, and transdiagnostic approach to measuring psychopathology (Kotov et al., 2017, 2021). This approach involves measuring symptoms across the full spectrum of the dimensional continuum in transdiagnostic samples, thereby studying psychopathological outcomes “as they are”, without imposition of diagnostic decision rules and inclusion/exclusion criteria. Using this approach, we recently observed that exposure to sexual and Interpersonal Physical Violence elevates scores on two of five, broad-band dimensions of typical and maladaptive personality traits: an *Antisocial Schizotypy* dimension encompassing externalizing behaviours (e.g., norm-violation) and schizotypal phenomena (e.g., unusual experiences) and a *Negative Affectivity* dimension encompassing internalizing symptoms, such as depression and anxiety (Constable, Tiego, Pavlovich, Tran, et al., 2025). Interpersonal Physical Violence was strongly linked to elevated Negative Affectivity in females and Antisocial Schizotypy in males, with later age of exposure and higher dosage leading to more severe trait expression. In contrast, Sexual Trauma was a significant risk factor for both Negative Affectivity and Antisocial Schizotypy regardless of age of onset, with no additional risk from multiple exposures. These findings underscore the complex interplay between psychological outcomes and trauma type, frequency, and survivor characteristics such as sex and age of exposure.

Here, we extended these findings to comprehensively test whether these complex links between trauma and psychopathology are at least partially mediated by the impact of trauma on the brain, as quantified using MRI-derived measures of regional grey matter volume (GMV) and inter-regional FC. We further examined whether direct and/or mediated effects of trauma on psychopathology were stronger when considering categorical (i.e., a lifetime diagnosis of PTSD) or dimensional outcomes, while accounting for the moderating effects of trauma type, age of onset, trauma frequency, and survivor sex. Across 252 distinct path models, we found no support for the neurobiological mediation hypothesis. Instead, our findings indicate that trauma exerts distinct and independent effects on both the brain and psychopathology.

## 2 Methods and Analysis

### 2.1 Participants

The initial sample comprised 1,110 participants (30.63% male; age range: 18–45 years, *M* = 29.58, *SD* = 7.75) recruited from the general community via the private company Trialfacts, who organised recruitment via online campaigns and social media platforms (https://trialfacts.com/; see Constable et al. 2025 and Supplementary Material 1.1 for details). All participants were right-handed, with no self-reported history of significant head injury, and provided written informed consent prior to participation. Full details of inclusion and exclusion criteria are provided in Supplementary Material 1.1. This project was approved by the Monash University Human Research Ethics Committee.

### 2.2 Psychopathological Measures

Participants completed three questionnaires as part of an online battery before attending an in-person session that involved diagnostic interviewing, MRI, and cognitive assessments. These questionnaires were chosen to measure dimensional variations in core domains of pathological and normative traits. We focus here on enduring traits rather than time-limited symptoms given the well-documented persistence of trauma-implicated psychopathology across the lifespan (Bonanno et al., 2011).

#### 2.2.1 The Computerized Assessment Test of Personality Disorder Static Form (CAT-PD SF)

The CAT-PD SF assesses maladaptive traits across 33 unidimensional domains (Simms et al., 2011). Each item is rated on a 5-point Likert scale (1, *very untrue of me* – 5, *very true of me*). Previous literature has determined the CAT-PD SF to be a robust, reliable, and valid instrument for measuring personality disorder severity (Long et al., 2021; Ringwald et al., 2023). Reliability was adequate in the present sample, with Cronbach’s alpha ranging from .65 to .95 for each domain (see Supplementary Material 1.2).

#### 2.2.2 The 60-item Next Big 5 -II (BFI-II)

The 60-item Next Big 5 -II (BFI-II) assessed the Big Five personality traits (Neuroticism, Extraversion, Conscientiousness, Agreeableness, and Openness) and their associated sub-facets. Each item was rated on 5-point Likert scales (1, *disagree strongly* – 5, *agree strongly*). Consistent with previous literature (Soto & John, 2017), reliability was adequate in the present sample at the domain but not sub-facet level, with Cronbach’s alpha ranging from .79 to .93, and .56 to .89, respectively (See Supplementary Material 1.2). We include this measure of normative personality traits to capture the full-spectrum of personality traits, with such measures complementing information captured by the CAT-PD-SF (Constable, Tiego, Pavlovich, Tran, et al., 2025).

#### 2.2.3 Principal Components Analysis of the CAT-PD SF and BFI-II

In our prior analyses of 842 participants with complete and valid behavioural data, we used principal components analysis (PCA) with varimax rotation to extract five granular, orthogonal constructs from BFI-II and CAT-PD data that effectively captured variance in typical and maladaptive personality traits (Constable, Tiego, Pavlovich, Tran, et al., 2025). Together, these five components explained 61% of the total variance and corresponded to latent dimensions indexing traits related to Negative Affectivity, Antagonism, Disinhibition, Antisocial Schizotypy, and Detachment (for PC loadings, see Supplementary Material 1.3). Importantly, these dimensions are dipolar, possessing an adaptive and maladaptive end (e.g., Antagonism to Agreeableness; Disinhibition to Conscientiousness) and largely parallel those described in previous literature (Kotov et al., 2021). For the present analyses, the same principal component (PC) scores for Negative Affectivity and Antisocial Schizotypy were investigated given their previously established associations with trauma exposure (Constable, Tiego, Pavlovich, Tran, et al., 2025).

#### 2.2.4 The Trauma History Questionnaire (THQ)

The 24-item THQ screened for various experiences argued to constitute traumatic event exposure (Hooper et al., 2011). Questions regarding exposure to radioactive disasters were dropped due to low endorsement rates. Participants responded *’yes’* or *’no’* to experiencing each trauma. If they responded ‘*yes*,’ they also indicated the number of times the trauma occurred and the earliest three ages of exposure. We organised items to form the Sexual Trauma and Interpersonal Physical Violence Trauma subscales for our main analyses as in Constable et al., 2025 given their particularly impactful nature; general non-interpersonal trauma and traumatic grief loads were also computed from this data and used as model covariates (see Supplementary Material 1.4).

#### 2.2.5 Socioeconomic Status

We additionally indexed family background (i.e., parental) and participant socioeconomic status (SES) using composite scores derived by averaging income estimates with acquired education level after both were pre-centred and scaled using respective medians and interquartile ranges. Income was estimated by cross-referencing employment industry and education level against Australian Bureau of Statistics (ABS) data (*ANZSIC,* 2013; *ASCED,* 2001). Education levels were rank-ordered based on the Australian Qualifications Framework (2011). By considering a range of information, the composites were deemed robust indicators of SES (Galobardes et al., 2007; Lindberg et al., 2022). See Supplementary Material 1.5 for validation results.

### 2.3. Diagnostic interview

Participants completed the Structured Clinical Interview for the Diagnostic and Statistical Manual 5^th^ Edition Research Version (SCID-5-RV) (First & Williams, 2017), a semi-structured diagnostic interview administered by trained research assistants. For each disorder, participants were coded with a 0 (no diagnosis), or 2 (meeting diagnostic threshold) for both current and lifetime presentations (those coded 1; subthreshold, were recoded as 0). Hierarchical exclusion rules were not applied due to concerns that this practice rests on problematic assumptions (e.g., that categorical disorders reflect distinct and mutually exclusive aetiologies) and evidence that it can distort associations between disorders and correlates of interest (Kotov et al., 2018). We focused on lifetime PTSD rates in the present study (which we coded as 0 – no PTSD; 1 –full criteria PTSD), as an alternative outcome to our dimensional measures, given that PTSD is one of the most commonly examined categorical disorders in the psychotraumatology literature.

### 2.4 Behavioural Data Cleaning and Preparation

We retained data only from participants who completed at least 85% of the testing battery. As the removal of inattentive participants improves statistical power (Maniaci & Rogge, 2014), we embedded 20 attention check items within the main battery (e.g., “Please select ’Moderately True of Me”). Individuals completing at least 85% of these items were screened for outliers, such that individuals with scores exceeding 1.5 inter-quartile ranges above quartile three were removed from the analyses. Our final sample comprised 842 people with adequate survey data (Male = 37.89%; age 18 – 45 years, *M* = 29.73, *SD* = 7.77; see Supplementary Material 1.1 for demographic details). Within this sample, we used binomial general linear models to predict patterns of missingness in the survey data, revealing that any residual unavailable data were Missing at Random (MAR) (Austin et al., 2021). The missing data were then imputed according to established procedures (Honaker et al., 2011) (see Supplementary Material 1.7 for further details).

### 2.5 MRI Data Acquisition and Data Cleaning

T1-weighted and functional MRI data were collected on a Siemens 3T Skyra (Siemens Healthineers, Erlangen, Germany). Details on imaging parameters, preprocessing pipelines, and head-motion can be found in Supplementary Material 1.6. Briefly, T1-weighted Magnetization-prepared 2 rapid-acquisition gradient-echoes (MP2RAGE) scans were first processed using the software package *presurfer* (Kashyap et al., 2021) (https://github.com/srikash/presurfer) before passing through Freesurfer’s (7.4.0) recon-all pipeline (Dale et al., 1999) to reconstruct cortical white and pial surfaces. We then parcellated the cortex into 300 discrete regions-of-interest using the well-validated Schaefer300 atlas (Schaefer et al., 2018), projected to individual surfaces using freely available code (https://github.com/ThomasYeoLab/CBIG/tree/master/stable_projects/brain_parcellation/Schaefer2018_LocalGlobal/Parcellations/project_to_individual). To extract subcortical volumes, the data were projected to MNI152 space using a linear transform and winsorization via antsRegistration (Tustison et al., 2021) before the Melbourne Scale 2 Subcortical atlas was used to divide the subcortex into 32 subcortical functionally homogenous regions (Tian et al., 2020). These data were concatenated to form a complete description of cortical-subcortical GMV data. Of the 1,110 participants recruited, 743 completed T1-weighted scan data collection. Ten individuals failed Freesurfer reconstructions. As such, adequate quality T1-weighted scans, determined by visual quality control, were available for a total of 733 participants.

For fMRI, multiple echoes acquired at each time point were first optimally combined using an echo-dependent weighted average (DuPre et al., 2021; Kundu et al., 2013, 2017). Motion and other noise sources were removed using a combination of Independent Components Analysis based Automatic Removal of Motion Artefacts (ICA-AROMA) (Pruim et al., 2015) and RIPTiDe (Frederick et al., 2012), in addition to 24-head motion parameter regression (Satterthwaite et al., 2013). This pipeline has shown strong performance on various benchmarks of denoising efficacy for multi-echo, multi-band fMRI data (Constable, Tiego, Pavlovich, Sangchooli, et al., 2025). The resulting images were spatially normalized to the MNI152 template and parcellated using a combined Schaefer300 and Melbourne Scale 2 Subcortical Atlas (Schaefer et al., 2018; Tian et al., 2020) before estimating inter-regional FC as the product-moment correlation between every pair of regional time series, as implemented in Nilearn based on Nipy (M. Brett et al., 2020; Gorgolewski et al., 2011). See https://github.com/cicadawing/Single-vs-Multi-Echo-fMRI-Denoising-Strategies for freely available code. Of the 1,110 participants recruited, 694 completed rs-fMRI data collection. A total of 105 participants were excluded based on preprocessing failures and/or excess head-motion, leaving 589 participants available for fMRI analysis.

### 2.6 Analysis

Complex high-dimensional mediation models are increasingly common in the literature (Chén et al., 2018; Pearl, 2014; Robins et al., 2010), but they have not yet been developed for the path models we consider, which also account for moderating factors and correlation of residuals (see 3.3). We therefore favoured a ‘serial mediation analysis’ approach in which we tested one mediator at a time. An implicit assumption of this strategy is that mediators and outcome variables between models are not confounded by one another (‘sequential ignorability’ – no unmeasured confounding). Violations of this assumption can inflate error and pose difficulties in isolating exact mediating mechanisms (Chén et al., 2018; Pearl, 2014; Robins et al., 2010). We note that much of the previous mediation literature linking trauma to psychopathology via neurobiology has been vulnerable to such violations, with analyses often targeting specific brain regions (sometimes a priori, sometimes in a serial exhaustive exploratory search) without model correction for variation in other correlated neurophenotypic measures. We therefore ensured that our mediating variables in each model were orthogonal using PCA of the imaging measures. We ran separate PCAs for the GMV and FC measures. All analyses were conducted in R (R Core Team, 2013).

#### 2.6.1 Data-Driven Dimension Reduction of MRI Data

##### GMV Data-Driven Dimension Reduction

For the GMV analysis, the input data matrix comprised 733 rows (participants with adequate T1 data and Freesurfer reconstructions) and 332 columns (number of brain regions). We used permutation testing to identify PCs accounting for statistically significant levels of variance, along with bootstrapping to assess the stability of the PCs over bootstrap samples. In this context, stability was defined as the similarity or congruence of component loadings across iterations; i.e., vectors describing component loadings for the same PC were compared across bootstrap samples using Spearman’s correlation (*r)* and Tucker Congruences, where ≥.95 for both statistics indicated stability (Lorenzo-Seva & ten Berge, 2006). To assess the statistical significance of regional loadings on each significant PC, we also permuted each column in the data matrix 10,000 times while keeping all other column vectors fixed. Originally observed loadings and those derived from the null distribution were then compared to determine the significance of each regional loading value (Linting et al., 2011). We retained only those PCs that were statistically significant (i.e., *p* <.05 uncorrected), stable under bootstrapping, and adequately identified (i.e., >2 significant loadings) (Kline, 2023).

##### FC Data-Driven Dimension Reduction

For the FC analysis, we first reduced the 332 X 332 FC matrix of each individual to a 20 X 20 matrix representing the average FC between every pair of 17 canonical functional brain networks for the cortex (Schaefer et al., 2018; Yeo et al., 2011) and the three functional brain networks identified at the subcortical level by the Melbourne Subcortical Atlas (Tian et al., 2020). This data reduction step was performed by simply taking the mean of the FC estimates within and between networks and served to reduce the number of neurobiological features, given the complexity of our models. The vectorised upper triangle of the 20 X 20 average network FC matrix (including the diagonal) was used as input for PCA. The input data matrix thus comprised 589 rows (participants with quality FC matrices) and 210 columns (number of FC estimates). PCs were retained if they were deemed significant and stable by permutation and bootstrap testing, as in the GMV analysis. To ensure that our FC findings were not due to our network-based coarse-graining approach, we also fitted the same mediation models to PCs extracted from a full 132-region FC matrix (i.e., without network averaging; see Supplementary Material 1.8 for a full description and results from the data reduction strategy, and Supplementary 2 for mediation parameter estimates for the Schaefer100-based sensitivity analyses).

#### 2.6.2 Path Models

We ran multiple path models with robust maximum likelihood estimation using *lavaan* (Rosseel, 2012), *lavaan.mi* (Jorgensen et al., 2022), and *semTools* (Jorgensen et al., 2022) for R. Our models varied in terms of the trauma predictor (i.e., either Interpersonal Physical Violence or Sexual Trauma), the neurobiological mediator (i.e., each GMV or FC PC was tested one-at-a-time), and the outcome (i.e., Antisocial Schizotypy, Negative Affectivity, or lifetime PTSD diagnosis). We therefore ran a total of *P × M × O* models, where *P* represents the number of predictors, *M* the number of mediators, and *O* the number of outcomes. Moderators in the model included sex and age of onset for Interpersonal Physical Violence models, and only sex for Sexual Trauma models because of the inclusion of non-exposed individuals. This approach was informed by our prior work where we established the relative importance of these contextual factors for these specific trauma types (Constable, Tiego, Pavlovich, Tran, et al., 2025).

A schematic of the models we consider is presented in Figure 1. Path *A* refers to the direct effect of trauma exposure on neurobiology (i.e., from the predictor to the mediator); Path *B* the direct effect of neurobiology on psychopathology (i.e., from the mediator to the outcome); Path *C’* the direct effect of trauma on psychopathology, accounting for the mediator; and Path *C* the total effect of trauma on psychopathology, prior to adjusting for the mediator (Kline, 2023) (note that last path has been established by prior work, and thus forms the starting point for the present analysis; see Constable et al., 2025). Indirect effects (mediation) are computed as the product of Paths *A* and *B*. The neurobiological mediation model is thus supported if this indirect path is statistically significant. The independent effects model is supported by the absence of a mediation effect, coupled with significant effects on Paths *A* and *C*.

**Figure 1.**
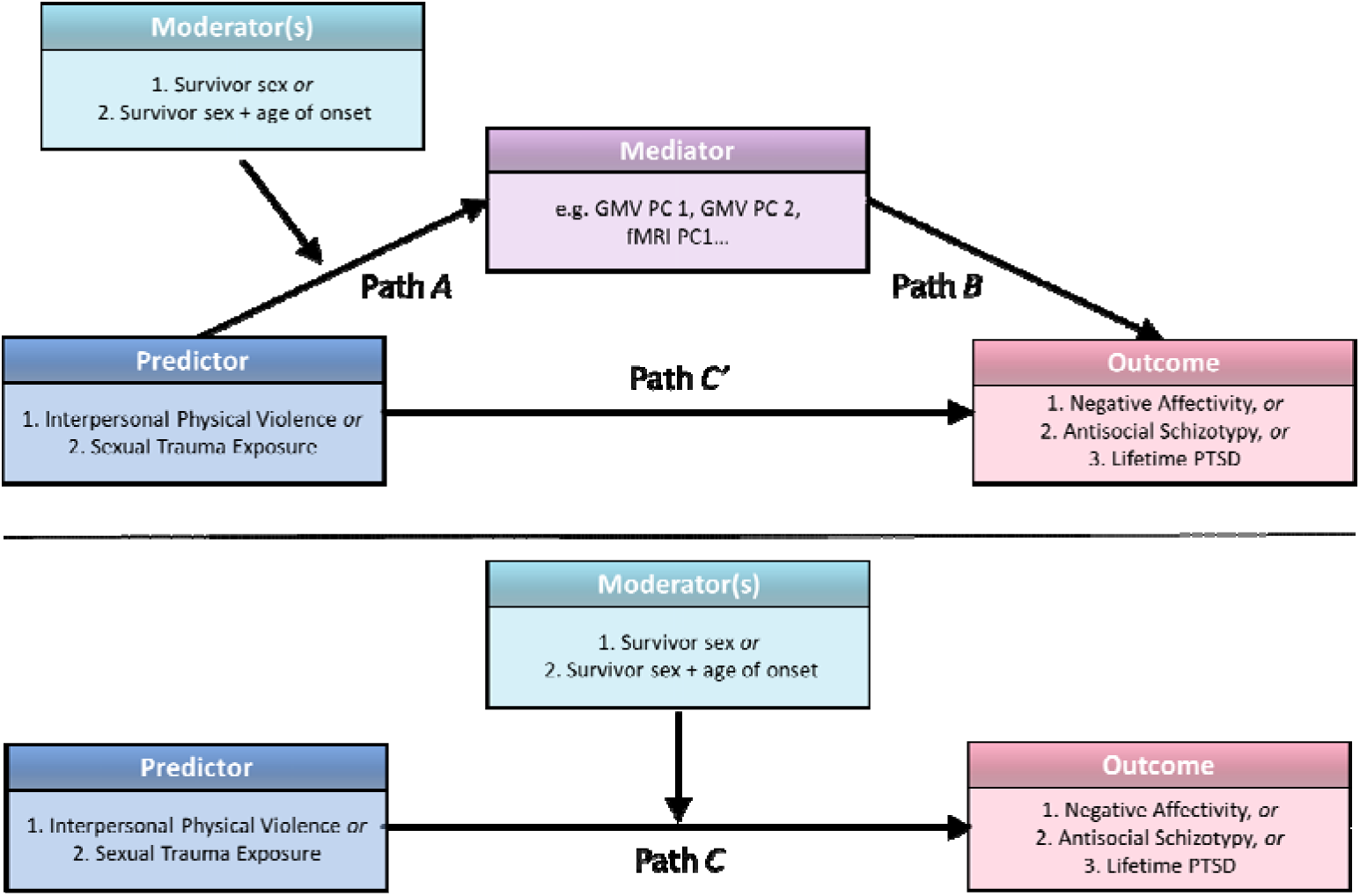
Schematic of the mediation models considered. Arrows indicate hypothesised directional effects. For simplicity, moderators for moderated indirect and moderated direct effects are only indicated along Path *A* in the top panel, but their effects were considered for all paths. The horizontal dotted line separates two conceptualisations of the mediation model: in the upper half, the effect of the neurobiological variable is partialled out along Path C′; in the lower half, Path C′ reflects the total effect of the predictor on the outcome without mediation variable input.

Monte Carlo simulations with 20,000 repeats were used to generate unstandardised robust confidence intervals for all moderated and unmoderated indirect effects, using a range of liberal (2.5%) to conservative (0.0001%) alpha levels (Preacher & Selig, 2012). Confidence intervals are generally considered superior to single-point significance tests in mediation analysis as they provide more accurate estimates of indirect effects and reduce the risk of both Type I and II errors for the sampling distribution given that the product of Paths *A* and *B* tend to be heavily skewed (Preacher & Hayes, 2008; Preacher & Selig, 2012). Given the volume of tests conducted (252 models, each with direct, indirect and moderated indirect effects estimated), mediation effects were first flagged if their confidence intervals excluded zero at relatively liberal thresholds. Confidence in these effects was considered stronger if they also remained significant under more conservative confidence interval ranges. Monte Carlo simulations were performed using the *semTools* package (Jorgensen et al., 2022).

For interpersonal physical violence, we used random slopes and random intercepts linear models to test dose-response relationships in exposed individuals alone, given that the most robust relationships for this trauma type are found in escalating amounts of exposure beyond a single instance rather than between exposed an non-exposed individuals (Constable, Tiego, Pavlovich, Tran, et al., 2025). Following our prior work, for both direct and indirect effects, we also evaluated interactions between survivor sex and trauma dose, and between age of onset and dose.

For Sexual Trauma, we previously found that one exposure is sufficient to elevate trait psychopathology, with no additional effects due to added exposures (Constable, Tiego, Pavlovich, Tran, et al., 2025). We therefore implemented random slopes and random intercepts models while binarising Sexual Trauma frequency into 0 (*Never Exposed*) and 1 (*Exposed*), whilst similarly investigating the moderating effect of sex for direct and indirect effects. Thus, unlike Interpersonal Physical Violence models, Sexual Traumas leveraged the full sample.

As per our prior work (Constable, Tiego, Pavlovich, Tran, et al., 2025), simple slopes for age of onset were examined for Interpersonal Physical Violence for ages 6, 10, 12, 16, and 25. Age 6 represents the earliest stage at which autobiographical memory is thought to stabilise (Nelson & Fivush, 2004) and is a sensitive developmental age for the development of elevated Antisocial Schizotypy following trauma exposure (Constable, Tiego, Pavlovich, Tran, et al., 2025). Ages 10, 12, and 16 are considered critical developmental periods for the onset of trauma (Stevens et al., 2018), particularly Negative Affectivity (Constable, Tiego, Pavlovich, Tran, et al., 2025). Age 25 was chosen to represent adulthood. We note that the retrospective nature of the study means that the recalled ages of onset may not to be precise. We therefore discuss results with regards to ages of onset according to broad developmental stages – i.e., early childhood, preadolescence, adolescence, and adulthood.

Covariates for all models included age, family SES, sex, general/non-interpersonal trauma dose, and sudden loss/grief trauma dose. To control for brain size, intracranial volume was included as a covariate when testing grey matter volume mediators beyond PC1 (Wang et al., 2024), which largely reflected global volume. To reduce multicollinearity (Kraemer & Blasey, 2006), all variables were grand-mean-centred (except for Interpersonal Physical Violence dose, which was centred at 1 to facilitate investigations into effects of single exposures at distinct ages of onset; i.e., simple intercept analyses; and Lifetime PTSD, which was coded 0, 1). Two-way interactions were computed from pre-centred predictors. In the Sexual Trauma model, Interpersonal Physical Violence trauma dose was added as a covariate (and vice versa) to ensure the specificity of trauma effects. To account for the widely reported interrelationships between trauma types (Finkelhor et al., 2007; Turner et al., 2010), residuals between trauma covariates were permitted to covary. This specification also enabled assessment of model fit: mediation models with only manifest variables are typically just-identified (*df* = 0), precluding estimation of model global fit indices. By allowing trauma variables to covary, the model gains degrees of freedom and allows an assessment of overall model fit.

Chi-square difference (Δχ²) testing was conducted for all models, comparing the observed and expected (model-implied) covariance matrices, with *p* > .05 indicating acceptable model fit. We also report the Root Mean Square Error of Approximation (RMSEA; ranging from 0 to 1), which assesses the difference between the observed and model-implied covariance matrices, such that lower values indicate better fit (Kline, 2023).

All non-covariate direct (moderated and unmoderated) effects were aggregated from all models before undergoing Benjamini-Hochberg False Discovery Rate (B-H FDR) correction to an alpha level of .05 (Benjamini & Hochberg, 1995). We investigated any interaction terms between trauma exposure and neurobiology significant at p< .05, uncorrected (i.e., simple slopes analyses, investigating the effect of trauma on neurobiology at specific values of a moderator). We relied on this lower threshold for evaluating interactions because omnibus statistical non-significance does not necessarily preclude moderating effects for interaction terms (Brambor et al., 2006, Esarey & Sumner, 2018). We nonetheless included *p*-values for omnibus interaction terms and simple slopes in the B-H FDR procedure to limit Type-1 error. The combined use of BH–FDR correction, where applicable (such as for the unmoderated and moderated direct effects of interest where *p* values were available) with a range of confidence intervals estimated otherwise (i.e., for indirect effects, where *p* values were unreliable and CI generation was required), was employed to identify robust findings (Y. Li et al., 2023; Pearl, 2014).

#### 2.6.3 Path Robustness

Because mediation models estimate Paths *A, B,* and *C*′ simultaneously and each path represents a partial effect, modifying one variable can influence the estimation of all paths. This interdependence is especially impactful when effects are small, unstable, or near the threshold of significance. Parameters that remain statistically and conceptually consistent (i.e., retaining the same sign) across reasonable analytic variations are often taken as evidence of robustness (Leamer, 1983; Simonsohn et al., 2020; Steegen et al., 2016). Accordingly, direct effects (Paths *A* and *B*) were interpreted only if they maintained statistical significance, consistent direction, and a comparable beta weight when other model components were altered in ways that were not expected to influence the path of interest. For example, if an association between the exposure and a mediator (Path *A*) was observed when the outcome variable was Negative Affectivity but disappeared when it was Lifetime PTSD, the effect was deemed non-robust, since a robust direct effect of trauma on the brain should be independent of the overall psychological outcome considered in the model. Similarly, a mediator–outcome association (Path *B*) present in the Interpersonal Physical Violence model but absent in the Sexual Trauma model (i.e., when the exposure variable was altered) was also considered non-robust. This approach ensured that interpreted direct effects were generalisable and not merely conditional on model specification.

As a final post hoc sensitivity check, we repeated analyses restricted to participants exposed to Sexual Trauma only to ensure main findings for Paths *A* and *B* were not an artefact of binarising exposure to Sexual Trauma. We used the same modelling strategy as for Interpersonal Physical Violence (see 3.3), testing significant Paths *A* and *B* and indirect effects while examining moderation by sex and age of onset in a sample comprised only of those exposed to that trauma type. Given the *post hoc* nature of this analysis, we applied FDR correction to parameters of interest in a separate family of tests, consistent with recommendations for liberal alpha thresholds in subgroup analyses (Thabane et al., 2013). We investigated only models with adequate overall fit (59 of 126 models tested). See Supplementary Material 3 for a report of the significant (pre-corrected) effects of interest.

#### 2.6.4 Aggregating results across PCs

We tested models for over 20 PCs for each imaging modality (see Results). To generate a simplified representation of the findings aggregated across PCs, we tallied the frequency with which each region or FC estimate appeared in an implicated PC. The direction of effect for each region/estimate was calculated as the product of (i) the sign of the trauma–PC association; and (ii) the sign of the region/FC loading onto that PC. As such, positive scores indicate that volume or FC increased in association with higher levels of exposure, while negative scores indicate that higher exposure was associated with volume reductions or stronger FC anticorrelations. Tallies were summed across respective regions and FC pairs to provide a coarse indication of effect direction and consistency across implicated PCs. Consequently, regions with summed tallies near zero either showed no convergent directionality (sum = 0), or did not significantly load onto any implicated PC.

## 4 Results

### 4.1 Dimension reduction of neurobiological variables

Permutation testing indicated that the first 22 PCs of GMV accounted for statistically significant portions of variance. Of these, only the first 20 were found to have at least 3 significant loadings (i.e., only the first 20 were adequately identified). We therefore retained these first 20 PCs, which had stable loadings across bootstrap resamples. These PCs explained 60.17% of the variance in the original data. The statistically thresholded spatial patterns of each PC are shown in Supplementary Material 4.

The PCA of the network-summarised FC measures identified 25 PCs accounting for significant portions of variance, but only the first 22 had at least 3 significant loadings. These 22 PCs had stable loadings across bootstrap samples and explained 67.95% of the variance of the original data. The statistically thresholded network patterns corresponding to each PC are shown in Supplementary Material 5.

### 4.2 Overall Model Sample Characteristics and Fit Statistics

Our mediation models tested one trauma type (i.e., sexual, or interpersonal physical violence), one mediator (i.e., one of the neurobiological PCs) and one psychopathological outcome variable (i.e., Antisocial Schizotypy, Negative Affectivity, or lifetime PTSD) at a time, resulting in 252 models in total. Details of the participants meeting our inclusion criteria concerning both adequate survey and MRI data quality are shown in Table 1.

**Table 1.** Sample characteristics are reported for each model.

| Modality | Characteristics | Sexual Trauma Models | Interpersonal Physical Violence Trauma Models |
| --- | --- | --- | --- |
| fMRI | Total *n* | 538 (196 Exposed) | 240 (All exposed) |
|  | Lifetime PTSD *n* | 103 | 60 |
|  | Mean Age, years (SD) | 29.60 (7.72) | 31.53 (7.34) |
|  | Male *n* (%) | 230 (43%) | 102 (43%) |
| GMV | Total *n* | 670 (257 Exposed) | 302 (All exposed) |
|  | Lifetime PTSD *n* | 128 | 74 |
|  | Mean Age, years (SD) | 29.60 (7.67) | 31.37 (7.40) |
|  | Male *n* (%) | 269 (40%) | 127 (42%) |

Of the 252 models tested, 239 (95%) were deemed acceptable representations of the data (i.e., χ^2^ *p* > .05). Only parameters of interest concerning models with acceptable fit were retained for FDR correction (see Supplementary Material 1.9 for an overall evaluation of the models tested). Supplementary Material 6 reports all parameters across all models (except for Mediation analysis results – see Supplementary Material 7).

### 4.3 Direct effects of Trauma on Neurobiological Phenotypes (Path A)

We first examined the main effects of trauma dose on neurobiological phenotypes - that is, Path *A* parameters. We only considered path *A* effects that were consistently detected independent of the outcome variable in the mediation model (see section 3.3). Our analysis revealed that exposure to Sexual Trauma did not predict individual differences in GMV or FC across any of the retained PCs and that this effect was not moderated by survivor sex. Post hoc sensitivity analyses, in which models were re-estimated in the subgroup exposed to Sexual Trauma only, also failed to identify significant findings following FDR correction.

In contrast to Sexual Trauma, more exposure to Interpersonal Physical Violence was associated with higher volumes in the ventral anterior thalamus, anterior caudate, nucleus accumbens shell, and lateral temporal and occipital regions (Fig 2). Exposure-related volume reductions were concentrated in dorsolateral prefrontal, parietal, and diffuse medial regions, as well as dorsoposterior thalamic nuclei and the amygdala (Figure 2A and 2B). More exposure to Interpersonal Physical Violence was also linked to stronger FC between default, control, visual, and attentional systems, along with stronger anticorrelations within each network and between most other pairs of networks, including between default subsystems, default and limbic/thalamic systems, and somatomotor and attention/limbic networks (Figure 2C).

**Figure 2.**
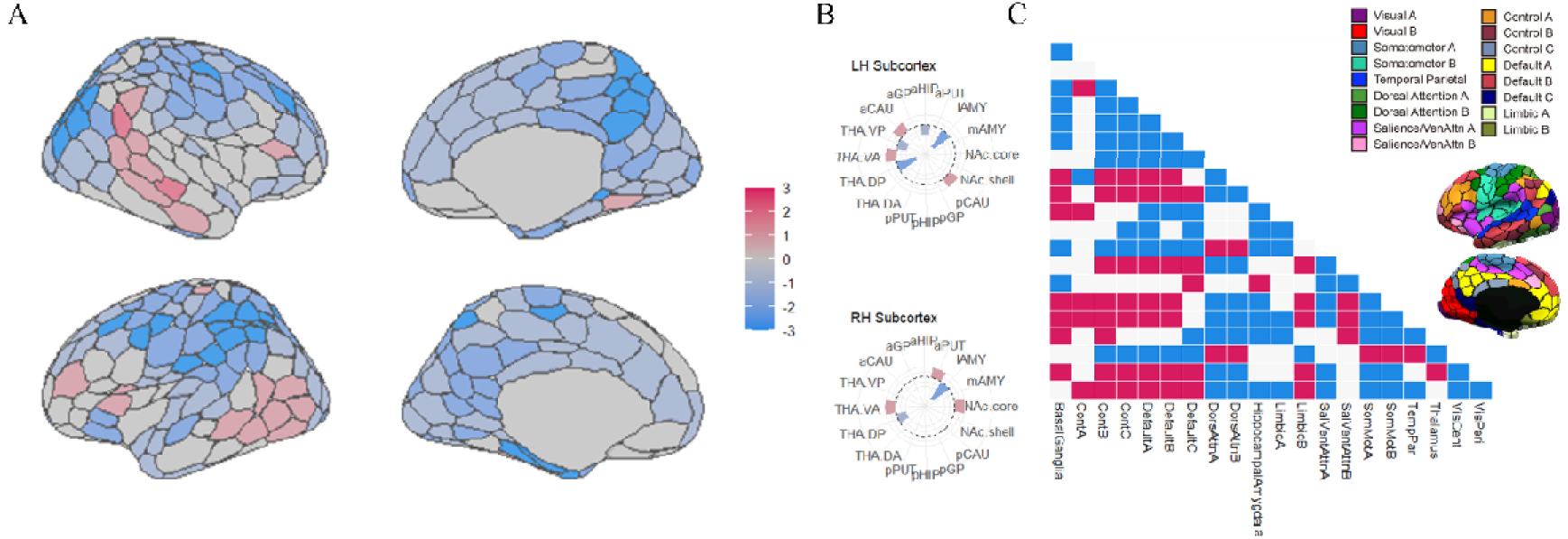
Direct main effects of interpersonal violence trauma dose on cortical regional GMV (A), subcortical regional GMV (B) and inter-network FC (C) aggregated across PCs. Red indicates cortical volume expansions or increased FC in association with greater exposure; Blue indicates cortical volume reductions or greater anticorrelated FC. Atlas in (C) adapted from previous work (Schaefer et al., 2018). See https://github.com/yetianmed/subcortex/ for nomenclature in (B).

We next considered the moderating effect of age of first exposure. The analysis of Interpersonal Physical Violence revealed that a single exposure at any developmental stage was not sufficient to yield detectable neurobiological changes (i.e., we found no significant effects of single exposure for any age of onset investigated; see age of onset simple intercept parameter results in Supplementary Material 6). However, dose-response effects of violence depended on age of onset, such that individuals exposed up to preadolescence exhibited greater ventral occipitotemporal volume reductions with higher exposure rates, suggesting heightened sensitivity to early and frequent trauma of this type within this specific brain region (Fig 3A). Individuals with later onsets (up to adulthood) displayed more widespread dose-dependent volume reductions, which were most evident across the medial cortical wall, the dorsolateral prefrontal cortex, and the parietal lobe, with some reductions also detected for the dorsoposterior thalamic nuclei and amygdalae. Volume increases were also more pronounced in lateral temporal and occipital regions given later first ages of onsets (see Figure 3A and 3B). These maps thus resembled an exaggeration of the main effect of Interpersonal Physical Violence on GMV. The analysis of FC revealed that more exposure to violence, coupled with later first ages of onset, was associated with stronger FC between most network pairs. The exceptions were links between default mode subsystems and between visual and somatomotor networks, which became more anticorrelated (Fig 3C).

**Figure 3.**
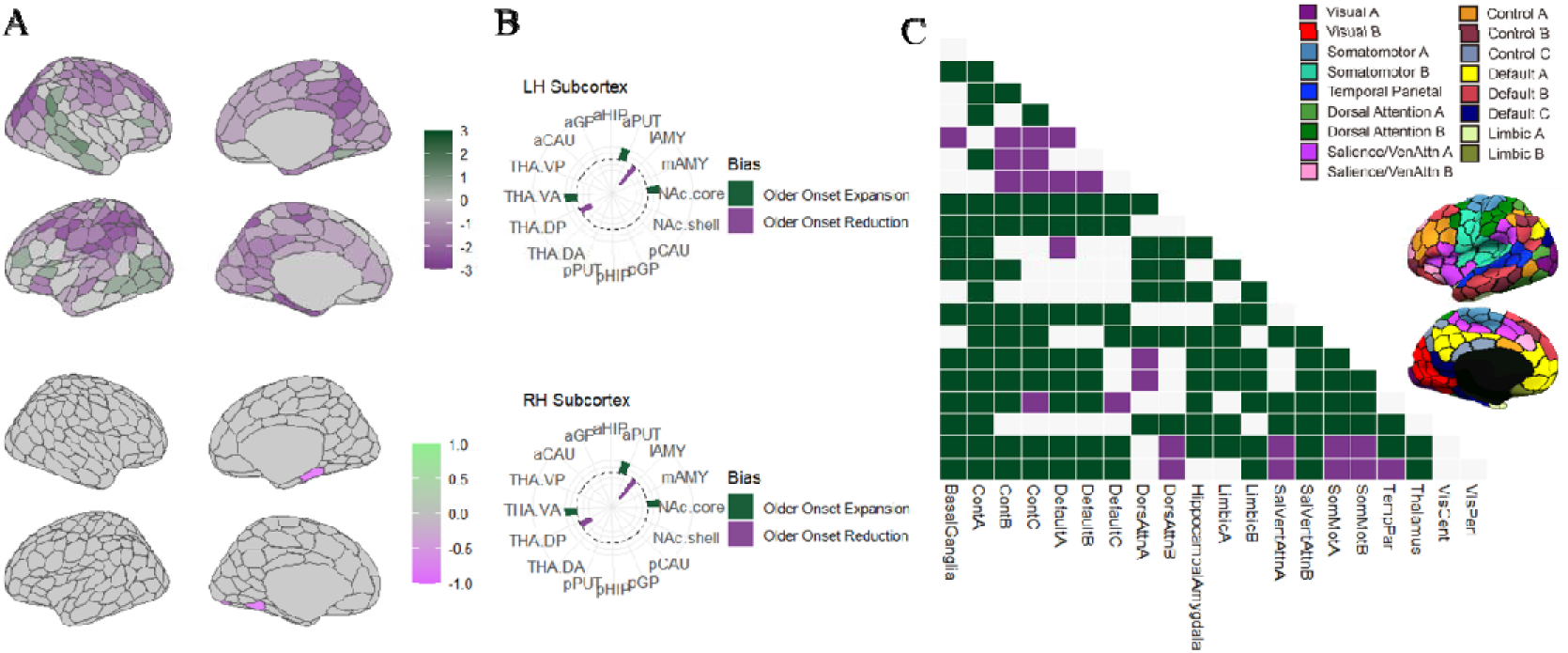
Effects of interpersonal violence trauma dose on (A) cortical regional GMV, (B) subcortical regional GMV, and (C) inter-network FC aggregated across PCs, shown as a function of age of onset. In (A), the top panel illustrates cortical pattern changes more pronounced with later first exposures (up to adulthood), while the bottom panel illustrates the only trend where dose-responses were stronger for early-developmental exposure relative to later first ages of exposure (trend strongest up to preadolescence). In (B) and (C), relevant effects were always more pronounced given later first exposures relative to early first exposures. Green indicates cortical volume expansions or increased FC with greater exposure; darker greens denote stronger effects with later first exposure. Purple indicates cortical volume reductions or greater anticorrelated FC; darker purples denote stronger effects with later first exposure. Atlas in (C) adapted from previous work (Schaefer et al., 2018). See https://github.com/yetianmed/subcortex/ for nomenclature in (B).

The dose-response effect of Interpersonal Physical Violence on the brain was also moderated by survivor sex (Figure 4). Males exhibited more widespread exposure-related cortical reductions, particularly across medial occipital, dorsolateral prefrontal cortical, and parietal regions, alongside more pronounced expansions in ventrolateral prefrontal, temporo-occipital and subcortical regions. Exposure-related FC increases between control sub-systems, between control and default subsystems, and between limbic and default areas were also more pronounced in males. In contrast, females showed stronger dose-related GMV reductions in less diffuse ventrolateral prefrontal and medial parietal cortices, with FC changes for females not being more pronounced than that detected for males.

**Figure 4.**
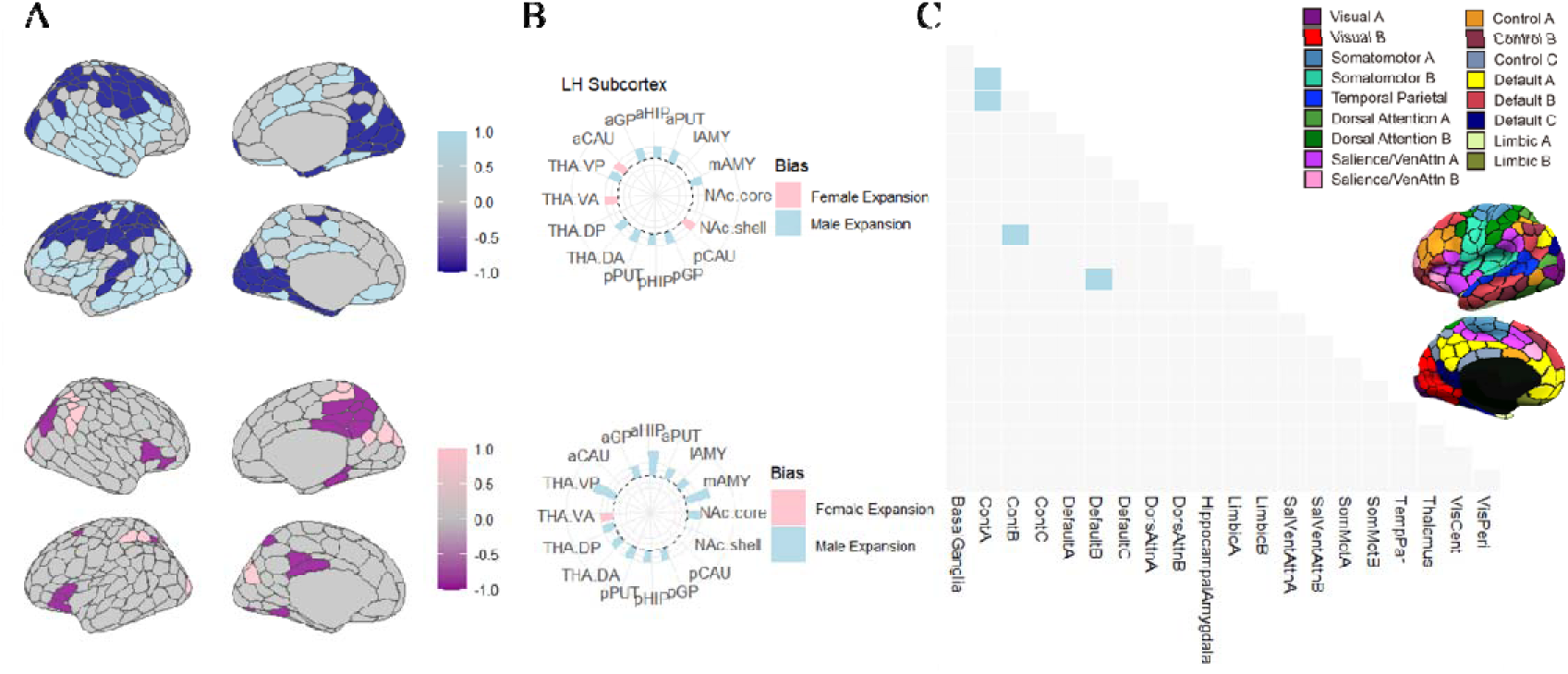
Effects of interpersonal violence trauma dose on (A) cortical regional GMV, (B) subcortical regional GMV, and (C) inter-network FC aggregated across PCs, shown as a function of sex. In (A), the top panel illustrates cortical pattern changes more pronounced in males, while the bottom panel illustrates changes more pronounced in females. In (C), relevant effects were always more pronounced in males. Light blue/pinks indicate cortical volume expansions or increased FC with greater exposure. Dark blues/magentas indicate cortical volume reductions or greater anticorrelated FC. Overall, blues indicate that the trend is more pronounced in males; pinks indicate that the trend is more pronounced in females. Atlas in (C) adapted from previous work (Schaefer et al., 2018). See https://github.com/yetianmed/subcortex/ for nomenclature in (B).

### 4.4 Direct effects of Neurobiology on Psychopathology (Path B)

We found no evidence for direct effects of GMV or FC on psychopathological outcomes independent of model specification. We did observe that fMRI PC17 predicted Lifetime PTSD in the Interpersonal Physical Violence model (post-FDR) and in the post-hoc Sexual Trauma sensitivity models (pre-FDR only; both β ≈ – 0.20). In this context, lifetime PTSD presence was linked to stronger anticorrelations between default subsystems and temporoparietal, dorsal attention, and limbic networks, as well as stronger anticorrelations between broad visual and ventral attentional networks. The trend also implicated increased FC between peripheral visual and limbic networks. However, these results should be considered tentative given the lack of consistency across model specifications.

### 4.5 Testing for mediation effects

We found no robust evidence supporting statistically significant neurobiological mediation of the effects of either Sexual Trauma or Interpersonal Physical Violence on psychopathological outcomes (see Supplementary Material 7). For FC, these findings did not depend on the parcellation used (Supplementary Material 2).

There was some evidence that certain GMV and FC PCs mediated the effects of Interpersonal Physical Violence on Lifetime PTSD at *lenient* thresholds (i.e., 97.5% and 99% CI), but these effects should be interpreted with caution given the large number of models considered here (see Supplementary Material 1.10) and the fact that the confidence intervals barely excluded zero, indicating borderline evidence at best (Gelman & Stern, 2006). Across the retained mediation models, we evaluated 1,694 mediation-related parameters (239 unmoderated indirect effects; 478 sex-stratified indirect effects; 615 age-of-onset–specific indirect effects; 362 indices of moderated mediation). Controlling the familywise error rate at 0.05 with the conservative Bonferroni adjustment yields a per-test α ≈ 2.75 × 10□□, equivalent to requiring ≈ 99.997% CIs (Bland & Altman, 1995). Thus, even “conservative” 99% intervals are insufficiently stringent here, especially given the large number of non-mediation parameters also estimated and excluded from this correction calculation. Strength of evidence for mediation was also similarly weak in the Schaefer100 + Tian S2 sensitivity analysis. Furthermore, evidence for GMV-based mediation was inconsistent when effects were re-estimated using standardised rather than unstandardised coefficients, suggesting that the initially apparent mediation was contingent on arbitrary scaling choices (see Supplementary Material 1.10). Taken together, any apparent mediation effects were at most marginally significant and disappeared under reasonable analytic variations, suggesting they were spurious (Leamer, 1983; Simonsohn et al., 2020; Steegen et al., 2016).

## Discussion

Many studies implicitly assume that the influence of trauma on psychopathology is at least partly mediated by the commonly documented effects of trauma on the brain, but this neurobiological mediation model is seldom tested directly. In this study, we comprehensively tested for neurobiological mediation of the effects of distinct trauma types on dimensional and categorical psychopathological outcomes while considering the moderating influences of contextual factors such as survivor sex and age of onset. We broadly replicated prior work linking Interpersonal Physical Violence to altered GMV and FC (Admon et al., 2013; Cassiers et al., 2018; L. Li et al., 2014; Nkrumah et al., 2023; Tomoda, Suzuki, et al., 2009) by demonstrating that dose–response effects vary according to survivor sex and the developmental timing of trauma onset. In contrast, Sexual Trauma showed no robust associations with neurobiological phenotypes, which failed to reliably predict psychopathology. We also failed to converge on any robust evidence of mediation, regardless of whether the overall psychopathological outcome was indexed dimensionally (via two broadband transdiagnostic psychopathological traits previously linked to trauma exposure) (Constable, Tiego, Pavlovich, Tran, et al., 2025), or categorically, via lifetime PTSD diagnosis. Our analysis thus favours an independent effects model for GMV and FC, in which specific forms of trauma exert distinct influences on the brain and mental health, over the commonly assumed neurobiological mediation hypothesis.

### 5.1 The direct effects of Interpersonal Physical Violence on GMV and FC

Greater exposure to Interpersonal Physical Violence predicted increased volume in specific thalamic and striatal areas, in addition to lateral temporal and occipital regions, along with more widespread volume reductions concentrated in prefrontal, parietal, and medial occipital cortices. Increased exposure was also linked to greater FC between default, control, visual and attentional systems and more pronounced anticorrelations across most other pairs of systems. The observed parietal and prefrontal volume losses are consistent with recent studies of child abuse (Nkrumah et al., 2023) and other physically abused samples (Cassiers et al., 2018; Tomoda, Suzuki, et al., 2009). Occipital reductions and disrupted limbic–prefrontal coupling similarly parallel well-established findings in PTSD (Admon et al., 2013; L. Li et al., 2014), although evidence for such functional changes in the context of physical abuse specifically has been mixed (Cisler, 2017). The role of occipital regions across trauma types appears relatively underexplored (Cassiers et al., 2018).

Our analysis indicates that these associations varied by the developmental timing of trauma. Specifically, while we found that a single exposure at any stage of development was not consistently linked to neurobiological change, for both individuals first exposed early and later in development, higher levels of trauma exposure were related to more pronounced effects on the brain. This does not appear to have been previously found in the literature. This pattern aligns with our previous findings in this sample showing that escalating or repeated Interpersonal Physical Violence has a stronger impact on mental illness severity than a single incident (Constable, Tiego, Pavlovich, Tran, et al., 2025). In ventral occipitotemporal regions, cumulative violence exposure was linked to greater reductions for individuals first exposed up to preadolescence, suggesting that early trauma may heighten vulnerability of this system under repeated exposure. Such regions are implicated in visual object and face recognition (Kravitz et al., 2013), although the precise role that occipitotemporal volume reductions have in the context of PTSD, for example, remains unclear (L. Li et al., 2014). We also caution against speculation of the behavioural consequences of such brain changes.

Individuals first exposed later in development showed steeper dose–response trajectories across a broader range of cortical and subcortical regions, potentially reflecting the effects of more recent chronic trauma exposure and reduced opportunities for neural recovery. This interpretation is consistent with evidence that recent exposure increases risk for psychopathology (Dunn et al., 2017), although caution must be exercised as we did not explicitly measure time since last trauma exposure. Notably, the pattern observed in this context resembled an exaggerated form of the main effect of Interpersonal Physical Violence on the brain.

We also found that the relationship between Interpersonal Physical Violence exposure and neurobiology varied as a function of survivor sex. Females generally exhibited stronger dose-related volume reductions in focal parietal and ventrolateral prefrontal regions, alongside more pronounced expansions in select thalamic and striatal nuclei. Males showed greater dose-dependent grey matter volume increases across widespread subcortical, temporal, lateral occipital and prefrontal regions, accompanied by more pronounced reductions across broad parietal and medial occipital cortices. Although prior work has emphasised that sex-related heterogeneity is critical to consider in trauma research, it is seldom addressed in practice (Cassiers et al., 2018).

Much of the existing literature on sex differences has focused exclusively on early trauma without stratifying by trauma type, and in some cases, only while focusing on individuals meeting criteria for conventionally defined psychopathological diagnoses (Helpman et al., 2017a), which complicates direct comparisons with our findings. For example, early trauma in males has been linked to volume loss in prefrontal, amygdala, and hippocampal regions; in females it has been associated with amygdala enlargement (Helpman et al., 2017a; Herzog & Schmahl, 2018). These divergences are thought to stem from sex differences in stress-related and gonadal hormone systems (Fonkoue et al., 2020; Helpman et al., 2017b; Stevens et al., 2018). Our more granular modelling approach, focusing on dose-response effects of Interpersonal Physical Violence experienced across the lifespan, revealed a different pattern, in which males showed diffuse cortical reductions with concurrent temporal, prefrontal, and subcortical expansions, whereas females showed more focal prefrontal and parietal reductions with limited subcortical involvement. Notably, lower parietal volumes emerged consistently across our models, aligning with evidence of the central importance of parietal volume reductions in child physical abuse, even when controlling for other trauma types such as neglect (Nkrumah et al., 2023).

Our results suggest that the degree to which lower parietal volumes are observed in such contexts is modulated by survivor sex and age of trauma exposure.

### 5.2 Distinct developmental windows of sensitivity and sex-specific vulnerabilities to interpersonal violence–related traumatic stress

Our findings also indicate that there are distinct developmental windows of sensitivity and sex-specific vulnerabilities to interpersonal violence–related traumatic stress. Ventral occipitotemporal regions appear more susceptible to dose-response effects when exposure first begins early in development (up to preadolescence), whereas broader cortical–subcortical dose-response associations emerge when exposure begins later (or, speculatively, when trauma is more recent). Furthermore, males appear more vulnerable to diffuse cortical remodelling than females in the context of interpersonal physical violence. These findings appear novel, and converge with behavioural evidence indicating that, at escalating levels of such exposure, males are more susceptible than females to more severe psychopathology (Constable, Tiego, Pavlovich, Tran, et al., 2025).

It is possible that such direct effects of Interpersonal Physical Violence on the brain are at least partly explained by injuries sustained during the trauma. Head and neck injury is capable of producing widespread neuroanatomical alterations (B. L. Brett et al., 2022) and is frequently observed in survivors of physical violence attending medical care (Brink et al., 1998; Injury in Australia, 2025; Zohrevandi et al., 2024), but we excluded individuals with a history of significant head trauma occasioning loss of consciousness. Some studies suggest that repeated minor sub-concussion may yield subtle neurobiological effects (Delang et al., 2025), yet this remains a relatively underdeveloped area of the literature, and there were no clinically significant abnormalities in any of our subjects based on expert radiologist inspection. These considerations suggest that the effects we identify are more likely related to the possible neurotoxic effects of glucocorticoids released during the stress response cycle (Sapolsky et al., 1990), although a more detailed characterisation of the survivors’ medical history around the time of the trauma would be required to evaluate this possibility.

### 5.3 The direct effects of Sexual Trauma on the brain

We found no direct effects of Sexual Trauma on any of the neurobiological phenotypes tested. Prior studies have reported associations between childhood sexual abuse and reduced hippocampal, frontal, occipital, parietal, and caudate volumes (Andersen et al., 2008; Cassiers et al., 2018; Heim et al., 2013; Tomoda, Navalta, et al., 2009). However, most of these studies were based on small samples (often <100), a limitation that has been widely recognised as problematic in neuropsychiatric research due to the heightened risk of spurious or unstable findings (Button et al., 2013). Furthermore, many have sought to isolate “pure” effects of Sexual Trauma by excluding individuals with exposures to other trauma types, which may yield atypical subgroups given that exposure to multiple trauma types is the norm rather than the exception for trauma survivors (Finkelhor et al., 2007; Turner et al., 2010). Our use of a larger, more generalisable sample, in which polytrauma exposure was statistically controlled rather than used as a basis for exclusion, may thus be more representative of the population of people affected by this trauma.

The lack of an effect of Sexual Trauma on the brain is nevertheless notable given prior evidence in this sample that even a single instance of Sexual Trauma is sufficient to raise psychopathology risk (Constable, Tiego, Pavlovich, Tran, et al., 2025). Relative to interpersonal physical violence, however, Sexual Trauma remains especially prone to underreporting due to secrecy, shame, and stigma (Baldwin et al., 2019), and tends to encompass a wider scope of experiences (e.g., coercive Vs. forced) (Masters et al., 2015). Capturing and modelling such experiential heterogeneity may be important for improving the reliability of neurobiological estimates. Further research is needed to determine whether the apparent null associations between Sexual Trauma and GMV/FC detected here reflect a true absence of effect, given our large sample size compared to many past studies, or limitations in our ability to capture the experiential heterogeneity of Sexual Trauma.

### 5.4 Associations between Neurobiology and Psychopathology

We observed limited and inconsistent evidence for statistically significant associations between neurobiological measures and psychopathology, and these associations were generally weaker than those linking trauma exposure and neurobiology. This result aligns with a growing realization that brain imaging phenotypes often correlate weakly with psychometric measures of psychopathology (Marek et al., 2022; Ooi et al., 2022; Pavlovich et al., 2024). In this context, it is possible that the neurobiological phenotypes we considered here, despite being among those most widely investigated in the psychotraumatology and psychiatric literature, lack sensitivity for revealing associations with behaviour. For example, while a small number of studies have reported associations between trauma exposure and regional neural activity and FC, the majority, particularly those using resting-state data, have found no such link (Cassiers et al., 2018). Recent work suggests that measures of brain activity obtained during active engagement, either with movies or cognitive tasks, can improve effect sizes in brain-behaviour correlation analyses (Eickhoff et al., 2020; Finn & Bandettini, 2021; Jääskeläinen et al., 2021; Meer et al., 2020). The use of alternative imaging modalities may also be helpful in this regard. For instance, positron emission tomography can be used to probe neurotransmitter densities relevant for mental illness and recently developed tracers can index synaptic density (Meyer et al., 2020; Singh et al., 2024; Zheng et al., 2025), which is strongly modulated by physiological stress responses (Bath et al., 2017; Woodburn et al., 2021). However, the financial cost and invasive nature of such assays may preclude their deployment in large samples. Future work could consider the possible mediating role of such neurophenotypes, which remain comparatively underexplored in the trauma–neuroscience literature.

### 5.5 A lack of evidence for neurobiological mediation

We found no robust evidence that GMV and FC changes mediate the link between psychological trauma exposure and psychopathology, despite the heavy emphasis on these neurophenotypes in the psychotraumatology literature. While some previous studies have reported evidence for mediation (Chen et al., 2024; Harb et al., 2024; Opel et al., 2019; Wan et al., 2022), these were generally conducted in small samples (ranging from 93 to 152 participants, inflating the risks Type 1 and II error) (Button et al., 2013). For an exception, see Wan et al., 2022). The smallest subsample in our analyses was double the average sample size in prior such mediation studies, making it unlikely that power limitations alone explain our differing conclusions. Even if much larger samples are required to detect mediation via GMV or FC changes, we contend that any true effect would be very small and confined to specific subgroups of survivors. Indeed, hypothesised mediation pathways are rarely followed uniformly across all individuals (i.e., population or sample-level findings rarely explain all individual-level trends in mediation studies), particularly when each of the model path estimates effect sizes are small (Bogdan et al., 2024), as expected in neuroimaging research (Marek et al., 2022; Tiego et al., 2023). For example, while our hypothesised pathway assumed that higher trauma load leads to greater neurobiological change, which in turn predicts more severe psychopathology, it appears that most individuals instead display different patterns of relationships - for example, demonstrating evidence of high trauma load and severe psychopathology with few associated detectable neurobiological deviations.

Much of the prior research claiming evidence for mediation has also collapsed distinct forms of adversity into broad childhood maltreatment composites prior to testing for mediation. It is therefore possible that aggregate adversity, rather than trauma subtype, is the critical driver of neurobiological change and subsequent psychopathology, meaning that our disaggregated approach obscured evidence of mediation. However, distinct forms of trauma exert differential effects on psychological outcomes (Constable, Tiego, Pavlovich, Tran, et al., 2025; Hyland et al., 2022; Keyes et al., 2012) and different forms of maltreatment have also been are linked to divergent neural profiles, with findings depending in part on how trauma types are defined and differentiated (Paquola et al., 2016). A more detailed consideration of the differential effects of distinct forms of trauma on the brain and mental health is therefore required in mediation studies.

### 5.6 Limitations and Conclusions

A key limitation of the present study is the lack of information regarding the recency of trauma exposure, which is an important moderator of trauma outcomes (Dunn et al., 2018). This information would clarify the relative contributions of exposure recency and developmental timing. We also lacked data on the temporal ordering of trauma exposure relative to the emergence of psychiatric issues or neurobiological changes. In cross-sectional mediation analyses, one cannot confidently claim that the mediator ‘causes’ the exposure, or whether the outcome ‘causes’ the mediator (‘reverse-causation’) (Y. Li et al., 2023) – especially where we report results where Lifetime PTSD is an outcome variable. Future research should employ longitudinal, prospective designs to mitigate biases inherent in retrospective self-reports whilst offering greater confidence in effect directionality. Analysis of large-scale prospective developmental cohorts akin to the ABCD study may be helpful here (Casey et al., 2018). This approach is also likely to support greater precision in participant accounts of ages of exposure and amounts of exposure experienced. Although some studies have examined these relations prospectively, they inherit many of the limitations of cross-sectional work in this area. Typically, such studies (i) do not disaggregate maltreatment subtypes to ensure trauma□subtype-specific inference for the purported mediation effects (e.g., physical vs. emotional abuse/name-calling vs. sexual abuse; Brieant et al., 2021; Edalati et al., 2023; Liu et al., 2025); and (ii) operationalise “adversity” using broad composites that mix traumatic with potentially non-traumatic experiences (e.g., including “a family member had a mental or emotional problem”, or “Has ANY blood relative of your child ever suffered from depression?” Brieant et al., 2021, Liu et al., 2025). Longitudinal analyses in sufficiently large samples that rely on improved measures of trauma and outcomes will be an important direction of future research.

Our study used a transdiagnostic sample with nationally representative rates of trauma exposure (Australian Bureau of Statistics, 2021; Mathews et al., 2023) and was designed to allow comparison of dimensional and categorical psychological outcomes while considering the effects of trauma type, dosage, age of onset, and survivor sex. Our findings suggest that interpersonal physical violence, but not sexual violence, is associated with variability in GMV and FC across several cortical and subcortical regions, particularly within parietal areas, in a way that depends on dosage, age of onset, and survivor sex. Critically, we found no evidence to indicate that these neurobiological effects mediate the influence of trauma on psychopathology, thus supporting an independent effects model over FC/GMV neurobiological mediation hypotheses.

## Data Availability

The data used in the present paper was collected as part of an ongoing project which is not yet publicly available. The project data will be released upon completion; inquiries may be directed to the senior author.

## Author contributions

TC, JT, and AF conceptualised the study, interpreted the results, and wrote the manuscript. TC prepared and analysed the data. KP, AS, YS, and PTL reviewed the code for data analysis. JT, NOT, BH, RO, JK, KF, KT, SB, JM, ME, RF, MB, and AF contributed to data acquisition and project oversight. AF was supported by the Australian Research Council (ID: FL220100184) and National Health and Medical Research Council (ID: 1197431). Jeggan Tiego was supported by NHMRC Investigator Grant 2033976.

## Code availability

Project code can be found at: https://github.com/cicadawing/Mediation-Study.Trauma-Neurobiology-and-Transdiagnostic-Mental-Health

