## Supplementary 1 for "The effects of trauma on neurobiological and psychopathological phenotypes in a transdiagnostic community sample"

**Section S1.1 – Sample Characteristics, Exclusion and Inclusion Criteria**

A total of 1110 participants (Male *n* = 440; 30.63%) were recruited from the general community via an online campaign targeting various social media and other outlets coordinated by the private company Trialfacts (https://trialfacts.com/). Written informed consent was required before study participation. Inclusion criteria were aged 18-45 years; right-handedness; European ancestry (defined as all four grandparents of European descent, which was necessary given that the broader project also involves genetics analysis); English as the first spoken language; no intellectual disability or learning disorder (e.g., dyslexia); no history of frequent headaches or migraines, no past seizures, concussions, or loss of consciousness that lasted more than 3 minutes; no neurological illness or history of neurosurgery; normal or corrected-to-normal vision; no metal in the body; not currently pregnant or attempting to conceive; no history of receiving hormone blockers or hormone replacement; no history of gender-affirming surgery; no history of steroid abuse; no exposure to transcranial magnetic stimulation (TMS) in the past six months, no more than two weeks of exposure to TMS in the past year, and no more than three months of continuous exposure to TMS in their lifetime; and no prior exposure to electroconvulsive therapy. Experience of psychiatric symptoms or past/current psychiatric diagnosis and/or treatment were not used as a base for exclusion, although a sub-group (N=527) was recruited specifically because they had no history of psychiatric treatment. Specifically, exclusion criteria for this group were identical to the rest of the sample, but they also had no history of psychiatric hospitalisation or substantial psychiatric medication use - defined as no psychiatric medication in the past 6 months, no more than 2 weeks of exposure in the past year, and no more than three months of continuous exposure to psychiatric medication at any point in their life. In the present context, this approach allowed use to investigate the effects of being exposed to trauma or not, in addition to the dose-response impacts within exposed individuals.

Key demographic details of the sample are provided in Table 1. The study was approved by the Monash Human Research Ethics Committee (Project ID: 12692 & 43629). Table 1 below details information on dataset demographics pre and post data-cleaning of the behavioural data.

Table 1 – Participant details pre and post removal of attention check outliers and gender non-conforming people

|  | Original Dataset | | Post Cleaning | |
| --- | --- | --- | --- | --- |
| Percent Male | 340 [30.63%] | | 319 [37.89%] | |
| Percent Female | 549 [49.46%] | | 523 [62.11%] | |
| Percent Other Gender | | 36 [3.24%] | | 0 [0%] |
| Percent No Gender Response | | 185 [16.67%] | | 0 [0%] |
| Percent TAFE/National Certificate/Diploma Educated | 93 [8.38%] | | 85 [10.1%] | |
| Percent Bachelor Educated | 112 [10.09%] | | 108 [12.83%] | |
| Percent Secondary Educated | 53 [4.77%] | | 47 [5.58%] | |
| Percent Masters Educated | 218 [19.64%] | | 189 [22.45%] | |
| Percent Bachelor with Honours Educated | 170 [22.7%] | | 238 [28.27%] | |
| Percent Graduate Diploma Educated | 4 [15.32%] | | 155 [18.41%] | |
| Percent Primary Educated | 17 [0.36%] | | 3 [0.36%] | |
| Percent PhD Educated | 17 [1.53%] | | 17 [2.02%] | |
| Mean Age | 29.58 | | 29.73 | |
| Total Sample Size | 1110 | | 842 | |

All 842 of participants included in our analysis after data cleaning were asked: “Please select your gender,” with response options: ‘Male’, ‘Female’, ‘Nonbinary/Gender Diverse’, and ‘My gender is not listed. I identify as:’, with an open-text field available for the latter two responses. A subset of 761 were also asked about their biological sex at birth. To maximize our sample size, we relied on the first question to assign participant sexes and retained only participants who responded Male or Female, under the assumption that non-cis-gender individuals would endorse one of the other responses. The agreement in responses to the gender-based and sex-based questions in the subset of 761 individuals who completed both and endorsed Male or Female as their gender was 98.5%. Given the use of robust estimators and Monte Carlo simulations to estimate confidence intervals, any residual misclassification in sex assignment for the remaining ~1.5% of the sample is expected to exert negligible influence on the reported parameter estimates and the conclusion that trends reflect sex rather than gender differences. Our prior analyses in behavioural-level data alone also indicated that error in sex classification cannot explain our findings (Constable, Tiego, Pavlovich, Tran, et al., 2025).

**Section S1.2 – Cronbach’s Alpha Results**

Table 2 – Cronbach’s alpha results for each of the subscales of interest in the 842 participants with clean behavioural data

| *CAT-PD Subscales:* |  |  |  |  | *BFI-II Subscales:* |  |
| --- | --- | --- | --- | --- | --- | --- |
| Lability | 0.84 |  |  |  | Extraversion | 0.86 |
| Anger | 0.85 |  |  |  | Agreeableness | 0.79 |
| Anhedonia | 0.85 |  |  |  | Conscientiousness | 0.88 |
| Anxiousness | 0.90 |  |  |  | Negative_Emotionality | 0.93 |
| Callousness | 0.86 |  |  |  | Open_Mindedness | 0.84 |
| Cognitive_Problems | 0.87 |  |  |  | Sociability | 0.84 |
| Depressiveness | 0.90 |  |  |  | Assertiveness | 0.78 |
| Domineering | 0.79 |  |  |  | Energy_Level | 0.74 |
| Emotional_Detachment | 0.88 |  |  |  | Compassion | 0.56 |
| Exhibitionism | 0.82 |  |  |  | Respectfulness | 0.68 |
| Fantasy_Proneness | 0.81 |  |  |  | Trust | 0.71 |
| Grandiosity | 0.78 |  |  |  | Organization | 0.81 |
| Health_Anxiety | 0.80 |  |  |  | Productiveness | 0.77 |
| Hostile_Aggression | 0.76 |  |  |  | Responsibility | 0.75 |
| Irresponsibility | 0.83 |  |  |  | Anxiety | 0.84 |
| Manipulativeness | 0.82 |  |  |  | Depression | 0.85 |
| Misstrust | 0.86 |  |  |  | Emotional_Volatility | 0.89 |
| Nonperseverence | 0.88 |  |  |  | Intellectual_Curiosity | 0.67 |
| Nonplanfulness | 0.86 |  |  |  | Aesthetic_Sensitivity | 0.75 |
| NormViolation | 0.80 |  |  |  | Creative_Imagination | 0.75 |
| Peculiarity | 0.86 |  |  |  |  |  |
| Perfectionism | 0.84 |  |  |  |  |  |
| Relationship_Insecurity | 0.85 |  |  |  |  |  |
| Rigidity | 0.79 |  |  |  |  |  |
| Risk_Taking | 0.87 |  |  |  |  |  |
| Romantic_Disinterest | 0.87 |  |  |  |  |  |
| Rudeness | 0.86 |  |  |  |  |  |
| SelfHarm | 0.88 |  |  |  |  |  |
| Social_Withdrawal | 0.78 |  |  |  |  |  |
| Submissiveness | 0.82 |  |  |  |  |  |
| Unusual_Beliefs | 0.65 |  |  |  |  |  |
| Unusual_Experiences | 0.78 |  |  |  |  |  |
| Workaholism | 0.88 |  |  |  |  |  |

**Section S1.3 – PC’s Extracted from BFI-II and CAT-PD Data**

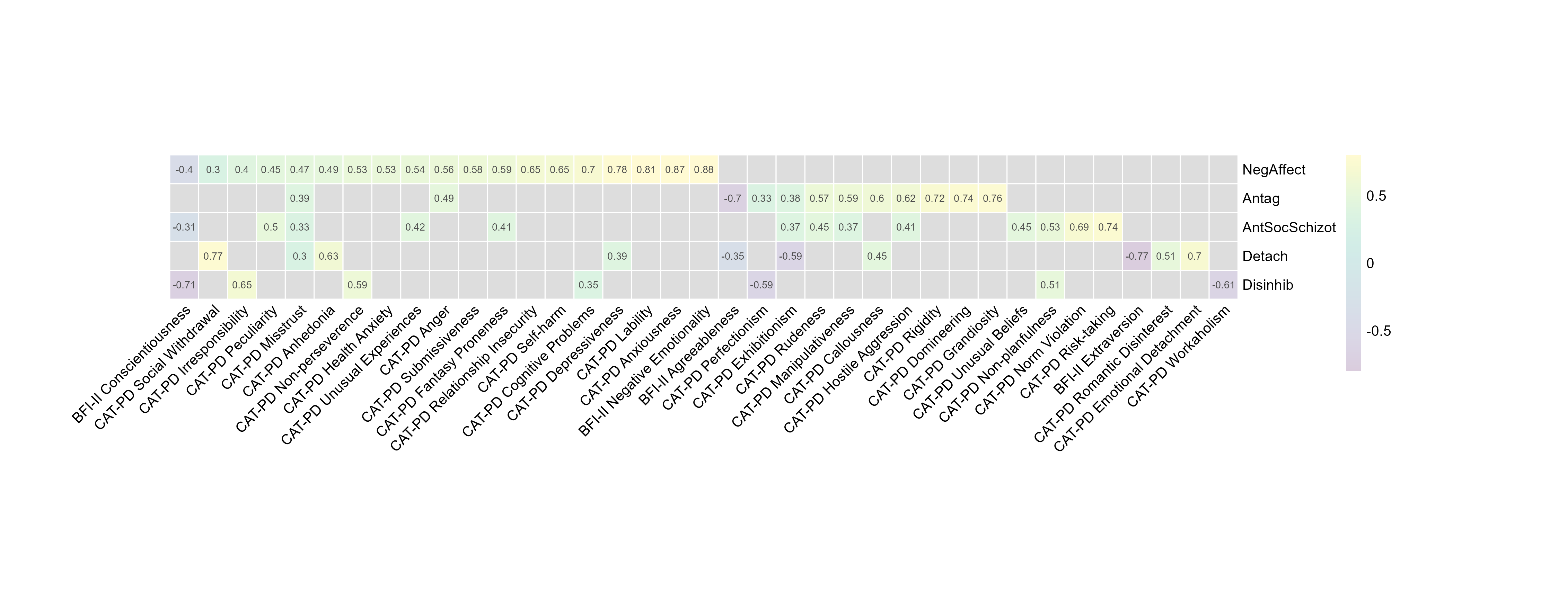

Figure 1. Figure describes the pattern and strength of component loadings for each component extracted in Tier 5 of the Bass-Ackwards hierarchy. Component loadings < .3 have been greyed. NegAffect - Negative Affectivity, Antag - Agreeableness to Antagonism, Disinhib - Conscientiousness to Disinhibition, AntSocSchizot - Antisocial Schizotypy, and Detach - Extraversion to Detachment. Loadings below an absolute value of 0.3 have been omitted from the table for interpretability (Costello & Osborne, 2005; Tabachnick & Fidell, 2013). These same PCs are similarly reported in (Constable, Tiego, Pavlovich, Tran, et al., 2025).

**Section S1.4 – THQ Item Grouping**

Table 3. THQ organisation. The supposed subscale organisation intended by instrument creators is described in the far-left column. Item regroupings are illustrated according to a key described at the bottom of the table. Please also see (Constable, Tiego, Pavlovich, Tran, et al., 2025).

| Original Subscale Structure | THQ item | Description | Item Regrouping |
| --- | --- | --- | --- |
| Crime-related events | 1 | Has anyone ever tried to take something from you by using force or the threat of force, such as a stick-up or mugging | □ |
|  | 2 | Has anyone ever attempted to rob you or actually robbed you (ie., stolen your personal belongings)? | □ |
|  | 3 | Has anyone ever attempted to or succeeded in breaking into your home when you were not there? |  |
|  | 4 | Has anyone ever attempted to or succeeded in breaking into your home while you were there? | □ |
| General disaster and other trauma related events | 5 | Have you ever had a serious accident at work, in a car, or somewhere else? | **✰** |
|  | 6 | Have you ever experienced a natural disaster such as a tornado, hurricane, flood or major earthquake, etc. where you felt you or your loved ones were in danger of death or injury? | **✰** |
|  | 7 | Have you ever experienced a 'man-made' disaster such as a train crash, building collapse, bank robbery, fire, etc, where you felt you or your loved ones were in danger of death or injury? | **✰** |
|  | 8 | Have you ever been in any other situation in which you were seriously injured? | **✰** |
|  | 9 | Have you ever been in any other situation in which you feared you might be killed or seriously injured? | **✰** |
|  | 10 | Have you ever seen someone seriously injured or killed? | **✰** |
|  | 11 | Have you ever seen dead bodies (other than at a funeral) or had to handle dead bodies for any reason? |  |
|  | 12 | Have you ever had a close friend or family member murdered, or killed by a drunk driver? | ▩ |
|  | 13 | Have you ever had a spouse, romantic partner, or child die? | ▩ |
|  | 14 | Have you ever had a serious or life-threatening illness? | **✰** |
|  | 15 | Have you ever received news of a serious injury, life-threatening illness, or unexpected death of someone close to you? | ▩ |
|  | 16 | Have you ever had to engage in combat while in military service in an official or unofficial war zone? | □ |
| Sexual and physical trauma events | 17 | Has anyone ever made you have intercourse or oral or anal sex against your will? | ◇ |
|  | 18 | Has anyone ever touched private parts of your body, or made you touch theirs, under force or threat? | ◇ |
|  | 19 | Other than the two previous incidents mentioned, have there been any other situations in which another person tried to force you to have an unwanted sexual contact? | ◇ |
|  | 20 | Has anyone, including family members or friends, ever attacked you with a gun, knife, or some other weapon? | □ |
|  | 21 | Has anyone, including family members or friends, ever attacked you without a weapon and seriously injured you? | □ |
|  | 22 | Has anyone in your family ever beaten, spanked, or pushed you hard enough to cause injury? | □ |
| OTHER | 23 | Have you ever experienced any other extraordinarily stressful situation or event that is not covered above? |  |
| Key |  | Trauma Category | |
| □ |  | Interpersonal Physical Violence Trauma |  |
| ◇ |  | Sexual Trauma |  |
| ▩ |  | Sudden Grief/Loss Trauma |  |
| **✰** |  | General/Non-Interpersonal Trauma |  |

Three items explicitly probed for the experience of sexual assault (i.e., a sexual encounter made “against your will,” “under force or threat,” or which involves “force… to have an unwanted sexual contact;” items 17, 18, and 19). Sexual abuse has been investigated previously in the literature as a trauma category, such that items similarly capturing non-consensual sexual contact have been aggregated (Bernstein et al., 2003). These THQ items were thus aggregated to form a ‘Sexual Trauma’ category.

Seven items explicitly probed for the experience of threat to physical safety due to interpersonal circumstances (items 1, 2, 4, 16, 20, 21, and 22 broadly probe for experiences of direct robbery, home invasion or frank physical assault). Physical abuse has been investigated previously in the literature, such that items screening for experiences such as being hit hard enough to cause bruising or injury have been aggregated (Bernstein et al., 2003). These THQ items were thus aggregated to form a category concerning ‘Interpersonal Physical Violence Trauma.’ Item 4 (which probes for the experience of a home invasion while present at home) was also included here, as it similarly probed for the experience of threat to physical safety due to interpersonal circumstances.

Four items explicitly probed for experiences of threat to physical safety due to non-interpersonal circumstances (items 5, 6, 7, and 14). Several other items, such as those concerning the experience of “other situations” occasioning serious injury (i.e., items 8, 9, and 10) were also deemed to capture information regarding non-interpersonal trauma. In previous literature investigating non-interpersonal trauma, items screening for the experience of natural disasters, serious personal illness, and serious accidents have similarly been aggregated (Baker et al., 2021; Yoo et al., 2018). Items 5, 6, 7, 8, 9, 10, and 14 alone were therefore aggregated to form a General Trauma category.

Three items explicitly probed for the experience of sudden loss (items 12, 13, and 15). Suddenly losing a close one, for example, due to suicide, homicide, or an accident, has been found to constitute an important risk factor for a variety of internalising disorders, including post-traumatic stress disorder and depression (Guldin et al., 2017). As such, these items were grouped to form a Grief Trauma category.

Item 23 (“Have you ever experienced any other extraordinarily stressful situation or event that is not covered above?”) was deemed too broad for inclusion. Item 11 was excluded because numerous respondents reported exposure to deceased bodies in a university setting, such as cadaver studies for health students. Item 3 (“Has anyone ever attempted to or succeeded in breaking into your home when you were not there?”) was omitted due to its incongruity with the other item categorizations, and a dearth of prior literature supporting its inclusion in any grouping.

Items were grouped according to the principles outlined above. Participants who recorded *‘no’* to ever experiencing an item were given an associated frequency score of 0. First ages for each of the trauma groupings were calculated by finding the minimum age recorded (if experienced and supplied) between the relevant items for that item grouping. A total score was also calculated to reflect total trauma exposure (this did not include items not included in the subgrouping procedure).

**Section S1.5 – Socioeconomic Measure Validation**

SES has been previously observed to covary with trauma exposure (Walsh et al., 2019). In the present analysis Family SES was significantly correlated with total trauma exposure as measured by the Trauma History Questionnaire (THQ); ρ = –.11, p = .007), whereas Participant SES was not (ρ< .01, p = .276). Both Family and Participant SES correlated with adult intelligence, as measured via the Wechsler Abbreviated Scale of Intelligence - Second Edition Composite score (WASI-II) (Wechsler, 1999, 2018): Family SES: ρ= .20, p < .001; Participant SES: ρ= .14, p < .001. This replicates prior findings on the association between SES and intelligence (Bradley & Corwyn, 2002; von Stumm & Plomin, 2015). The results therefore indicate composite validity.

**Section S1.6 – MRI Preprocessing**

All MRI data were collected on a Siemens 3T Skyra (Siemens Healthineers, Erlangen, Germany).

*T1 Data Preprocessing*

T1-weighted images were acquired using a magnetization-prepared 2 rapid-acquisition gradient-echoes (MP2RAGE) sequence (TR = 5000ms, TE = 2.98ms, TI1/T12 = 700/2500ms, 1.0mm cubic voxel size). The MP2RAGE sequence involves multiplying two spatially aligned acquired structural images with different inversion times (INV1 and INV2) to form a single T1-weighted image (UNI). While the combination is associated with optimised grey-to-white matter contrast, improving tissue segmentation accuracy (Marques et al., 2010), it also increases noise in non-brain regions. Using the presurfer package, we generated a series of masks (e.g. whole-brain, non-brain regions) from INV2 (Kashyap et al., 2021) (<https://github.com/srikash/presurfer>) to suppress areas particularly affected by noise in UNI. Freesurfer’s recon-all pipeline was then applied for denoising, cortical reconstruction, and segmentation (Dale et al., 1999)– information also incorporated into functional data processing (below).

To extract region-wise grey matter volume estimates from cortical regions, we projected the Schaefer300 atlas to individual space before cortex division into 300 functionally homogenous regions (Schaefer et al., 2018). For subcortical regions, we registered the Freesurfer processed image to MNI152NLin2009cAsym space (Avants et al., 2008) before applying the Melbourne Scale 2 Subcortical Atlas, dividing the subcortex into 32 functionally homogenous regions (Tian et al., 2020).

*Functional Data Preprocessing*

Multi-echo-resting-state-functional magnetic resonance images (ME-rs-fMRI) data were collected using an interleaved sequence (isotropic voxel size = 3.2mm, slices = 40, repetition time (TR) = 0.91s, phase encoding direction = R-L, echo times TE1 = 12.60 ms, TE2 = 29.23 ms, TE3 = 45.86 ms, TE4 = 62.49 ms, flip angle = 56°, bandwidth = 2520 Hz/Px, echo spacing =0.00025, GRAPPA acceleration factor R= 2, Volumes = 767, Multi-band Factor = 4). An opposite phase encoding scheme (L-R) with 10 volumes, but otherwise identical parameters, was also acquired for susceptibility distortion correction.

We executed the preprocessing pipeline recommended by Constable et al. (2025) for ME-rs-fMRI data, which is as follows (see <https://github.com/cicadawing/Single-vs-Multi-Echo-fMRI-Denoising-Strategies>). fMRI images were first minimally preprocessed using the container fMRIPrep 23.1.3 (Esteban et al., 2019) based on Nipype 1.8.6 (Gorgolewski et al., 2011). In brief, workflows first involved the creation of spatial probability maps from T1 data describing the voxel-level tissue-specific probabilities (Zhang et al., 2001), followed by registration to standard space (MNI152NLin2009cAsym) (Avants et al., 2008). Both parameters for head motion correction (via rigid body alignment) (Jenkinson et al., 2002) and subsequent susceptibility distortion correction (with the reverse phase encoded image) (Andersson et al., 2003; Smith et al., 2004) were estimated from the first echo image before application to the remaining echo data. Individual echo time-series were otherwise preprocessed independently. Scans were then registered to T1w space for anatomical alignment using boundary-based registration (Greve & Fischl, 2009), before registration to standard space (MNI152NLin2009cAsym) (Avants et al., 2008). Slice timing correction was not performed due to short TR.

An advantage of ME acquisitions is that, with many measurements per radiofrequency pulse, voxel-wise signal decay (T2*) can be estimated (Posse et al., 1999). This was done by fitting echoes with reliable signal for a given voxel to a monoexponential signal decay model. The calculated T2* map was then used to optimally combine preprocessed BOLD data across echoes following Posse et al. (Posse et al., 1999). This involved the application of a weighted average across echoes to maximise signal dropout to tissue contrast ratios, a step executed using the software *tedana* (DuPre et al., 2021).

The 24-head motion parameters describing head rotation and translation estimated by fMRIPrep were then regressed from the data to mitigate head-motion-related variance using FSL (Smith et al., 2004). To further mitigate the influence of head motion, we then smoothed the data with a 3mm kernel before executing the Independent-Components Analysis Automatic Removal of Motion Artefacts (ICA-AROMA) algorithm (Pruim et al., 2015), which divided the time-series into orthogonal voxel clusters with similar signal signatures (independent components). We did not apply any additional preprocessing at this step and supplied a static reference image created via the median of motion-corrected volumes (estimated by fMRIPrep). Head-motion-related components were regressed from the non-smooth image, having been automatically identified by the algorithm based on robust heuristics explicated in the literature (e.g., high-frequency power, irregular oscillatory patterns, poor localisation to grey matter).

The toolbox Rapidtide was then used to estimate low-frequency oscillations (LFO) via the Regressor Interpolation at Progressive Time Delays (RIPTiDe) algorithm (Korponay et al., 2024; Tong et al., 2019) (<https://github.com/bbfrederick/rapidtide>). Probes were fit to each voxel to estimate blood-arrival time; nuisance information regressed from the voxel time-series. This was performed as an alternative to the more commonly practiced global signal regression – but which demonstrates superior denoising efficacy and true signal retention. This was a replication of the procedure applied in our prior work (Constable, Tiego, Pavlovich, Sangchooli, et al., 2025).

Finally, gray matter probability maps derived from fMRIPrep and the preprocessed BOLD images were registered to MNI152NLin2009cAsym space (Avants et al., 2008). These were multiplied together to limit partial volume effects. Data were then bandpass filtered (0.008 < f < .08 Hz).

We similarly parcellated fMRI data using the combined Schaefer400 and Melbourne Scale 2 Subcortical Atlas, dividing the cortex into 400 and the subcortex into 32 functionally homogenous regions in a single step (Schaefer et al., 2018; Tian et al., 2020). The time-series for each region was extracted and individual-specific FC matrices were calculated using product-moment correlations between regional time courses, as implemented in Nilearn, based on Nipy (Brett et al., 2009; Gorgolewski et al., 2011). These correlation values were then transformed into z-scores using Fisher’s r-to-z transformation (Fisher, 1915).

High head-motion participants were excluded from the data, identified as either mean frame-wise displacement (FD) exceeding 0.30mm, >20% of FD estimates exceeding 0.20mm, any FD estimate exceeding 5 mm, or >50% of data flagged as likely motion contaminated (Parkes et al., 2018). To account for pseudomotion likely present due to the rapid TR associated with multiband sequences, we followed prior work (Fair et al., 2020) and filtered the six rotation and translation head motion parameters estimated by fMRIPrep using a band-stop Butterworth filter [0.31–0.41 Hz]. Summed absolute successive differences were computed from these processed vectors and used to re-estimate FD for each time point (Fair et al., 2020). Timepoints exceeding 0.20mm in FD were flagged as likely motion contaminated. If fewer than 5 timepoints occurred between periods flagged, these originally unmarked volumes were also flagged (Gordon et al., 2014).

**Section S 1.7 – Missing Data Analysis**

Trauma frequencies and age of onset for each trauma type, Family SES, participant age, participant sex, and spectrum psychopathology data were used as input for Little’s Missing Completely at Random (MCAR) Test (Little, 1988), which revealed missing data that were not MCAR; χ2 = 154, *df* = 112, *p* = .005. Most missing data was due to missingness in the Family SES Composite. A binomial general linear model was used to predict missingness in Family SES, which revealed that Participant SES and Age were predictive of missingness in Family SES composite values; Participant SES as predictor - *B*= −.42, *SE* = 0.11, *z* = −3.75, *p* <.001; Age as predictor - *B*=.08, *SE* = .01, *z* =5.09, *p* <.001. The data were hence deemed Missing at Random (MAR), qualifying the use of imputation methods. A dataset containing variables for analysis (trauma frequency data, age of onset to the trauma types of interest, participant age, participant sex, Family SES, and spectrum psychopathology data) and Participant SES information was used for multiple imputations using the expectation-maximization with bootstrapping approach (EMB), executed via the *Amelia* package (Honaker et al., 2011)*.* This approach first estimates missing values using Expectation-Maximisation then applies bootstrapping to create multiple datasets to reflect uncertainty with estimation (Rubin, 2004). While Participant SES was not included in the overall SEM models, it was included during imputation for auxiliary information (information not considered during the main analysis). The package *semTools* and *lavaan.mi* (Jorgensen et al., 2022) was used to fit the structural-equation models estimated in *lavaan* (Rosseel, 2012) to each imputed dataset before subsequently pooling according to Rubin’s rules, accounting for within and between analysis variation (Rubin, 2004). Twenty-five imputed datasets to account for missingness were necessary to stabilise pooled estimates in the present study.

**Section S1.8 Permutation Significance Testing of PCs of Neurobiological Data**

PCAs were conducted for grey matter and functional-connectivity data separately. Individuals were excluded from the PCA only if they failed quality metrics for the relevant MRI type – not because of survey data. This ensured that dimensionality was estimated from the full-most dataset. The statistical significance of *n* PCs was then assessed using permutation testing with 10,000 repeats, *n* denoting the number of columns in the data matrix (Vitale et al., 2017). This involved creating a null distribution via concurrently randomly shuffling cells within each column vector, breaking the covariance structure of the overall data matrix while preserving original data distributions. Statistics for the original PCs were compared to those extracted from permuted datasets to determine if they represented meaningful structure or noise. To assess the contributions of loadings per *significant* PC, we also permuted each column in the data matrix 10,000 times while keeping all other column vectors fixed; originally observed loadings and those derived from the null distribution were then compared to assess whether each loading meaningful contributed to each PC (Linting et al., 2011). Originally significant PCs with < 3 significant loadings were dropped from further analyses due to under-identification (Kline, 2023). PC stability was also assessed using bootstrapped resampling; stability was determined if individual-level component scores and component loadings were similar over iterations (both determined with Pearson’s *r* > .95 or Tucker Congruences > .95) (Lorenzo-Seva & ten Berge, 2006). These operations were performed using the *syndRomics* package (Torres-Espín et al., 2021).

The first 22 PCs for the T1 data were significant, though only the first 20 had at least 3 significant loadings. These 20 components explained 60.17% of the variance in the initial data (Figure 2) and were replicable across bootstrap samples (Figure 3). The first 25 PCs for the fMRI data were significant, but only the first 22 had at least 3 significant loadings. These 22 PCs explained 67.95% of the variance in the data (Figure 4) and were replicable across bootstrap samples (Figure 5). For the FC sensitivity analyses, 98 significant PCs were retained (explaining 60.98% of the variance). Further refinement of this number was not explored due to computation costs. See Figure 6.

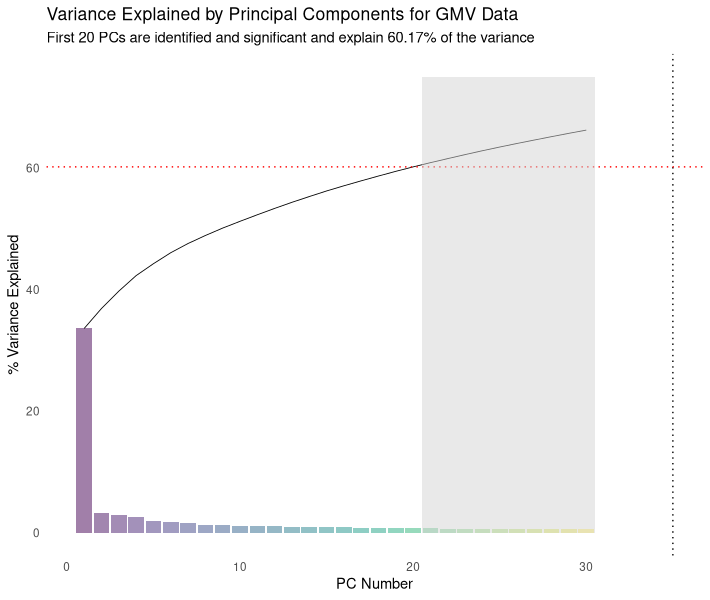

Figure 2. The percentage of variance explained by each principal component (PC) for the GMV data are shown in bars; cumulative variance is represented as a black line. PCA was conducted extracting the maximal number of PCs; only the first 30 are displayed here for convenience. The grey block indicates where the PCs begin to lose significance or have fewer than three significant loadings.

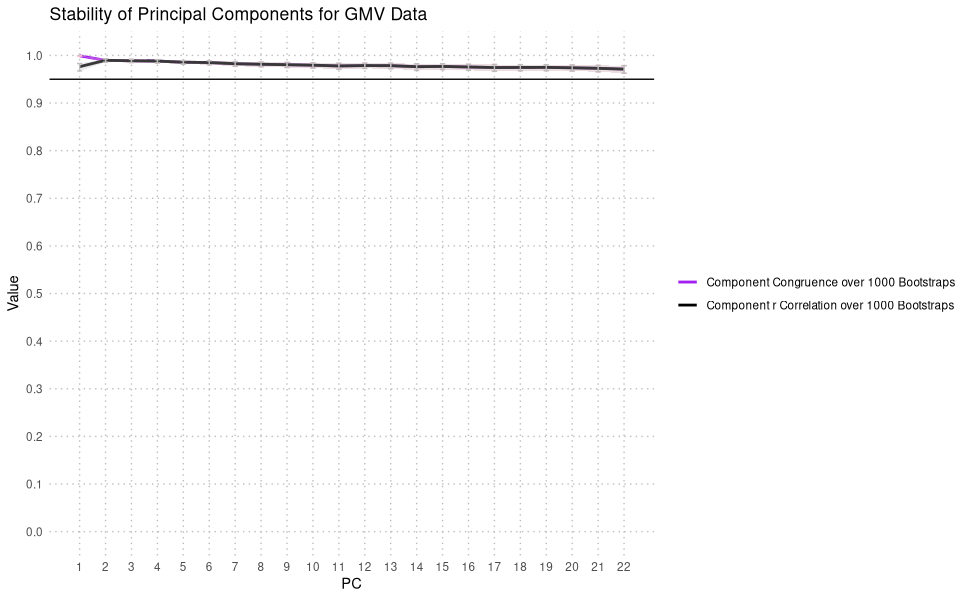

Figure 3. Component Congruence (factor loading similarities) and r (individual PC estimate) stability over 1000 bootstrap samples for the significant T1 Data PCs. All PCs appeared stable over bootstrap samples (both statistics > .95), indicating that the PCs were extracting the same information across iterations (Lorenzo-Seva & ten Berge, 2006).

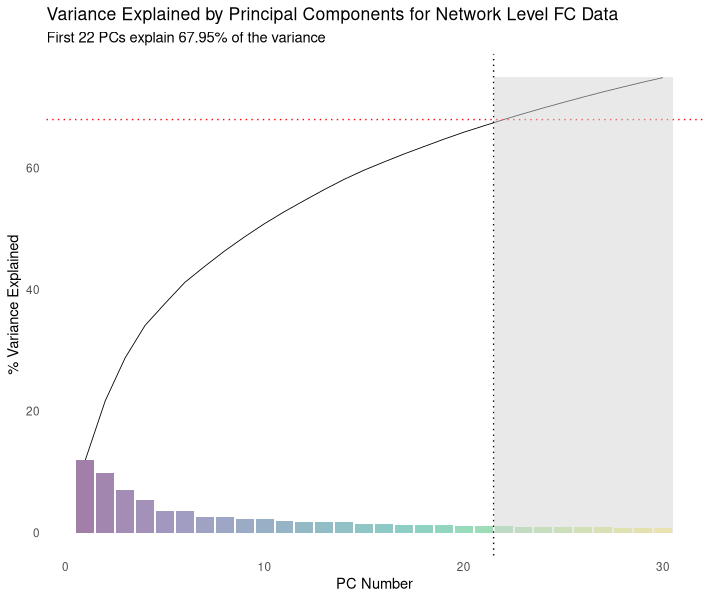

Figure 4. The percentage of variance explained by each principal component (PC) for the fMRI data are shown in bars; cumulative variance is represented as a black line. PCA was conducted extracting the maximal number of PCs; only PCs 1-30 are displayed here for convenience. The grey block indicates where the PCs begin to lose significance or have fewer than three significant loadings.

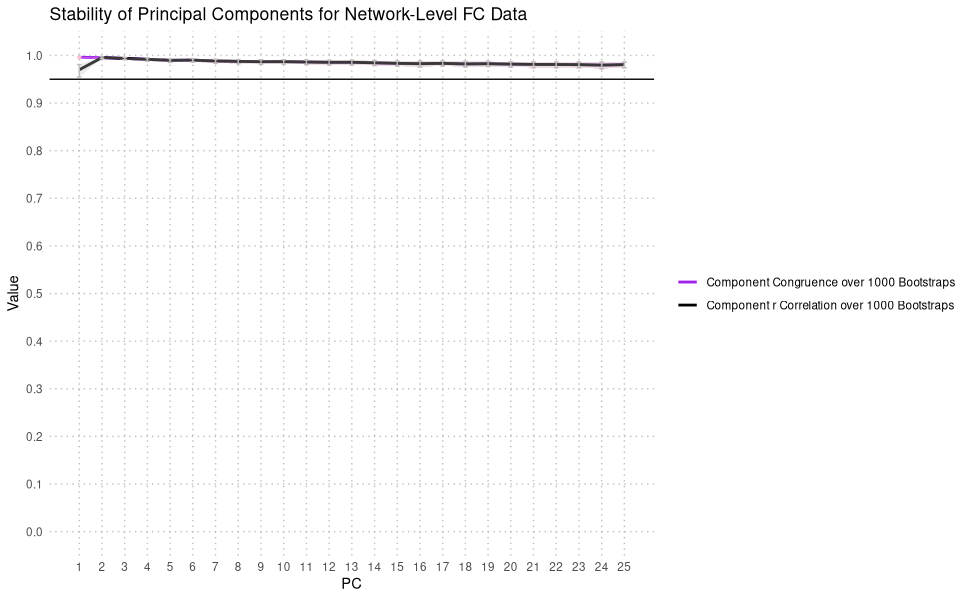

Figure 5. Component Congruence (factor loading similarities) and r (individual PC estimate) stability over 1000 bootstrap samples for the significant fMRI Data PCs. All PCs appeared stable over bootstrap samples (both statistics > .95), indicating that the PCs were extracting the same information across iterations (Lorenzo-Seva & ten Berge, 2006).

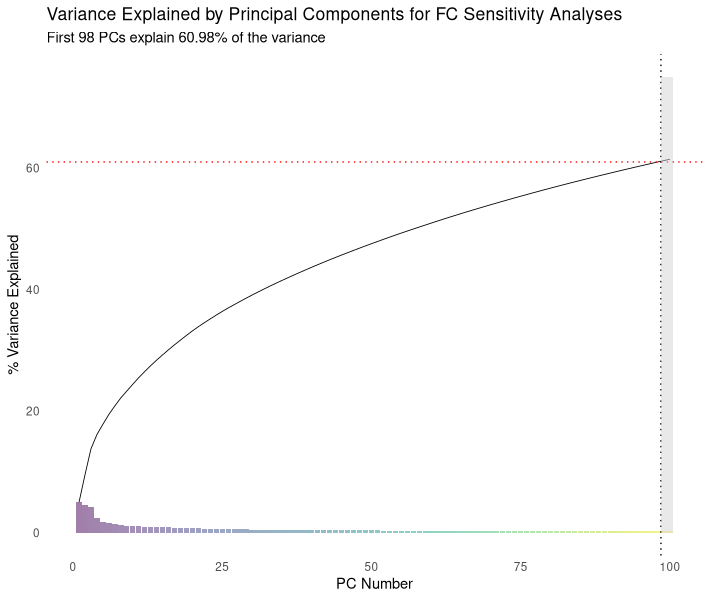

Figure 6. The percentage of variance explained by each principal component (PC) for the fMRI data parcellated with Schaefer100 + Tian S2 parcellation (sensitivity analyses) are shown in bars; cumulative variance is represented as a black line. PCA was conducted extracting the maximal number of PCs; only PCs 1-100 are displayed here for convenience. The grey block indicates where the PCs begin to lose significance.

**Section S1.9 Overall Model *df*, RMSEA, and Significance**

*Table 1.* Model degree of freedom.

| Modality | Psychopathological Outcome Variable | Sexual Trauma *df* | Interpersonal Physical Violence Trauma *df* |
| --- | --- | --- | --- |
| fMRI-Based | Lifetime PTSD | 15 | 21 |
|  | Antisocial Schizotypy | 15 | 21 |
|  | Negative Affectivity | 15 | 21 |
| GMV-Based | Lifetime PTSD | 18 (15 for PC1) | 21 |
|  | Antisocial Schizotypy | 18 (15 for PC1) | 21 |
|  | Negative Affectivity | 18 (15 for PC1) | 21 |

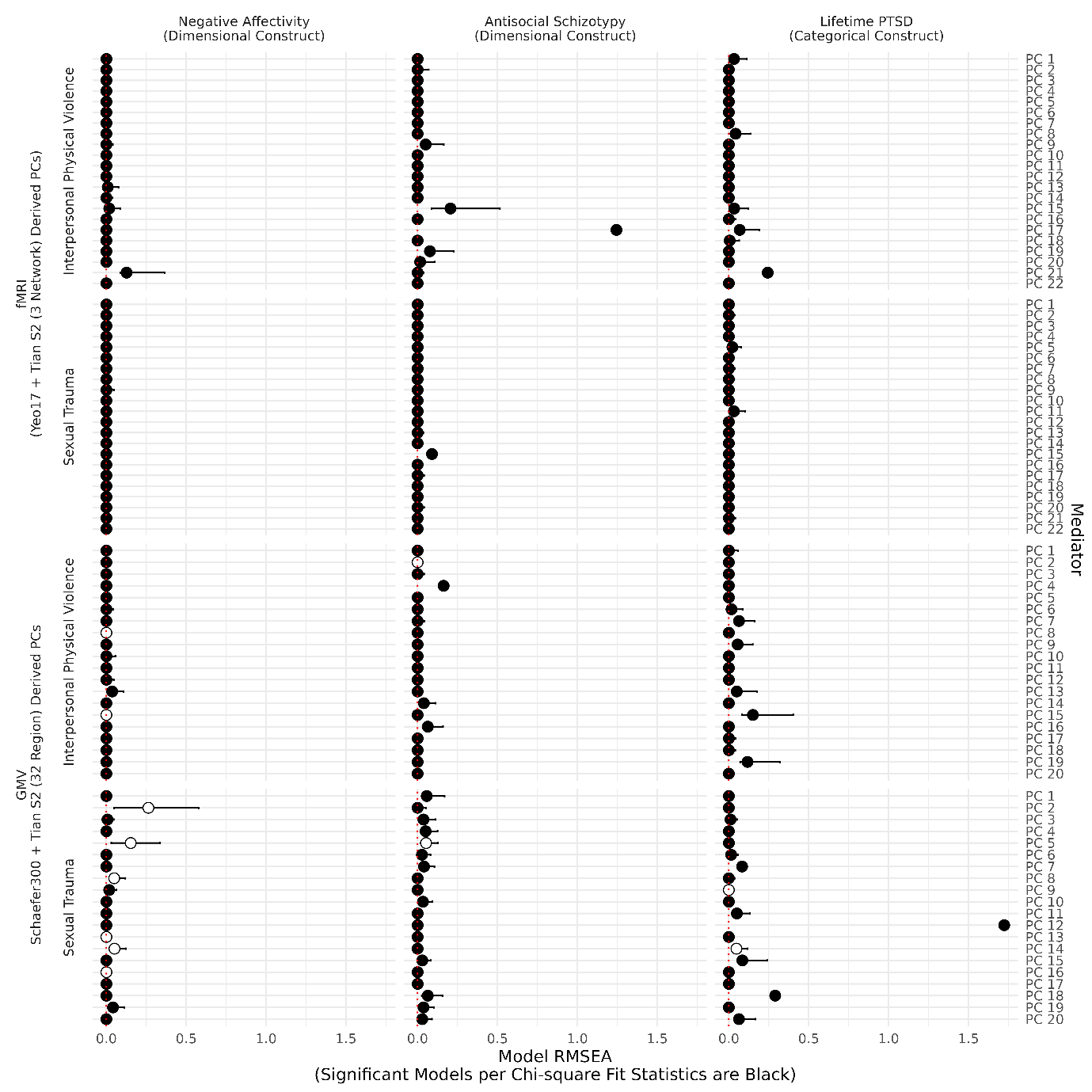

*Figure 2.* RMSEA values for each mediation model. Mediators are shown on the right *y-*axis and have been stratified by MRI modality and trauma type (left *y-*axis); overall psychopathology targets are presented in columns. Bars indicate the lower and upper bounds of RMSEA estimates. Black points denote models with acceptable fit based on chi-square tests (i.e., *p ≥* .05); white points indicate poor fit (i.e., *p <* .05). Simulation work indicates that small-sample RMSEA estimation rates stabilise at approximately *n* > 200 and *df* > 10; thus, the present models were adequately powered (*n* > *200;* *df ≥* 15) (Kenny et al., 2015; Shi et al., 2022).

**Section S1.10 Non-robust evidence of Mediation**

The following passage describes the weak evidence for Mediation for Interpersonal Physical Violence predicting Lifetime PTSD.

For fMRI PC10, overall tests indicated that the strength of mediation varied according to age of onset (*b* = –4.18e–05, CI 97.5%: [-8.7e–04, -4.0e–05]). Follow-up analyses provided borderline evidence (just non-inclusive of zero) that any mediation present was exclusive to individuals first exposed at age 25 (*b* = –0.0005, CI 97.5%: [–0.003, –4.74e–05]).

A similar pattern was observed for fMRI PC28 and fMRI PC 7 in sensitivity analyses using Schaefer100 + Tian S2-derived PCs at the two most liberal thresholds (omnibus *b* = –6.37e–05, CI 97.5%: [–1.41e–04, –4.18e–07]; omnibus *b* = –2.90e–05, CI 97.5%: [–5.90e–05, –7.40e–06], CI 99%: [–6.36e–05, –5.36e–06], respectively). We also observed mediation via fMRI PC 26 dependent on sex (omnibus *b* = –8.59e–04, CI 97.5%: [–2.13e–03, –5.80e–05]) in the prediction of Lifetime PTSD, here (though, not in the main analyses, indicating instability over brain parcellation strategies). However, no significant indirect effects were identified beyond the most liberal CIs investigated (97.5%) when stratifying by specific ages of onset and sex at follow-up. As such, a lack of robust mediation effects does not appear to be a consequent of brain parcellation strategy.

For GMV PC13, we observed weak evidence that the strength of the indirect effect of trauma dose on Lifetime PTSD via GMV PC13 varied by survivor sex (omnibus *b* = 8.34e–05, CI 97.5%: [1.83e–04, 1.50e–05]; CI 99.0%: [2.04e–04, 7.88e–06]). Evidence at follow-up indicated that any mediation present was exclusively for female survivors (*b* = –0.0008, CI 97.5%: [–.002, –6.19–05].).

To further assess the robustness of the already weak evidence for mediation via GMV PC13 described above, we re-estimated *standardised* effects at a 97.5% confidence interval (i.e., standardized = T). The magnitude of effect for the index of moderated mediation was negligible (*β* = .009, CI 97.5%: [.002, .020]) (Cohen, 2013), and no support for an indirect effect among females was observed (*β* = -.001, CI 97.5%: [-.071, .074]). We note that robust effects in path analyses should show convergence between standardised and unstandardized estimates (i.e., invariance to reasonable estimation strategies, including for scaling).
