## Supplementary 4 for "The effects of trauma on neurobiological and psychopathological phenotypes in a transdiagnostic community sample"

Representation for Principal Component 1

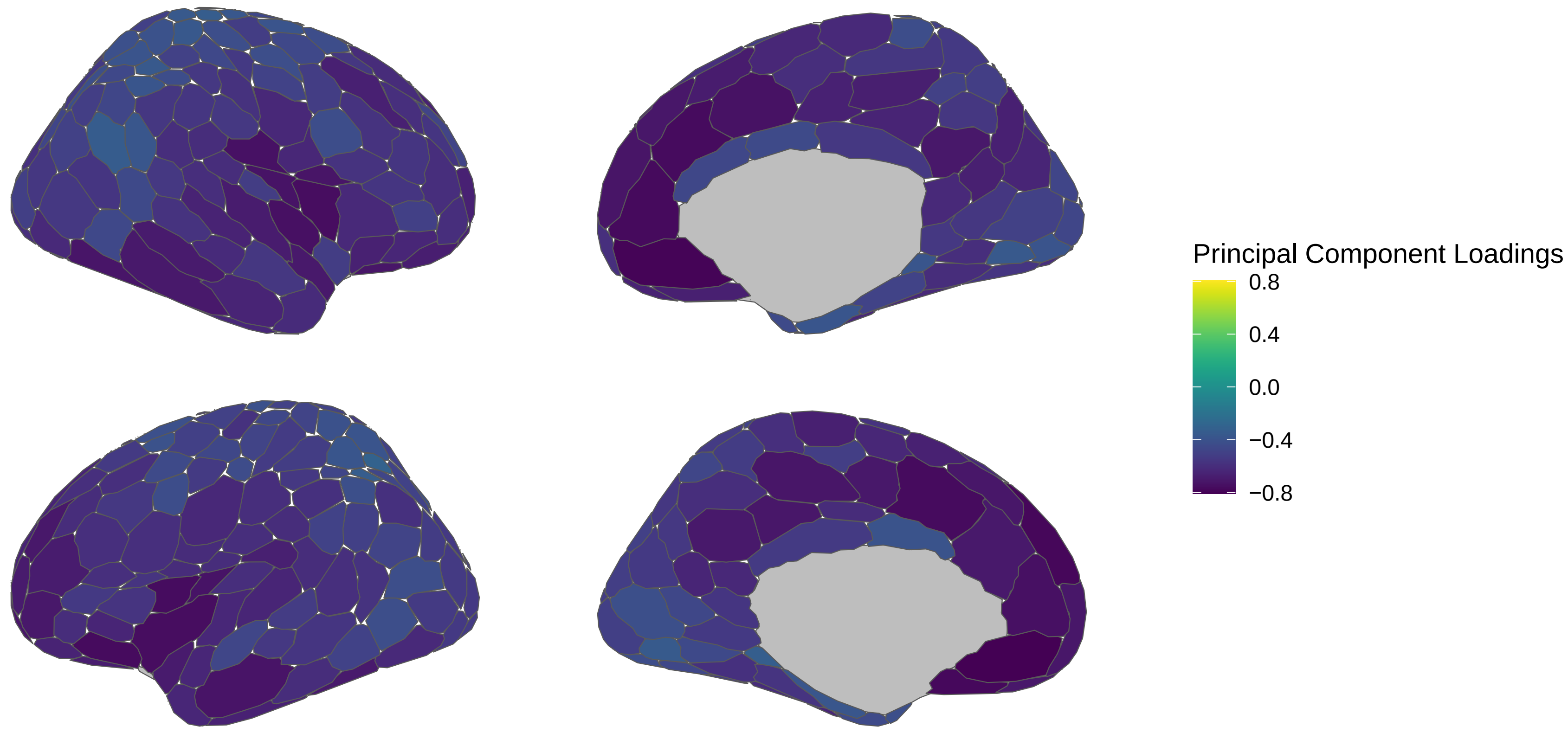


LH Subcortical Loadings

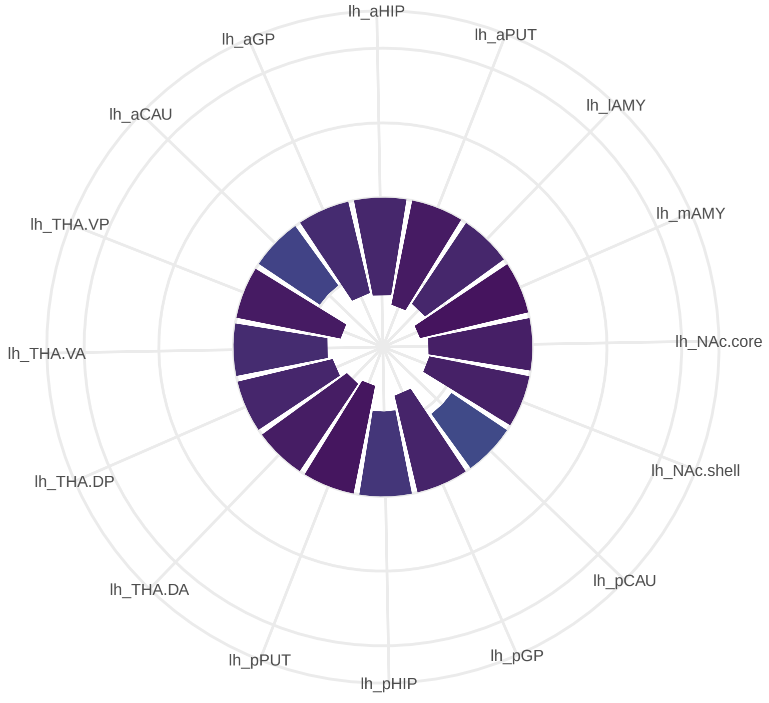


RH Subcortical Loadings

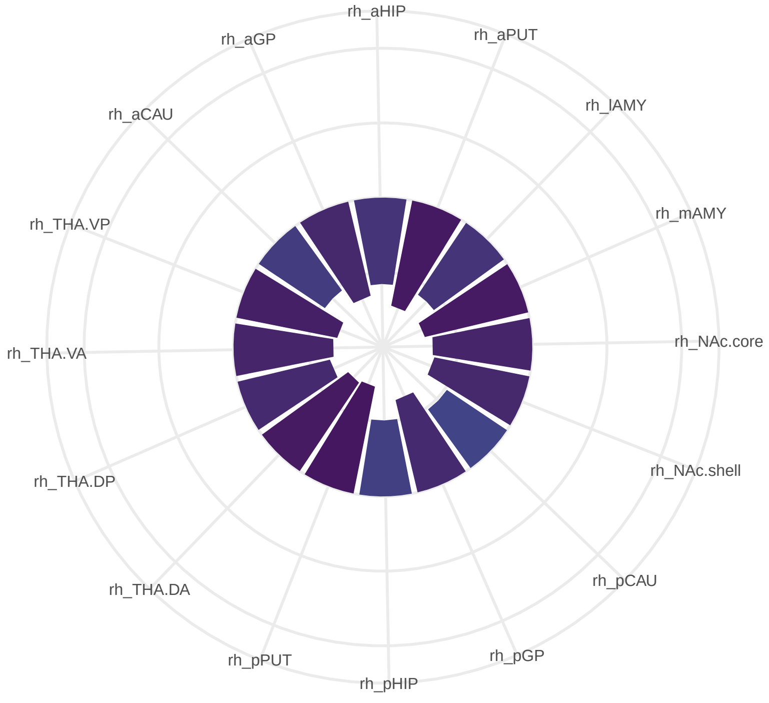

### Representation for Principal Component 2

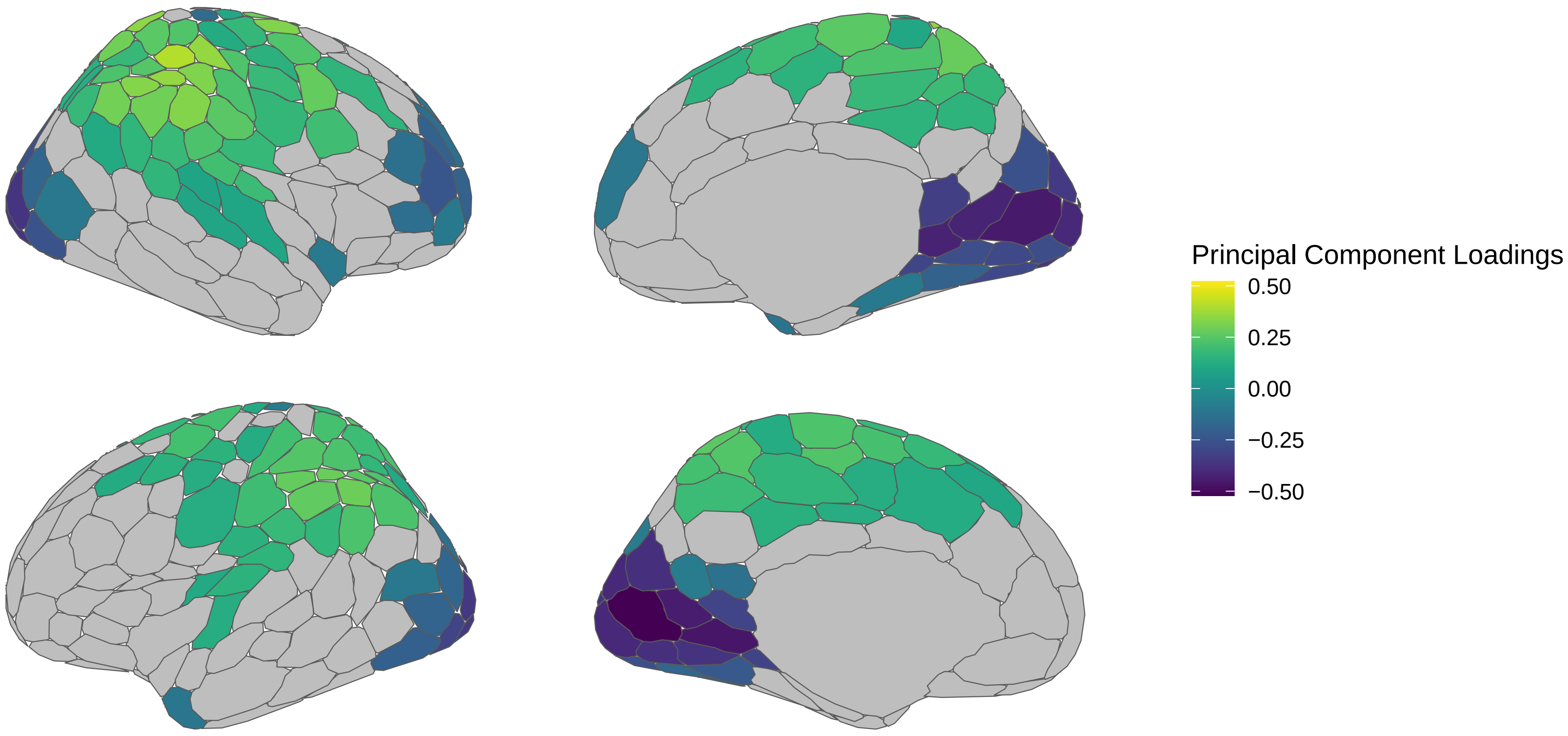


### LH Subcortical Loadings

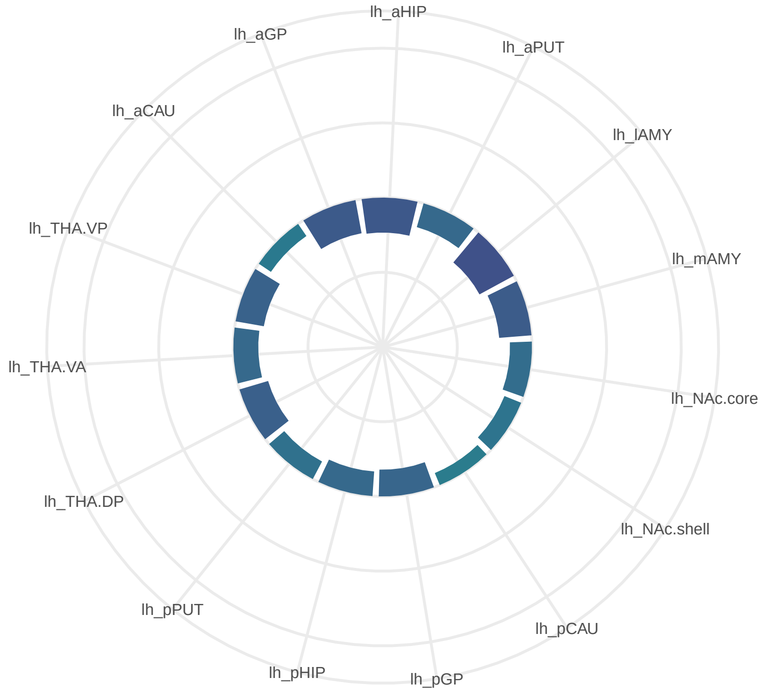


### RH Subcortical Loadings

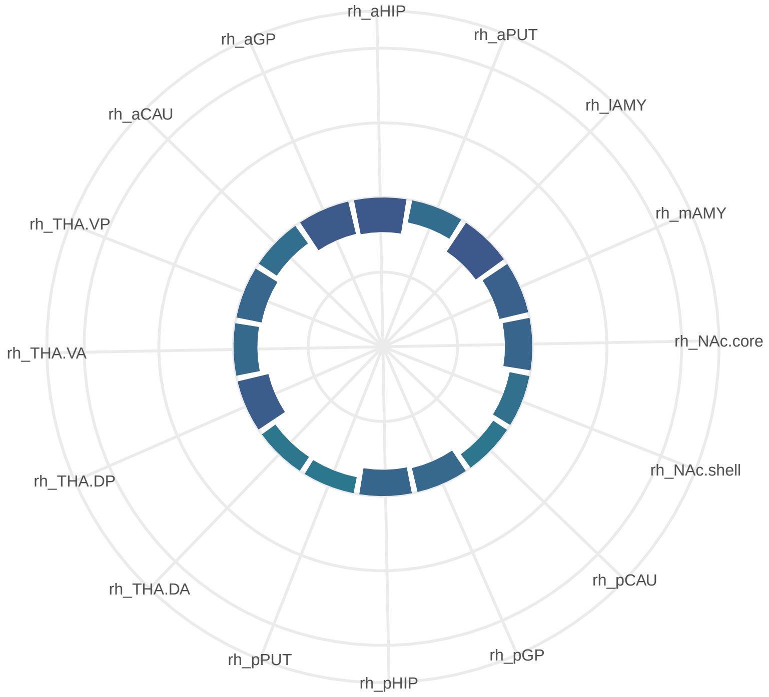

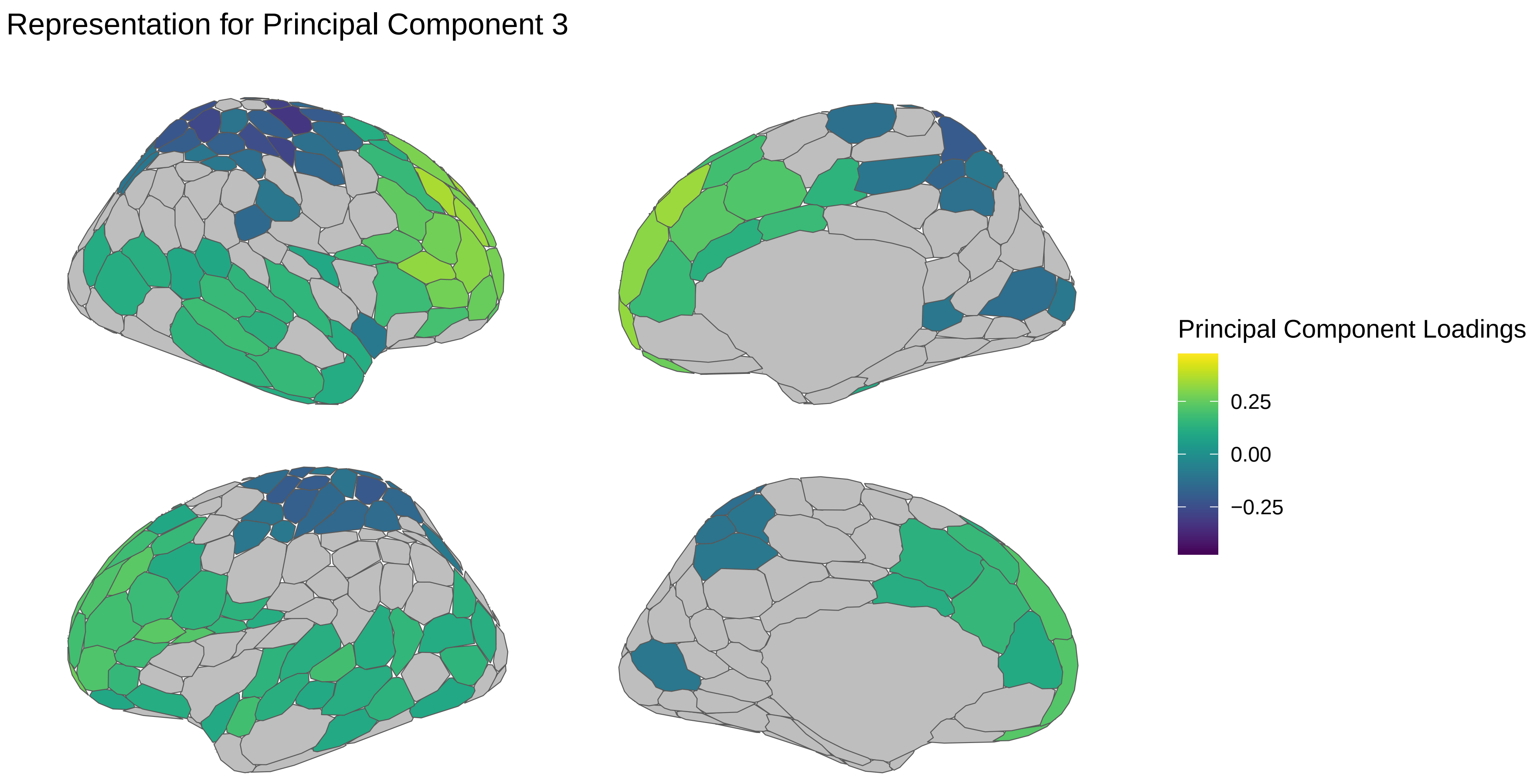

Representation for Principal Component 3
Principal Component Loadings
0.25
0.00
−0.25


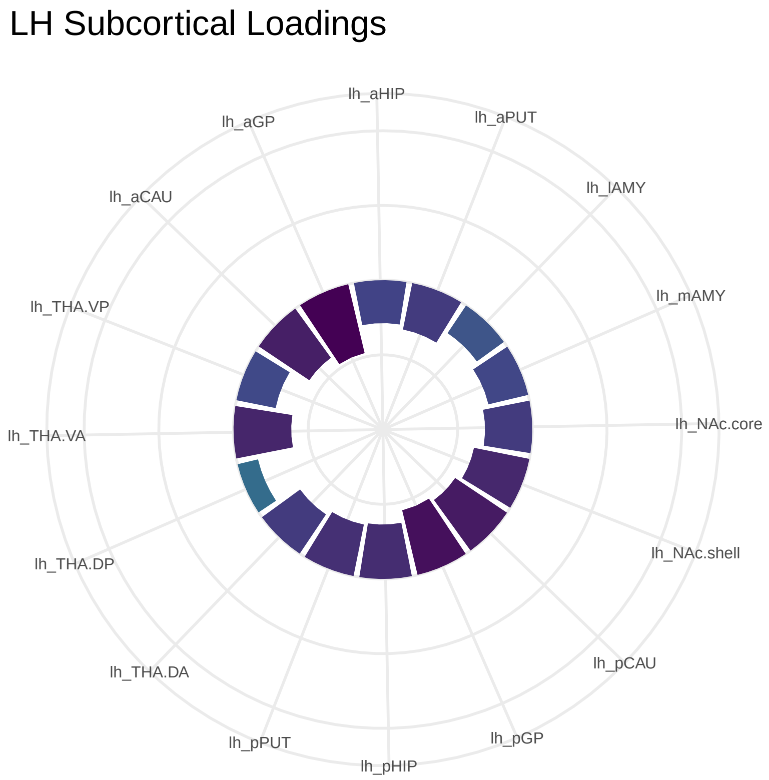

LH Subcortical Loadings
lh_aHIP
lh_aPUT
lh_lAMY
lh_mAMY
lh_NAc.core
lh_NAc.shell
lh_pCAU
lh_pGP
lh_pHIP
lh_pPUT
lh_THA.DA
lh_THA.DP
lh_THA.VA
lh_THA.VP
lh_aCAU
lh_aGP


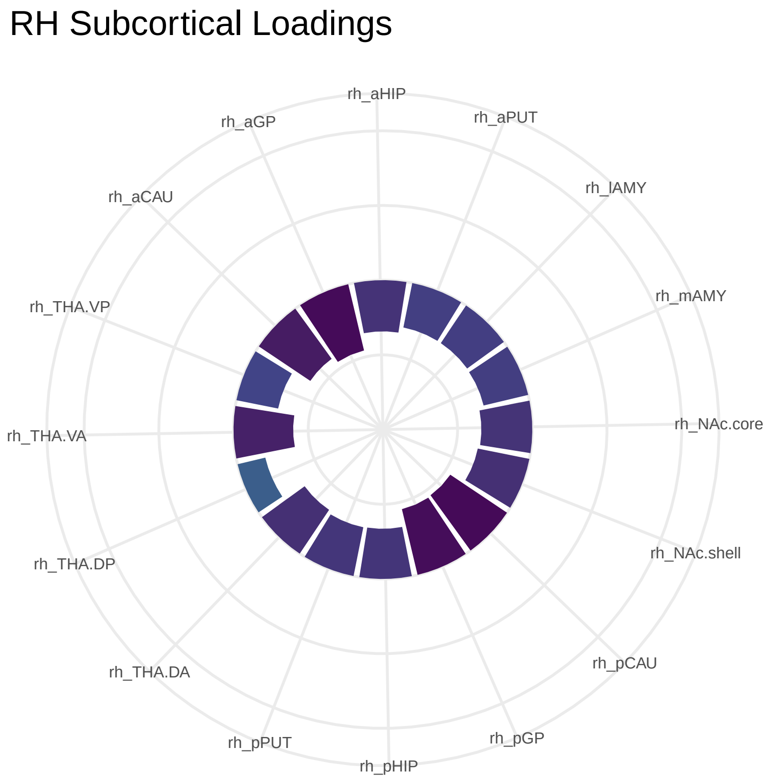

RH Subcortical Loadings
rh_aHIP
rh_aPUT
rh_lAMY
rh_mAMY
rh_NAc.core
rh_NAc.shell
rh_pCAU
rh_pGP
rh_pHIP
rh_pPUT
rh_THA.DA
rh_THA.DP
rh_THA.VA
rh_THA.VP
rh_aCAU
rh_aGP

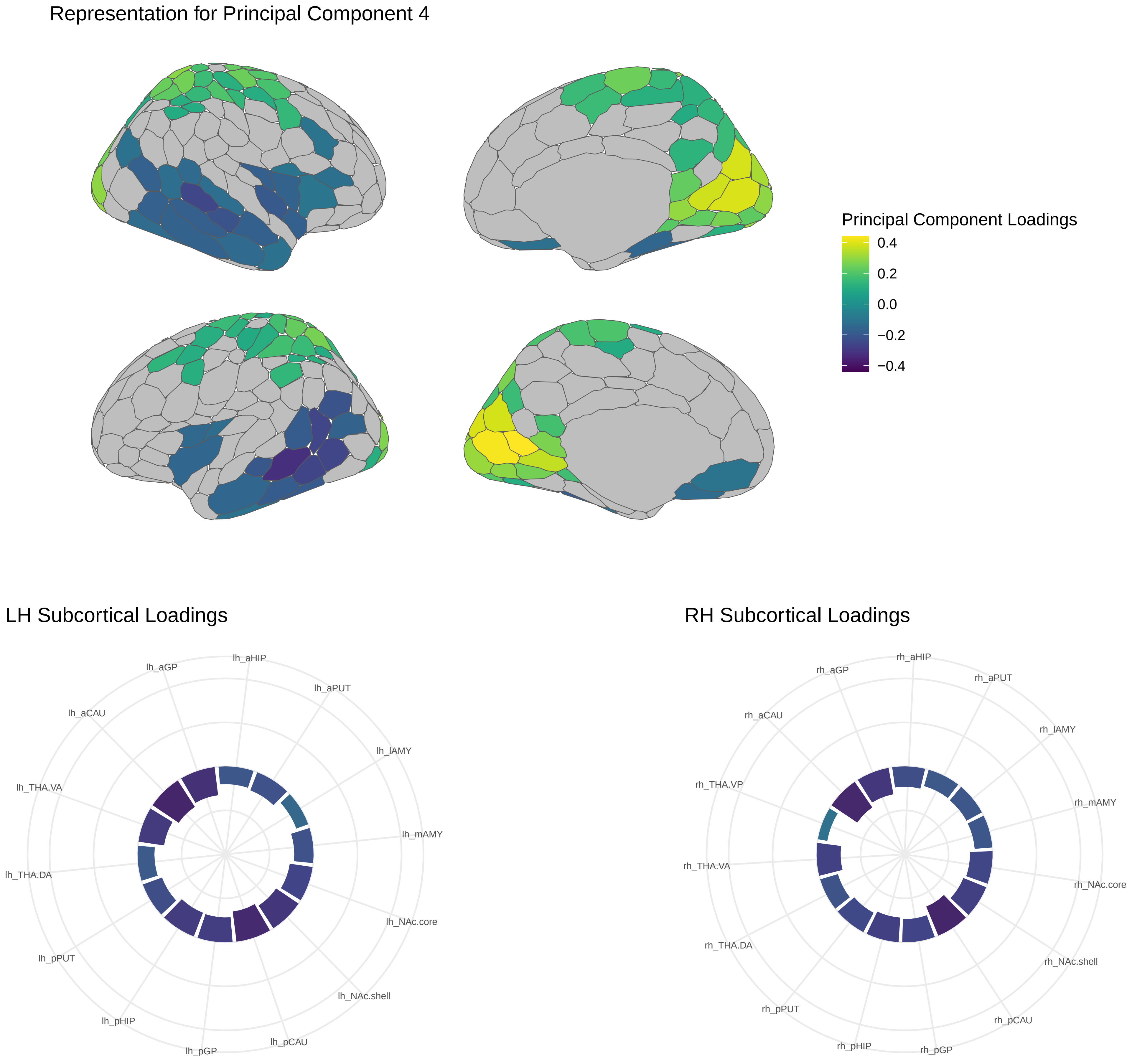
Representation for Principal Component 4
Principal Component Loadings
0.4
0.2
0.0
−0.2
−0.4
LH Subcortical Loadings
lh_aHIP
lh_aGP
lh_aPUT
lh_aCAU
lh_lAMY
lh_THA.VA
lh_mAMY
lh_THA.DA
lh_NAc.core
lh_pPUT
lh_NAc.shell
lh_pHIP
lh_pCAU
lh_pGP
RH Subcortical Loadings
rh_aHIP
rh_aGP
rh_aPUT
rh_aCAU
rh_lAMY
rh_THA.VP
rh_mAMY
rh_THA.VA
rh_NAc.core
rh_THA.DA
rh_NAc.shell
rh_pPUT
rh_pCAU
rh_pHIP
rh_pGP

Representation for Principal Component 5

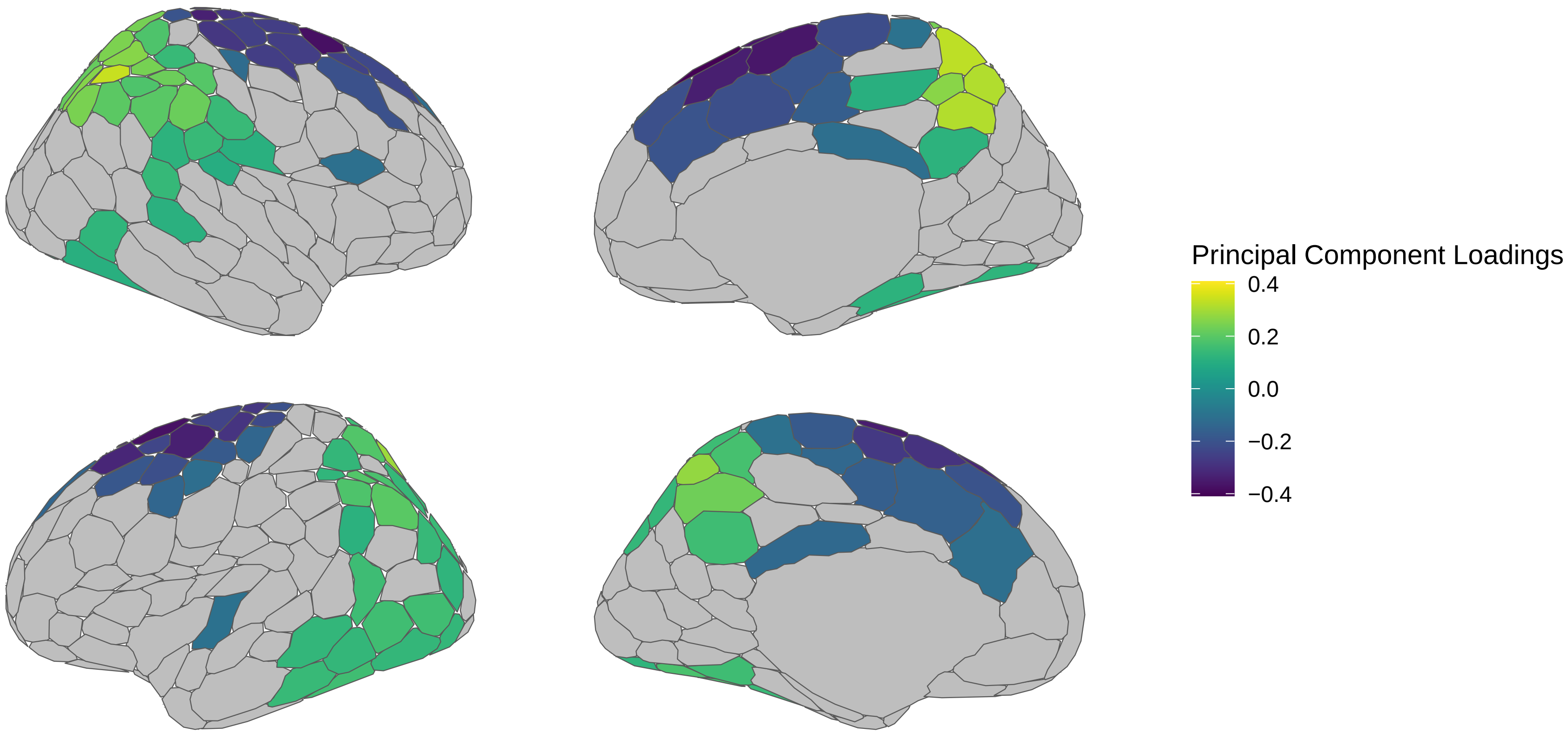


LH Subcortical Loadings



RH Subcortical Loadings

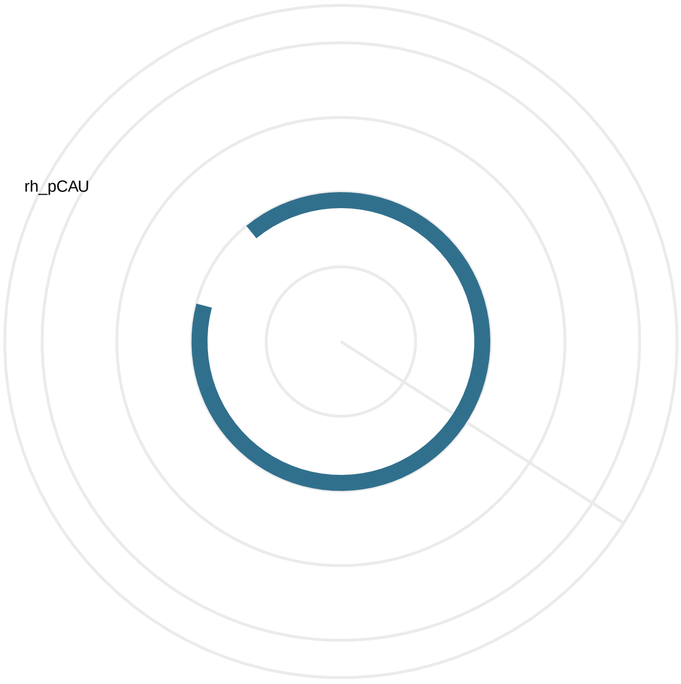

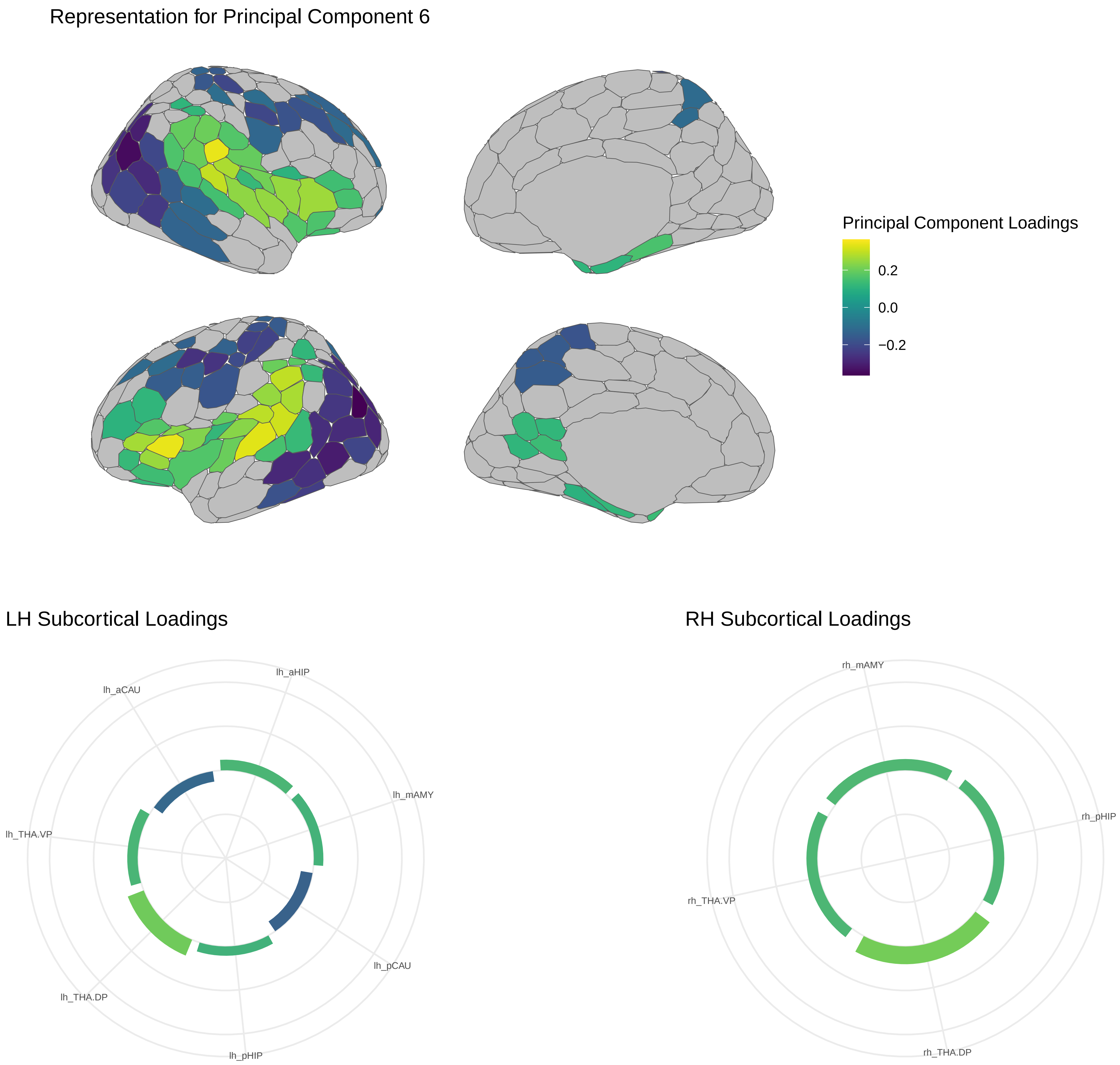
Representation for Principal Component 6
Principal Component Loadings
0.2
0.0
−0.2
LH Subcortical Loadings
lh_aHIP
lh_aCAU
lh_mAMY
lh_THA.VP
lh_pCAU
lh_THA.DP
lh_pHIP
RH Subcortical Loadings
rh_mAMY
rh_pHIP
rh_THA.VP
rh_THA.DP

### Representation for Principal Component 7

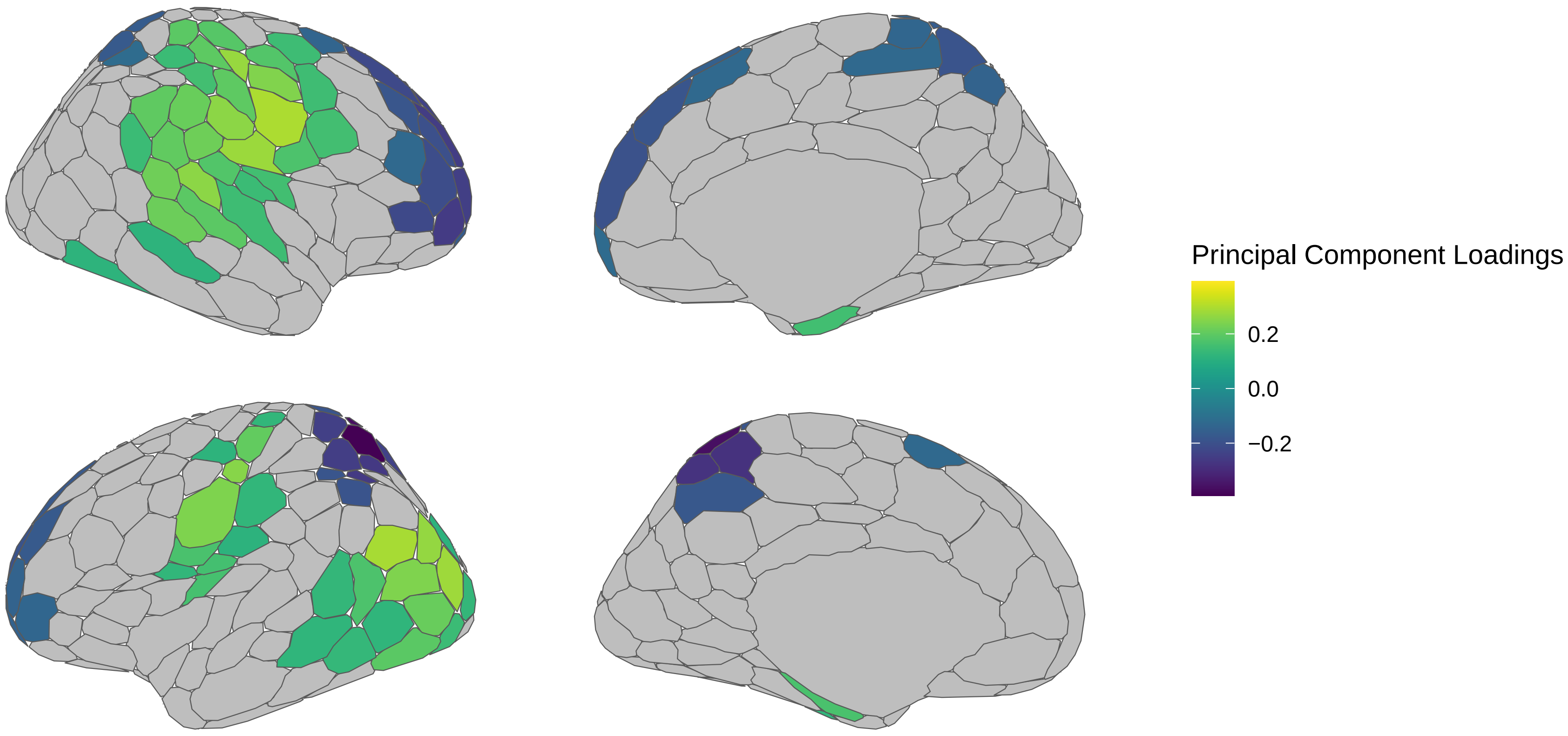


### LH Subcortical Loadings

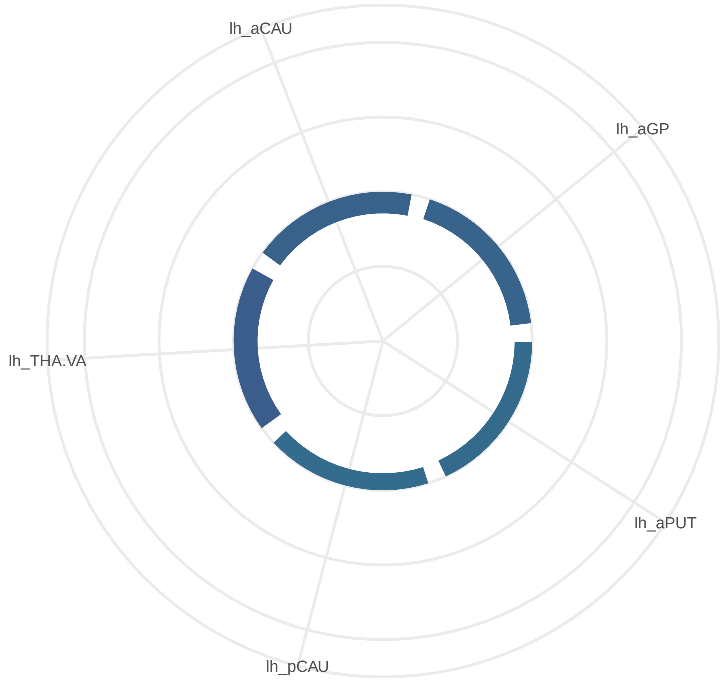


### RH Subcortical Loadings

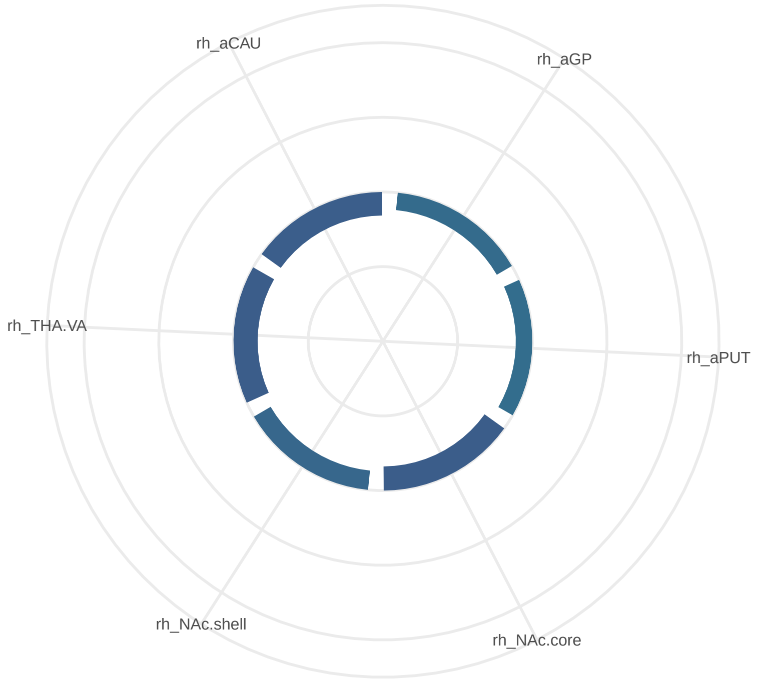

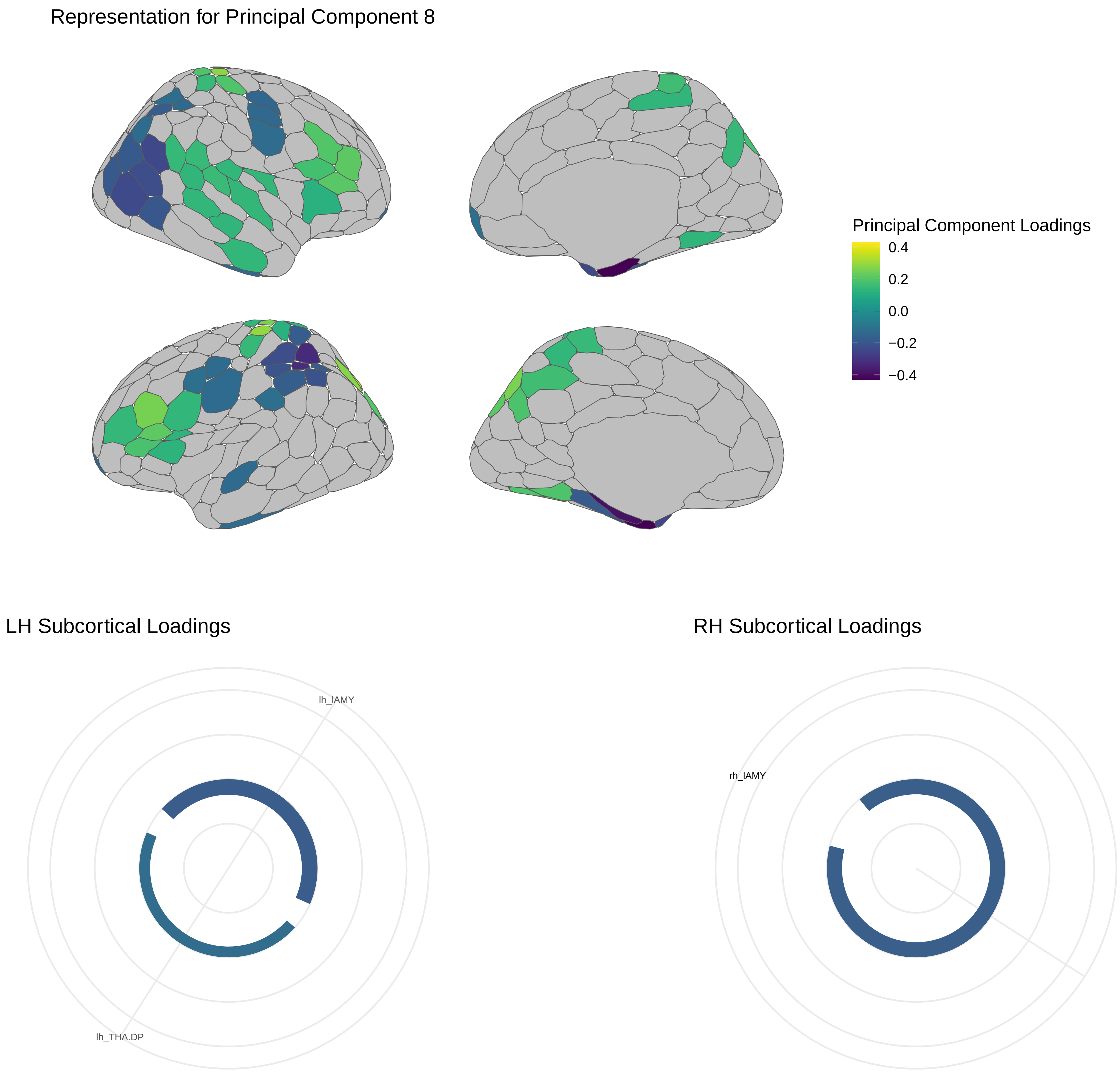
Representation for Principal Component 8
Principal Component Loadings
0.4
0.2
0.0
−0.2
−0.4
LH Subcortical Loadings
lh_lAMY
lh_THA.DP
RH Subcortical Loadings
rh_lAMY

Representation for Principal Component 9

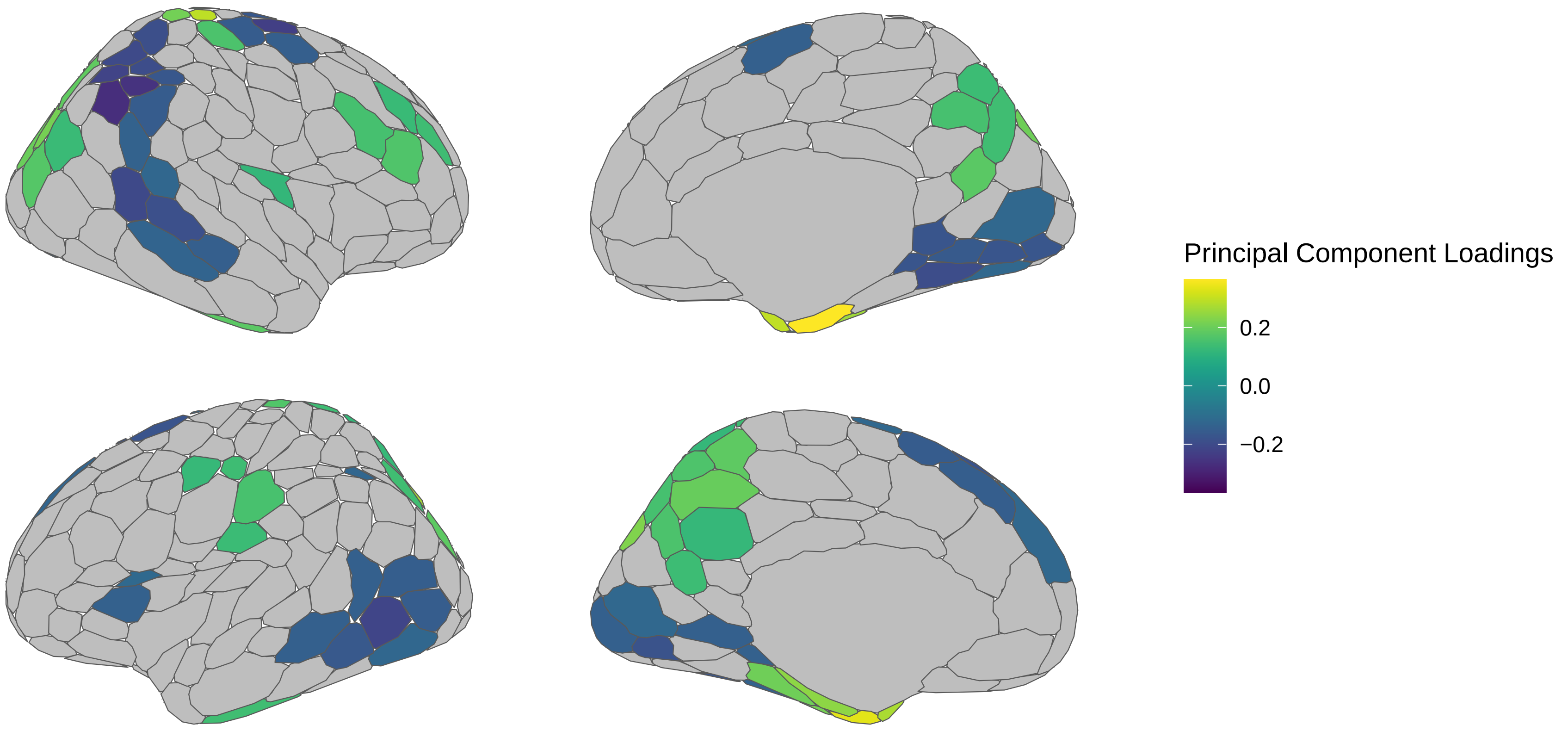


LH Subcortical Loadings

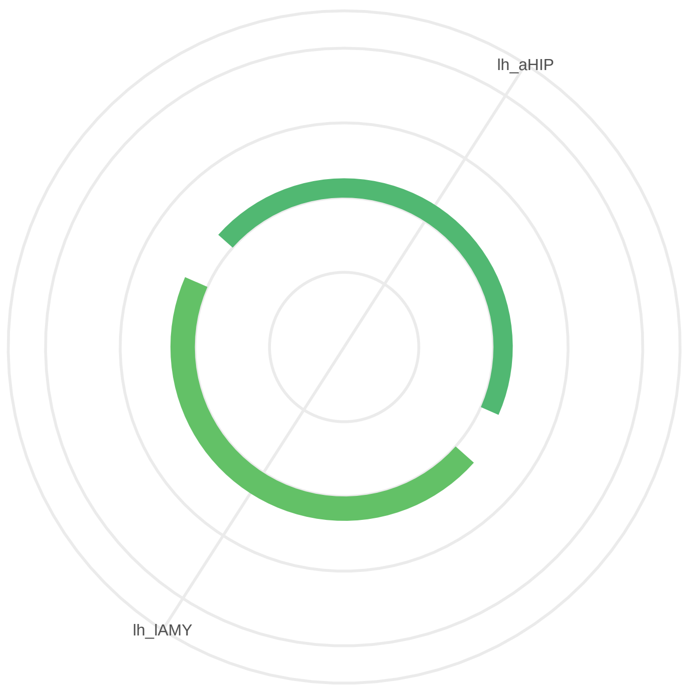


RH Subcortical Loadings

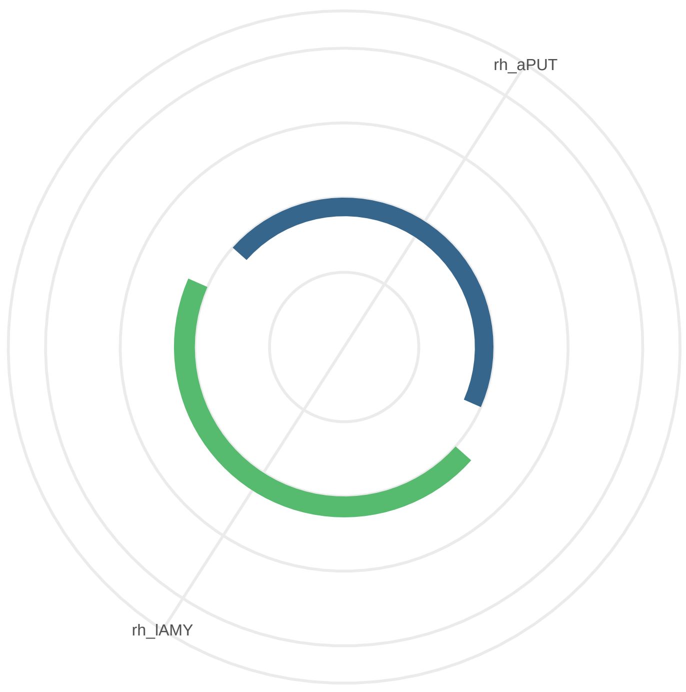

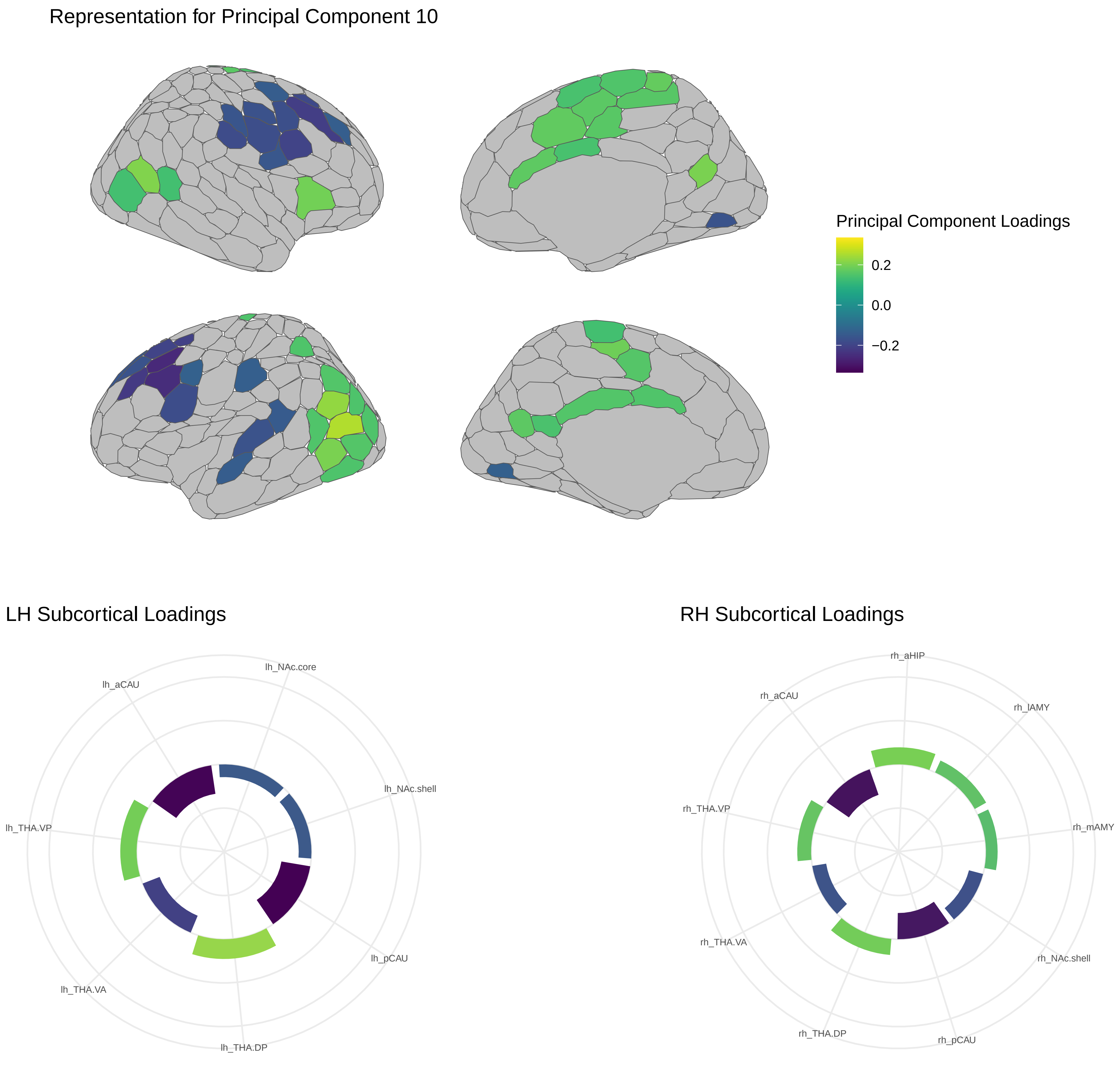
Representation for Principal Component 10
Principal Component Loadings
0.2
0.0
−0.2
LH Subcortical Loadings
lh_NAc.core
lh_aCAU
lh_NAc.shell
lh_THA.VP
lh_pCAU
lh_THA.VA
lh_THA.DP
RH Subcortical Loadings
rh_aHIP
rh_aCAU
rh_lAMY
rh_THA.VP
rh_mAMY
rh_THA.VA
rh_NAc.shell
rh_THA.DP
rh_pCAU

Representation for Principal Component 11

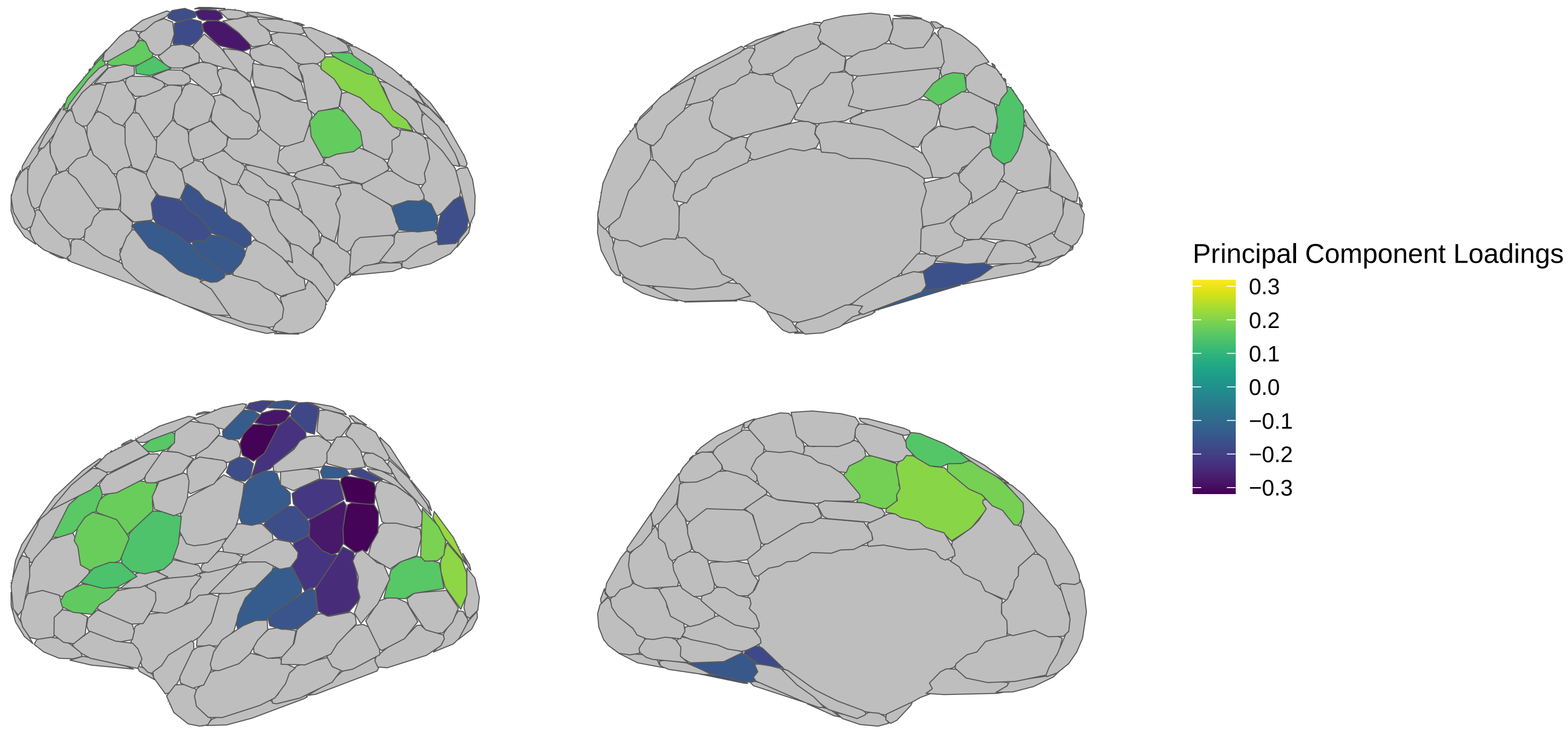


LH Subcortical Loadings



RH Subcortical Loadings

### Representation for Principal Component 12

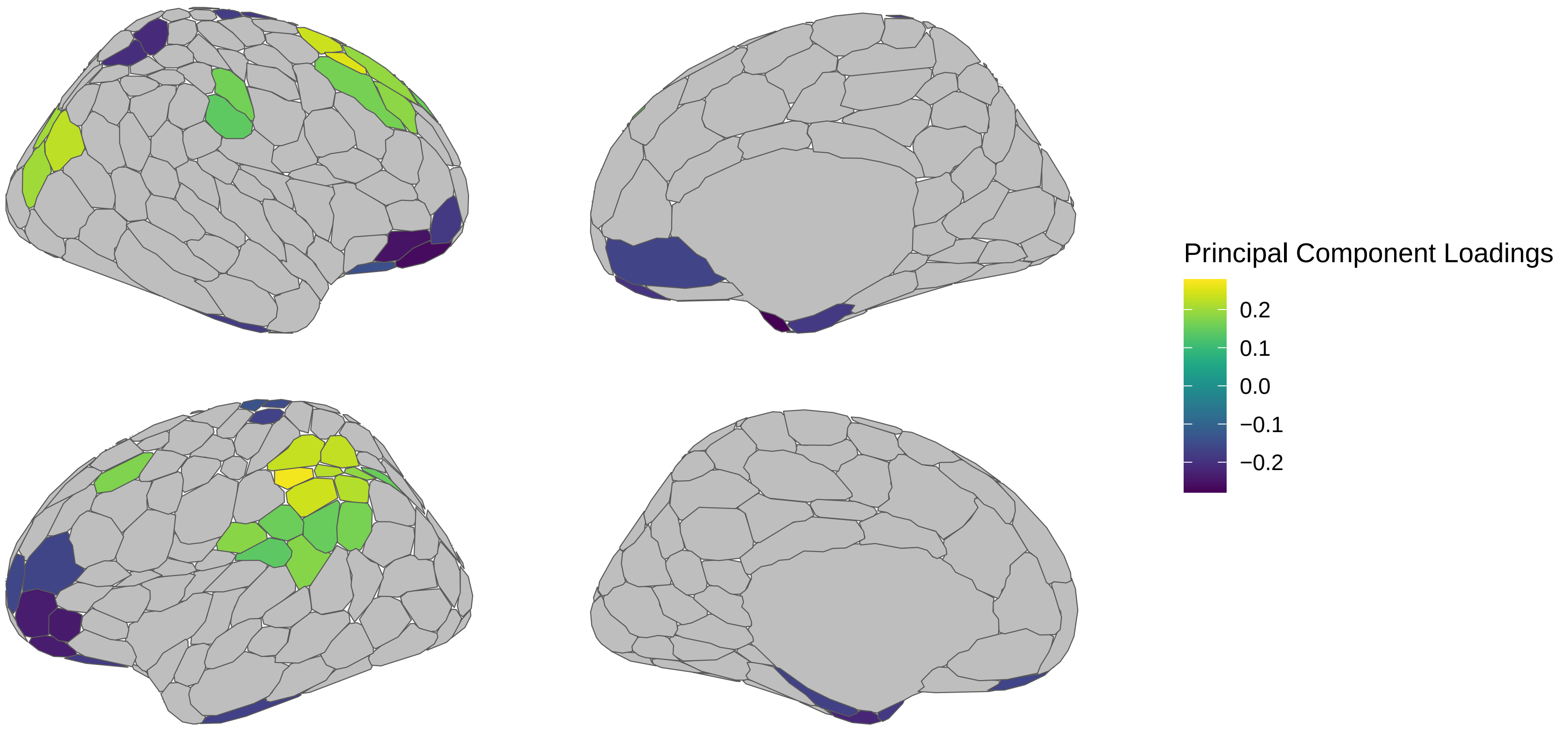


### LH Subcortical Loadings



### RH Subcortical Loadings

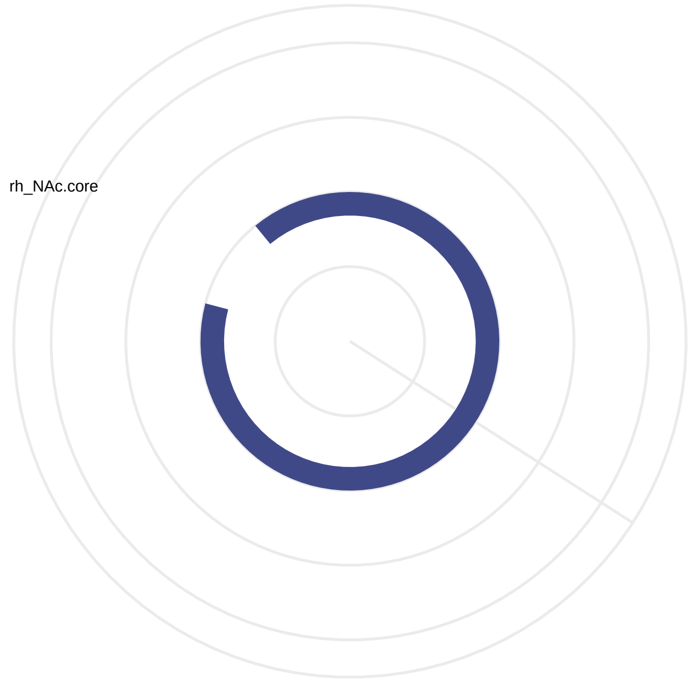

Representation for Principal Component 13

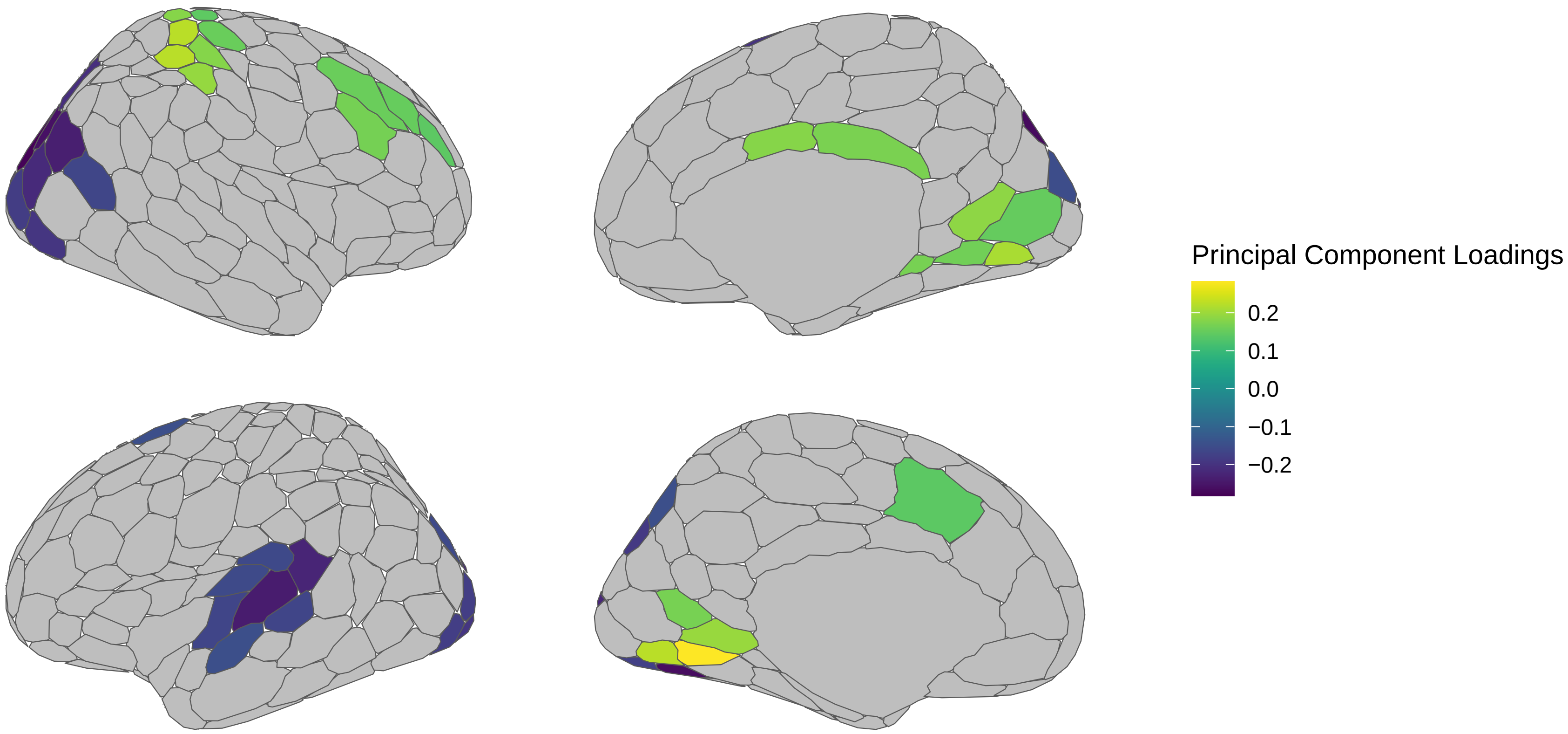


LH Subcortical Loadings



RH Subcortical Loadings

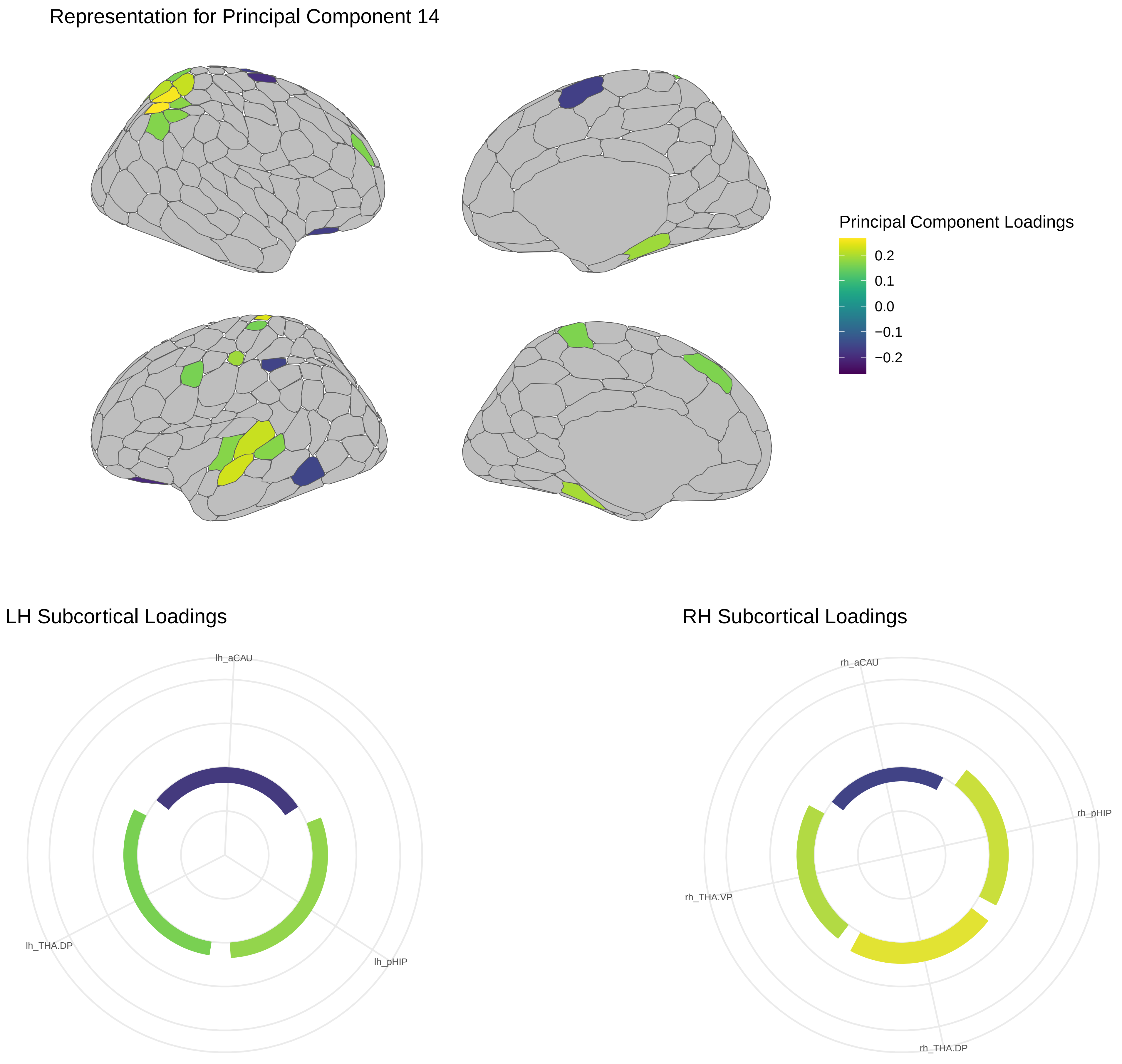
Representation for Principal Component 14
Principal Component Loadings
0.2
0.1
0.0
−0.1
−0.2
LH Subcortical Loadings
lh_aCAU
lh_THA.DP
lh_pHIP
RH Subcortical Loadings
rh_aCAU
rh_pHIP
rh_THA.VP
rh_THA.DP

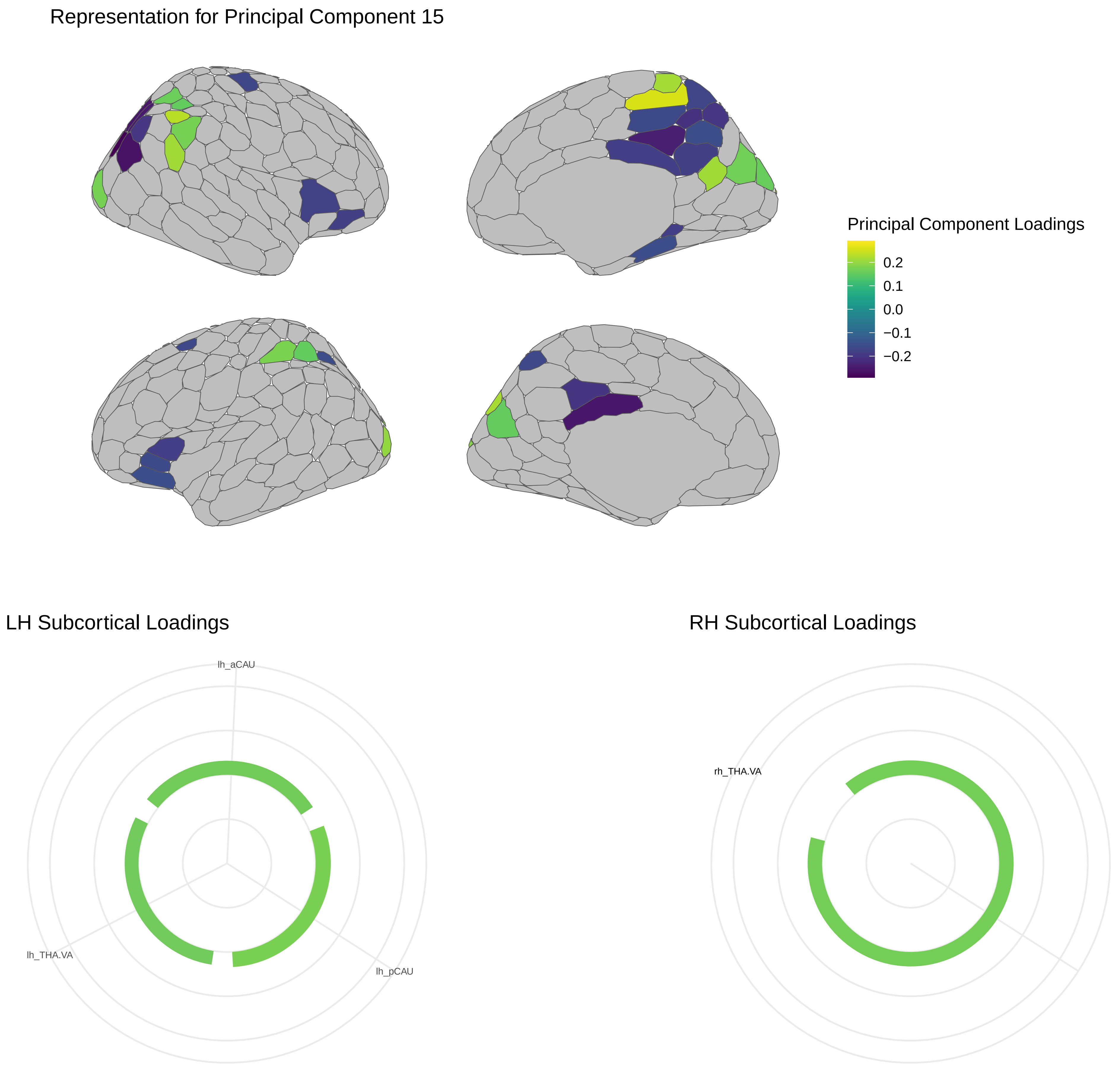
Representation for Principal Component 15
Principal Component Loadings
0.2
0.1
0.0
−0.1
−0.2
LH Subcortical Loadings
lh_aCAU
lh_THA.VA
lh_pCAU
RH Subcortical Loadings
rh_THA.VA

Representation for Principal Component 16

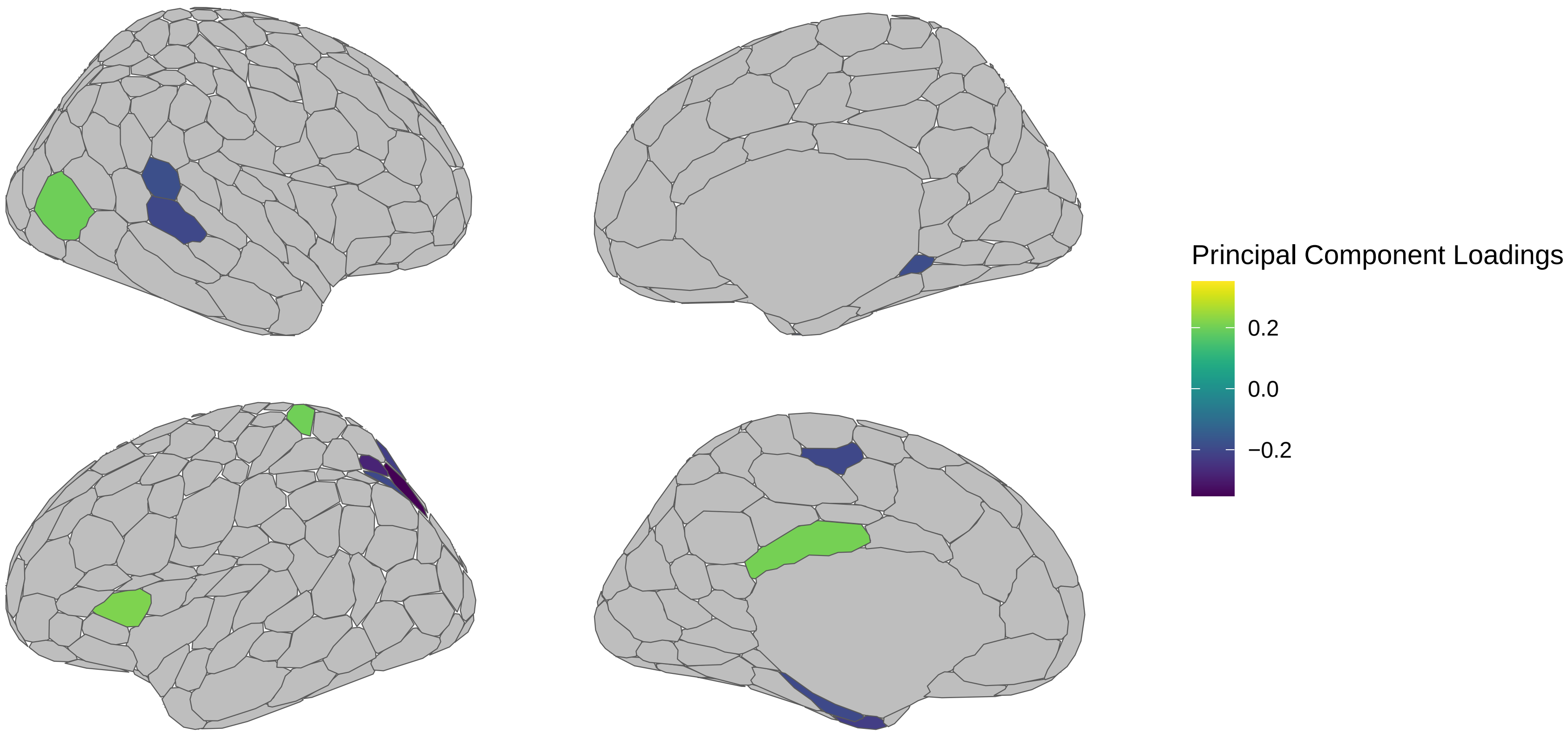


LH Subcortical Loadings



RH Subcortical Loadings

Representation for Principal Component 17

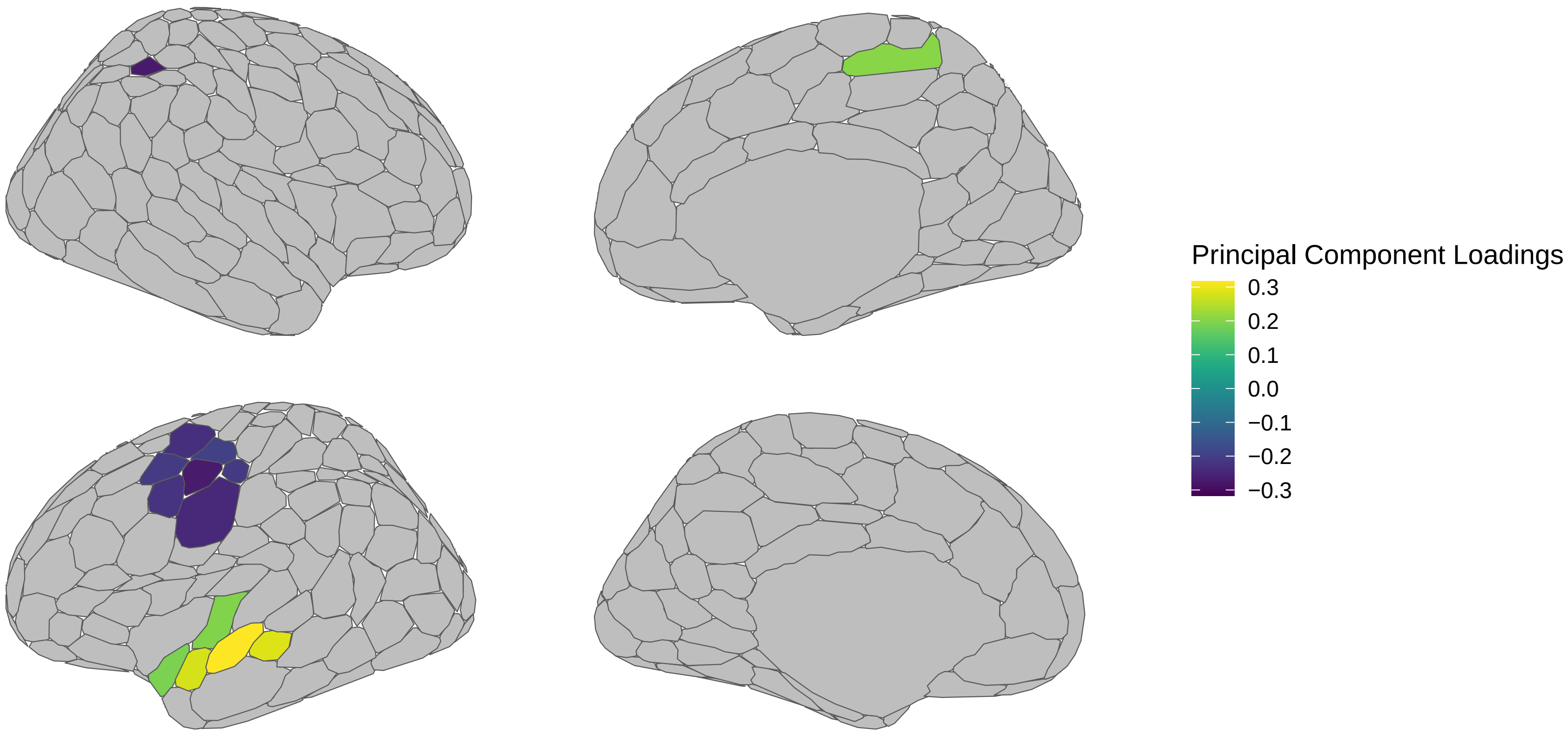


LH Subcortical Loadings



RH Subcortical Loadings

Representation for Principal Component 18

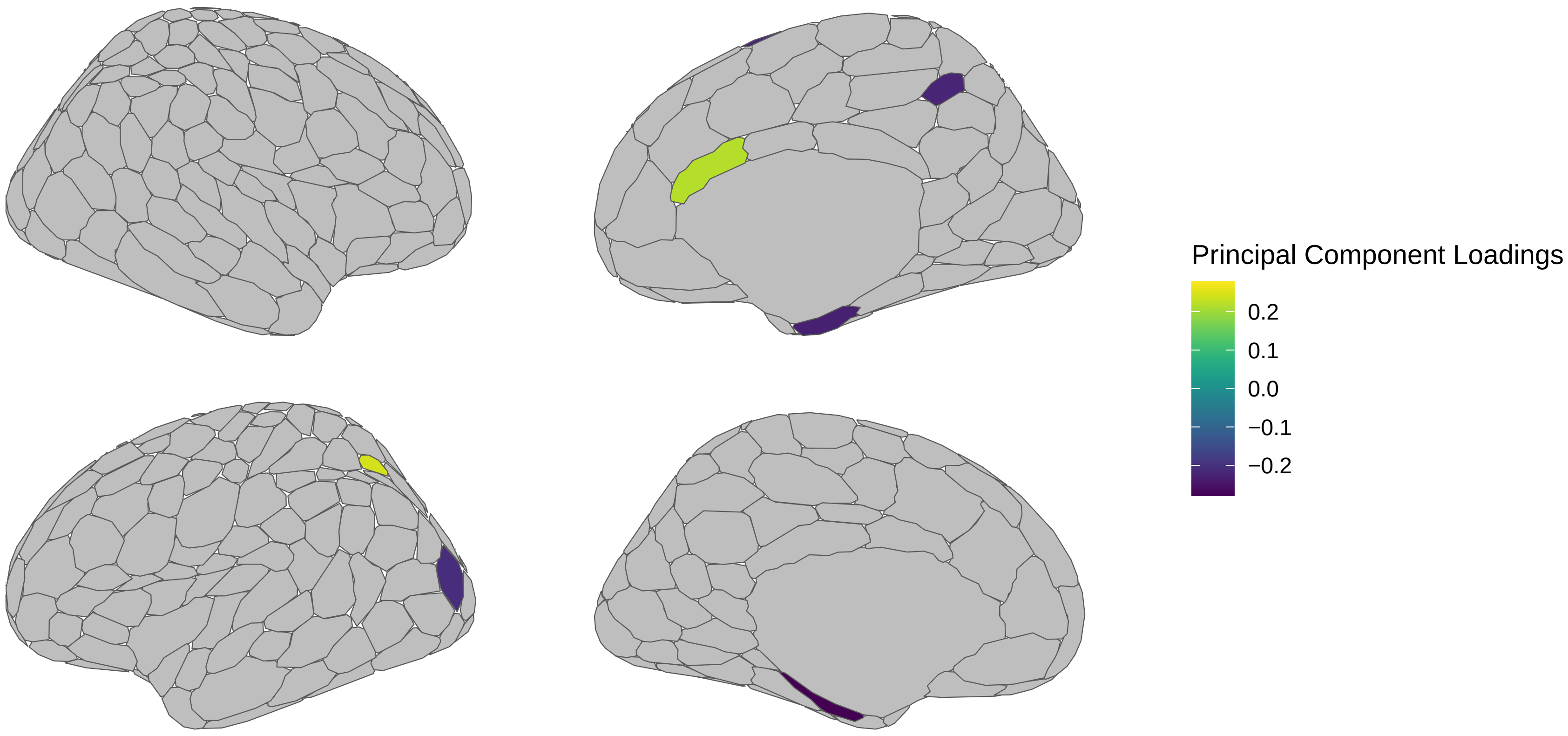


LH Subcortical Loadings



RH Subcortical Loadings

Representation for Principal Component 19

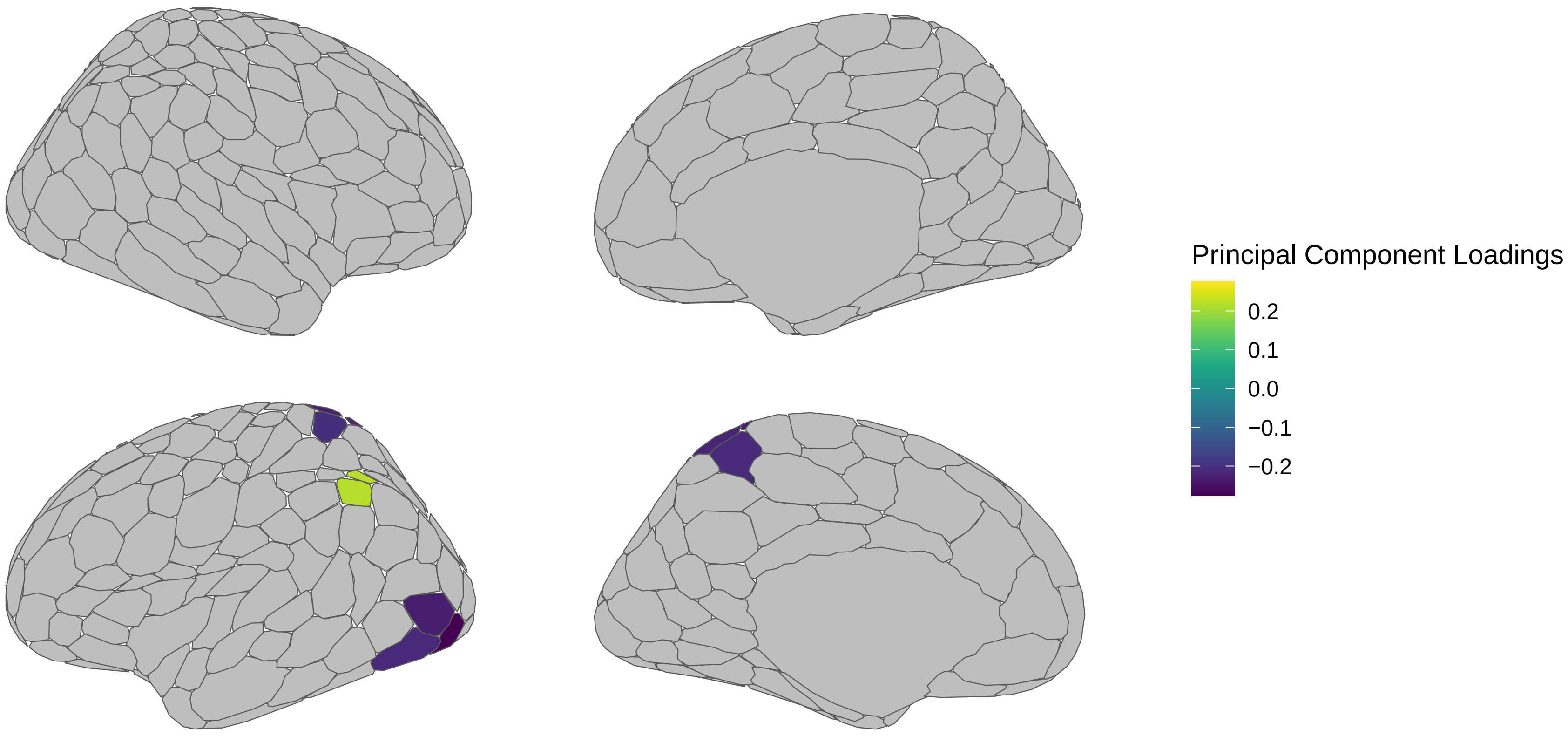


LH Subcortical Loadings



RH Subcortical Loadings

Representation for Principal Component 20

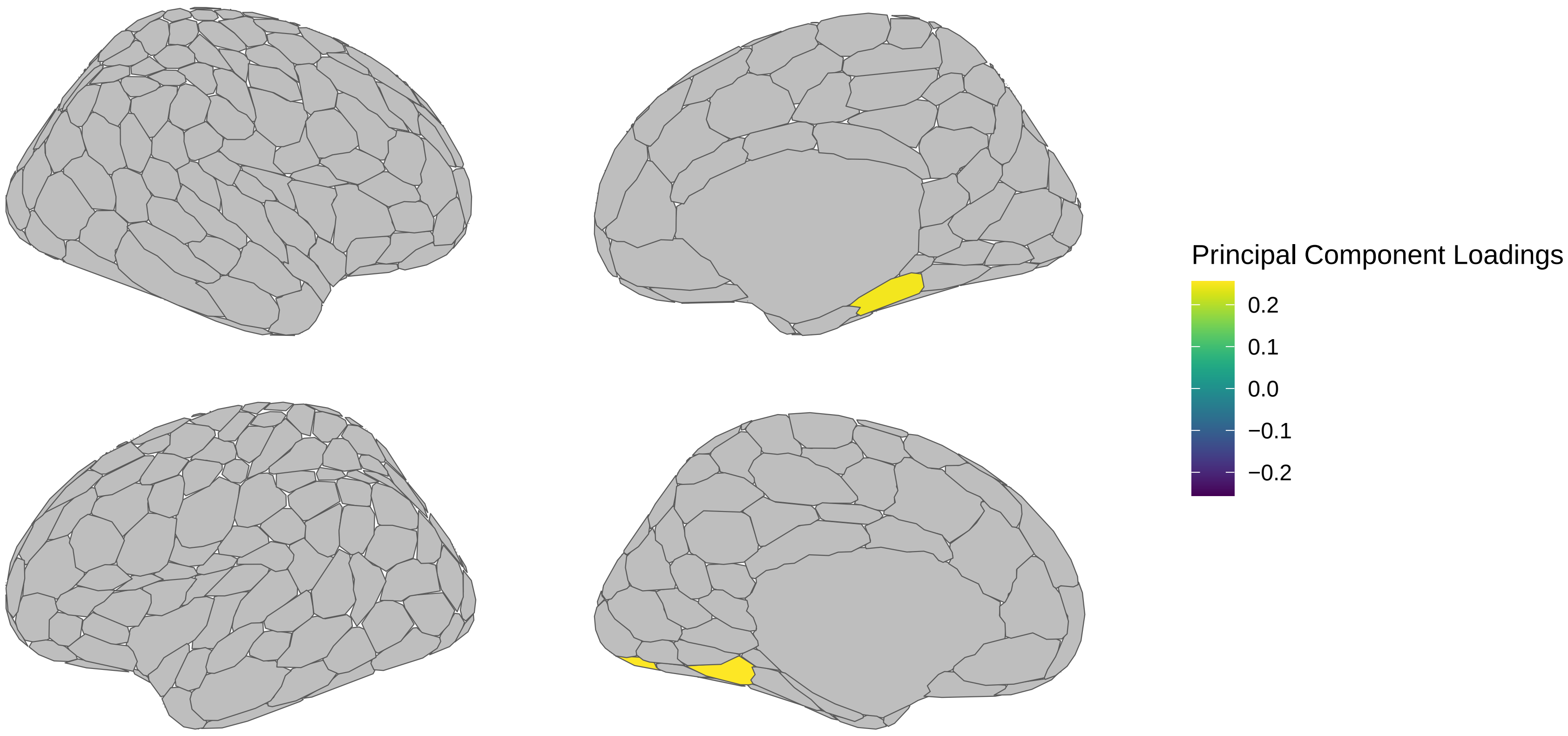


LH Subcortical Loadings



RH Subcortical Loadings

Representation for Principal Component 21

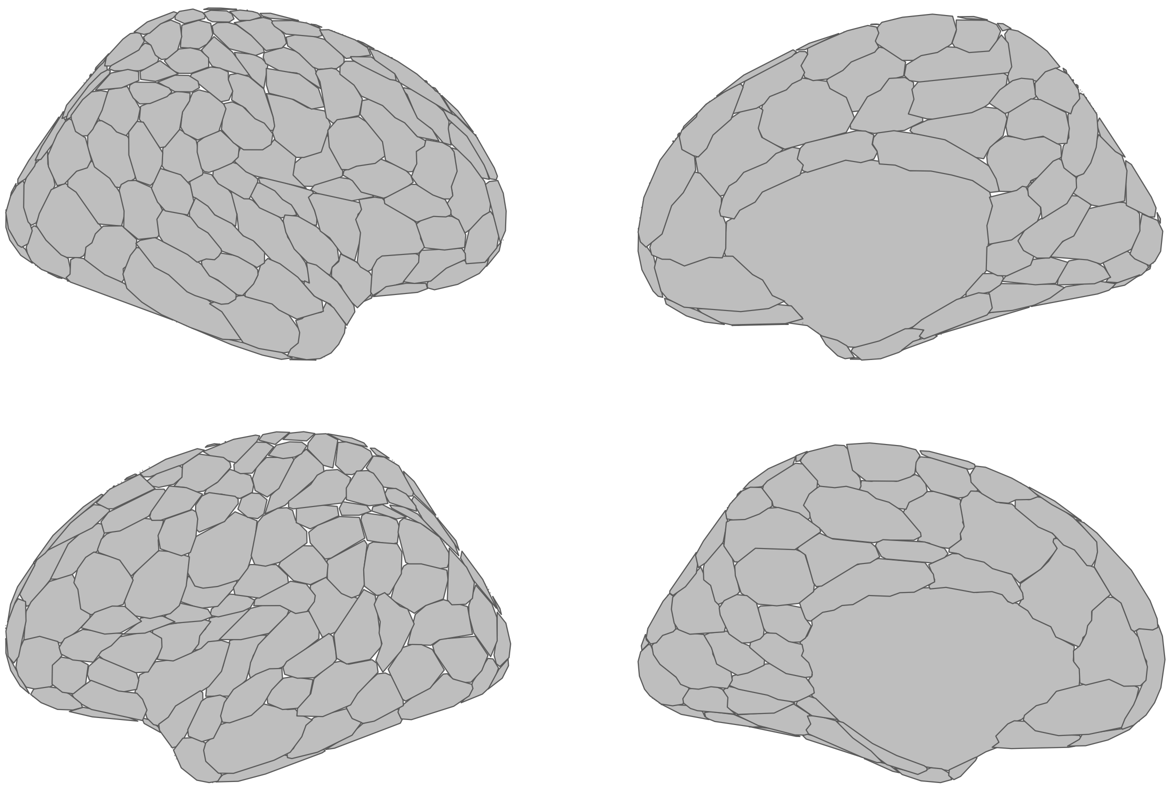

LH Subcortical Loadings



RH Subcortical Loadings

Representation for Principal Component 22

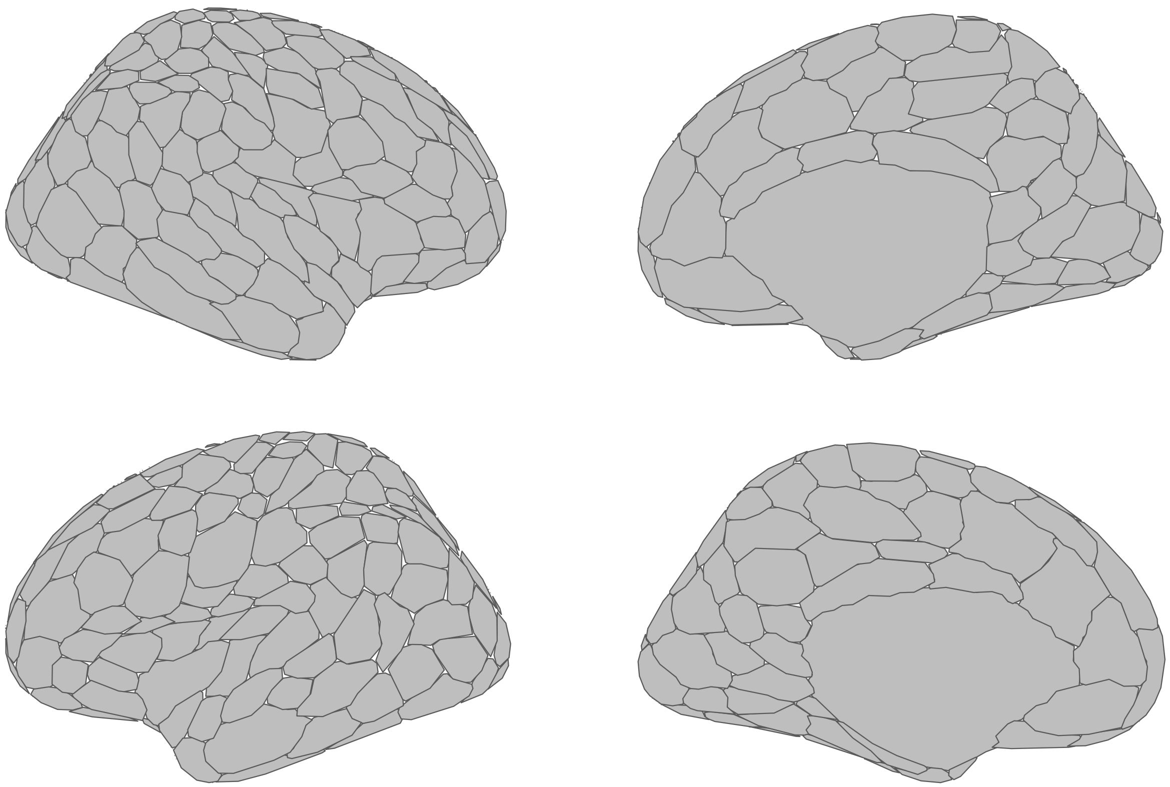

LH Subcortical Loadings



RH Subcortical Loadings
