## Supplementary 5 for "The effects of trauma on neurobiological and psychopathological phenotypes in a transdiagnostic community sample"

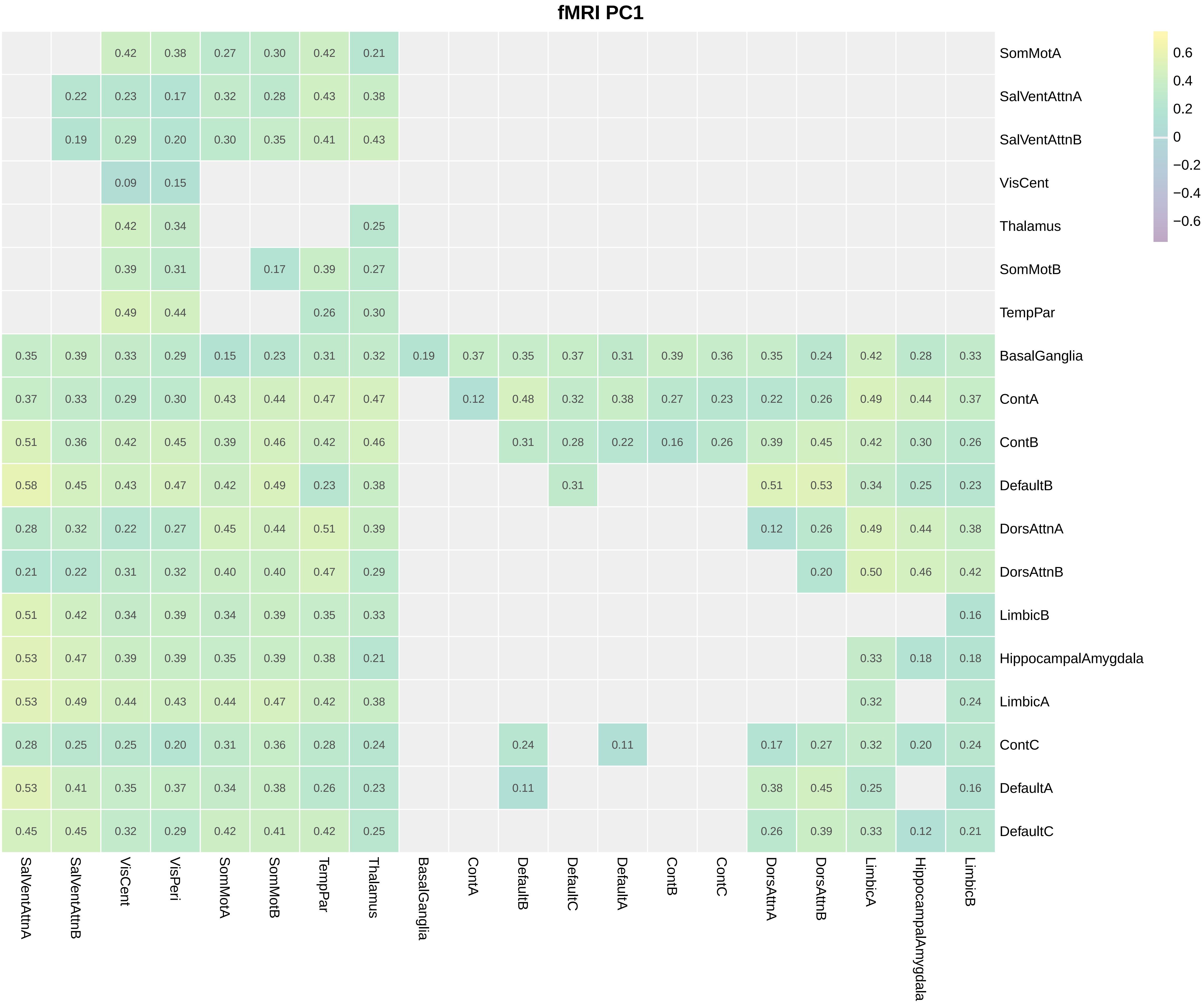
fMRI PC1
SomMotA 0.42 0.38 0.27 0.30 0.42 0.21
SalVentAttnA 0.22 0.23 0.17 0.32 0.28 0.43 0.38
SalVentAttnB 0.19 0.29 0.20 0.30 0.35 0.41 0.43
VisCent 0.09 0.15
Thalamus 0.42 0.34 0.25
SomMotB 0.39 0.31 0.17 0.39 0.27
TempPar 0.49 0.44 0.26 0.30
BasalGanglia 0.35 0.39 0.33 0.29 0.15 0.23 0.31 0.32 0.19 0.37 0.35 0.37 0.31 0.39 0.36 0.35 0.24 0.42 0.28 0.33
ContA 0.37 0.33 0.29 0.30 0.43 0.44 0.47 0.47 0.12 0.48 0.32 0.38 0.27 0.23 0.22 0.26 0.49 0.44 0.37
ContB 0.51 0.36 0.42 0.45 0.39 0.46 0.42 0.46 0.31 0.28 0.22 0.16 0.26 0.39 0.45 0.42 0.30 0.26
DefaultB 0.58 0.45 0.43 0.47 0.42 0.49 0.23 0.38 0.31 0.51 0.53 0.34 0.25 0.23
DorsAttnA 0.28 0.32 0.22 0.27 0.45 0.44 0.51 0.39 0.12 0.26 0.49 0.44 0.38
DorsAttnB 0.21 0.22 0.31 0.32 0.40 0.40 0.47 0.29 0.20 0.50 0.46 0.42
LimbicB 0.51 0.42 0.34 0.39 0.34 0.39 0.35 0.33 0.16
HippocampalAmygdala 0.53 0.47 0.39 0.39 0.35 0.39 0.38 0.21 0.33 0.18 0.18
LimbicA 0.53 0.49 0.44 0.43 0.44 0.47 0.42 0.38 0.32 0.24
ContC 0.28 0.25 0.25 0.20 0.31 0.36 0.28 0.24 0.24 0.11 0.17 0.27 0.32 0.20 0.24
DefaultA 0.53 0.41 0.35 0.37 0.34 0.38 0.26 0.23 0.11 0.38 0.45 0.25 0.16
DefaultC 0.45 0.45 0.32 0.29 0.42 0.41 0.42 0.25 0.26 0.39 0.33 0.12 0.21
SalVentAttnA
SalVentAttnB
VisCent
VisPeri
SomMotA
SomMotB
TempPar
Thalamus
BasalGanglia
ContA
DefaultB
DefaultC
DefaultA
ContB
ContC
DorsAttnA
DorsAttnB
LimbicA
HippocampalAmygdala
LimbicB
0.6
0.4
0.2
0
−0.2
−0.4
−0.6

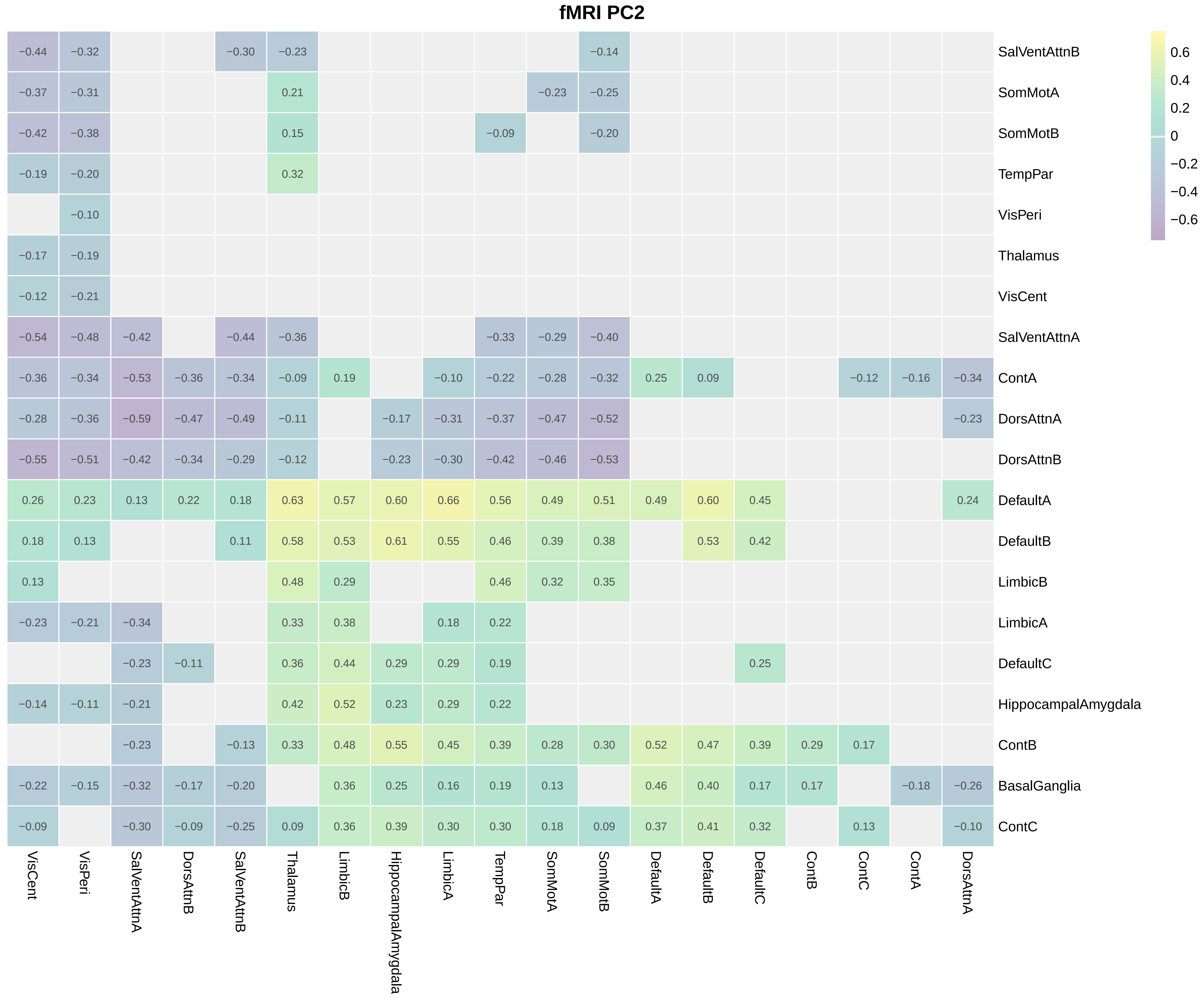
fMRI PC2
SalVentAttnB −0.44 −0.32 −0.30 −0.23 −0.14
SomMotA −0.37 −0.31 0.21 −0.23 −0.25
SomMotB −0.42 −0.38 0.15 −0.09 −0.20
TempPar −0.19 −0.20 0.32
VisPeri −0.10
Thalamus −0.17 −0.19
VisCent −0.12 −0.21
SalVentAttnA −0.54 −0.48 −0.42 −0.44 −0.36 −0.33 −0.29 −0.40
ContA −0.36 −0.34 −0.53 −0.36 −0.34 −0.09 0.19 −0.10 −0.22 −0.28 −0.32 0.25 0.09 −0.12 −0.16 −0.34
DorsAttnA −0.28 −0.36 −0.59 −0.47 −0.49 −0.11 −0.17 −0.31 −0.37 −0.47 −0.52 −0.23
DorsAttnB −0.55 −0.51 −0.42 −0.34 −0.29 −0.12 −0.23 −0.30 −0.42 −0.46 −0.53
DefaultA 0.26 0.23 0.13 0.22 0.18 0.63 0.57 0.60 0.66 0.56 0.49 0.51 0.49 0.60 0.45 0.24
DefaultB 0.18 0.13 0.11 0.58 0.53 0.61 0.55 0.46 0.39 0.38 0.53 0.42
LimbicB 0.13 0.48 0.29 0.46 0.32 0.35
LimbicA −0.23 −0.21 −0.34 0.33 0.38 0.18 0.22
DefaultC −0.23 −0.11 0.36 0.44 0.29 0.29 0.19 0.25
HippocampalAmygdala −0.14 −0.11 −0.21 0.42 0.52 0.23 0.29 0.22
ContB −0.23 −0.13 0.33 0.48 0.55 0.45 0.39 0.28 0.30 0.52 0.47 0.39 0.29 0.17
BasalGanglia −0.22 −0.15 −0.32 −0.17 −0.20 0.36 0.25 0.16 0.19 0.13 0.46 0.40 0.17 0.17 −0.18 −0.26
ContC −0.09 −0.30 −0.09 −0.25 0.09 0.36 0.39 0.30 0.30 0.18 0.09 0.37 0.41 0.32 0.13 −0.10
VisCent
VisPeri
SalVentAttnA
DorsAttnB
SalVentAttnB
Thalamus
LimbicB
HippocampalAmygdala
LimbicA
TempPar
SomMotA
SomMotB
DefaultA
DefaultB
DefaultC
ContB
ContC
ContA
DorsAttnA
0.6
0.4
0.2
0
−0.2
−0.4
−0.6

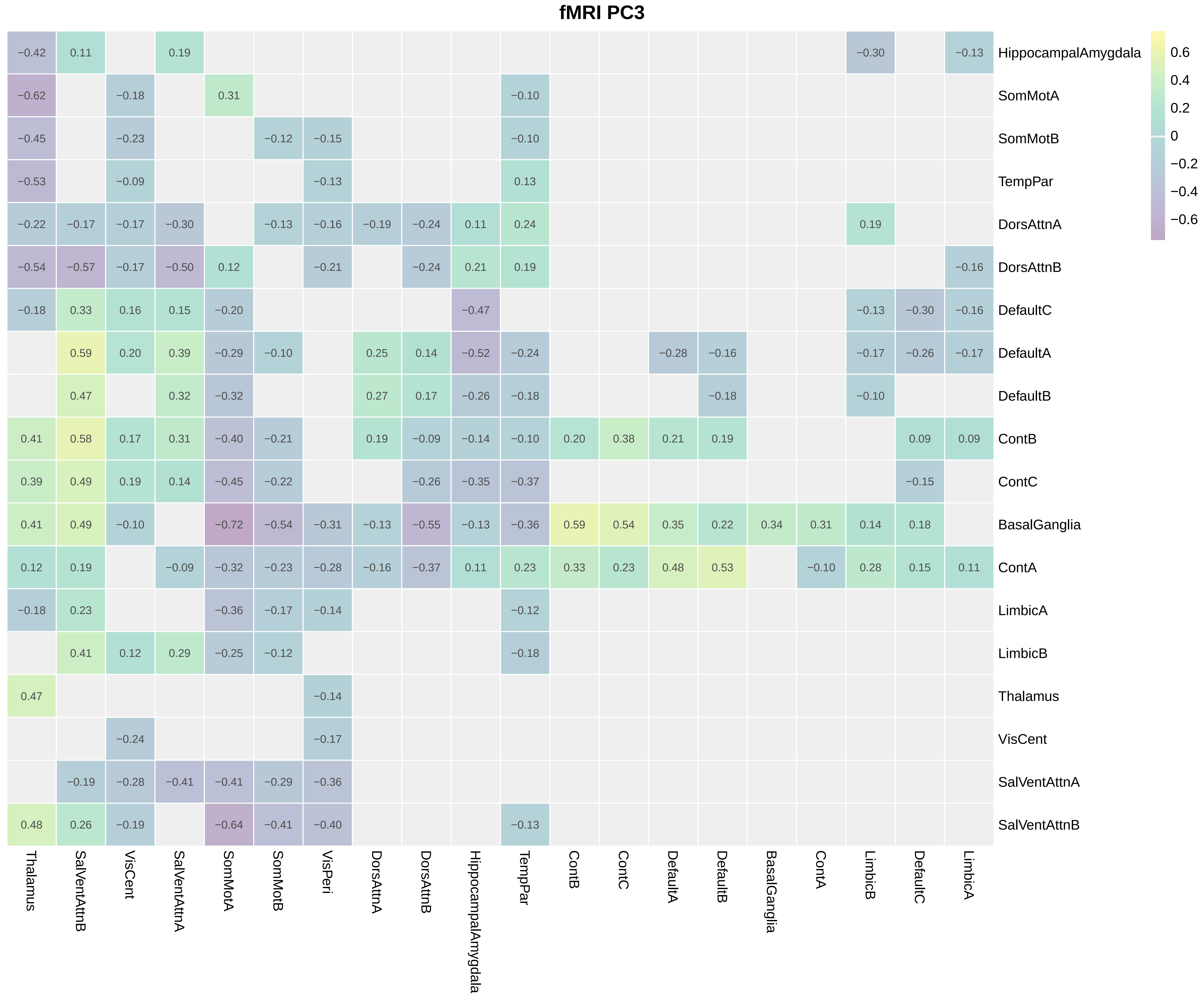
fMRI PC3
0.6
0.4
0.2
0
−0.2
−0.4
−0.6
HippocampalAmygdala
SomMotA
SomMotB
TempPar
DorsAttnA
DorsAttnB
DefaultC
DefaultA
DefaultB
ContB
ContC
BasalGanglia
ContA
LimbicA
LimbicB
Thalamus
VisCent
SalVentAttnA
SalVentAttnB
Thalamus
SalVentAttnB
VisCent
SalVentAttnA
SomMotA
SomMotB
VisPeri
DorsAttnA
DorsAttnB
HippocampalAmygdala
TempPar
ContB
ContC
DefaultA
DefaultB
BasalGanglia
ContA
LimbicB
DefaultC
LimbicA

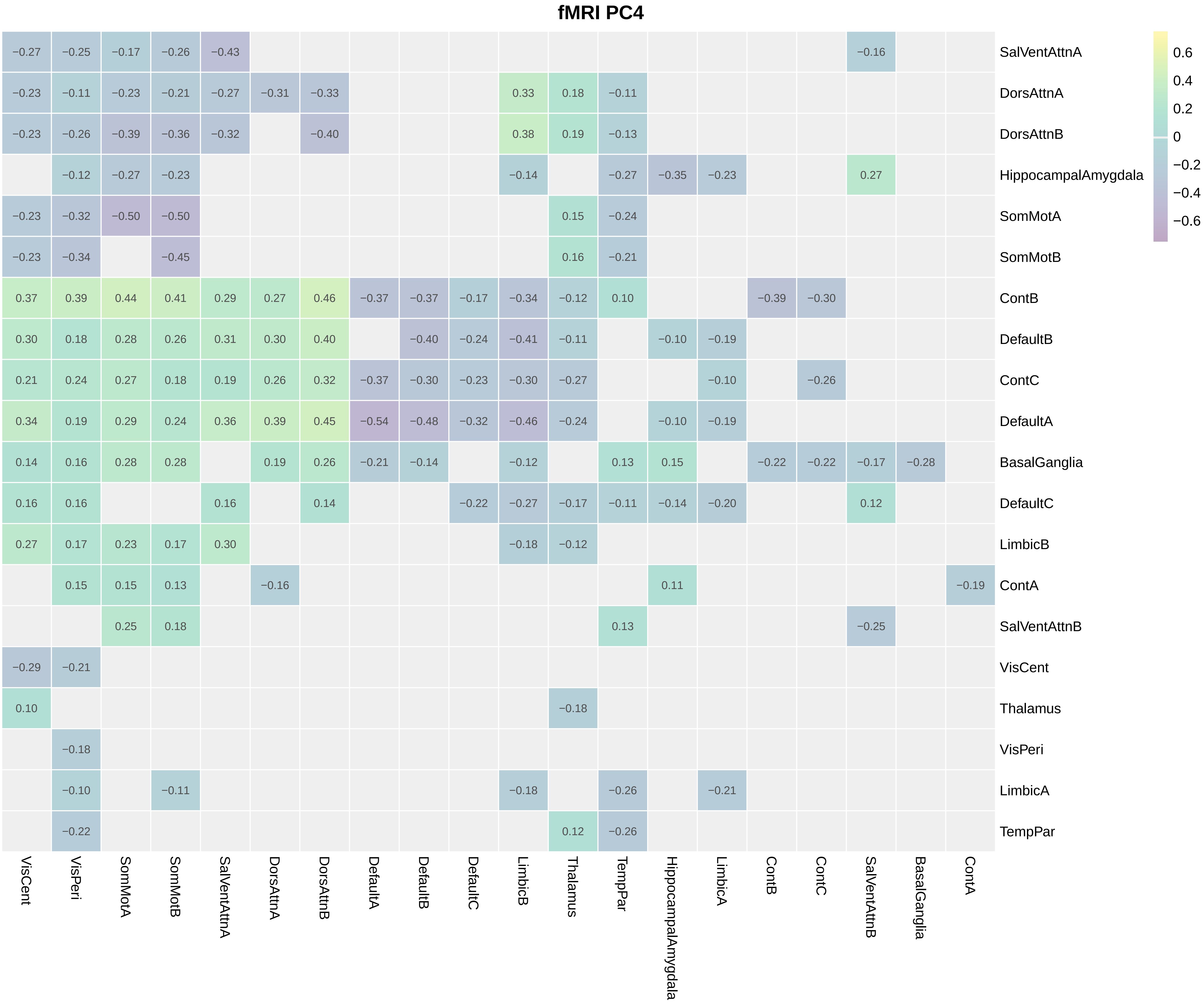
fMRI PC4
SalVentAttnA
−0.27 −0.25 −0.17 −0.26 −0.43 −0.16
DorsAttnA
−0.23 −0.11 −0.23 −0.21 −0.27 −0.31 −0.33 0.33 0.18 −0.11
DorsAttnB
−0.23 −0.26 −0.39 −0.36 −0.32 −0.40 0.38 0.19 −0.13
HippocampalAmygdala
−0.12 −0.27 −0.23 −0.14 −0.27 −0.35 −0.23 0.27
SomMotA
−0.23 −0.32 −0.50 −0.50 0.15 −0.24
SomMotB
−0.23 −0.34 −0.45 0.16 −0.21
ContB
0.37 0.39 0.44 0.41 0.29 0.27 0.46 −0.37 −0.37 −0.17 −0.34 −0.12 0.10 −0.39 −0.30
DefaultB
0.30 0.18 0.28 0.26 0.31 0.30 0.40 −0.40 −0.24 −0.41 −0.11 −0.10 −0.19
ContC
0.21 0.24 0.27 0.18 0.19 0.26 0.32 −0.37 −0.30 −0.23 −0.30 −0.27 −0.10 −0.26
DefaultA
0.34 0.19 0.29 0.24 0.36 0.39 0.45 −0.54 −0.48 −0.32 −0.46 −0.24 −0.10 −0.19
BasalGanglia
0.14 0.16 0.28 0.28 0.19 0.26 −0.21 −0.14 −0.12 0.13 0.15 −0.22 −0.22 −0.17 −0.28
DefaultC
0.16 0.16 0.16 0.14 −0.22 −0.27 −0.17 −0.11 −0.14 −0.20 0.12
LimbicB
0.27 0.17 0.23 0.17 0.30 −0.18 −0.12
ContA
0.15 0.15 0.13 −0.16 0.11 −0.19
SalVentAttnB
0.25 0.18 0.13 −0.25
VisCent
−0.29 −0.21
Thalamus
0.10 −0.18
VisPeri
−0.18
LimbicA
−0.10 −0.11 −0.18 −0.26 −0.21
TempPar
−0.22 0.12 −0.26
VisCent VisPeri SomMotA SomMotB SalVentAttnA DorsAttnA DorsAttnB DefaultA DefaultB DefaultC LimbicB Thalamus TempPar HippocampalAmygdala LimbicA ContB ContC SalVentAttnB BasalGanglia ContA
0.6
0.4
0.2
0
−0.2
−0.4
−0.6

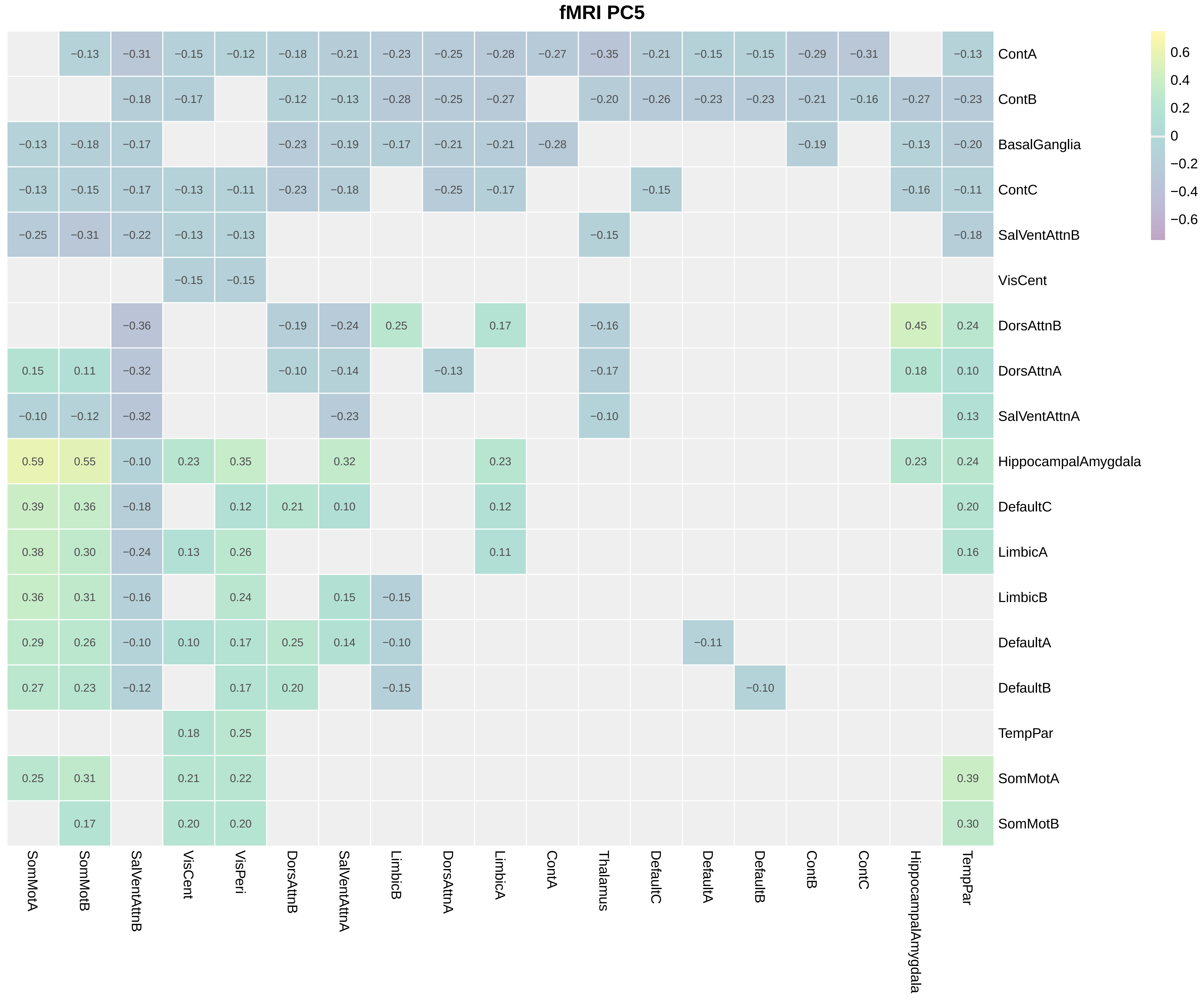

fMRI PC5
ContA
ContB
BasalGanglia
ContC
SalVentAttnB
VisCent
DorsAttnB
DorsAttnA
SalVentAttnA
HippocampalAmygdala
DefaultC
LimbicA
LimbicB
DefaultA
DefaultB
TempPar
SomMotA
SomMotB
SomMotA
SomMotB
SalVentAttnB
VisCent
VisPeri
DorsAttnB
SalVentAttnA
LimbicB
DorsAttnA
LimbicA
ContA
Thalamus
DefaultC
DefaultA
DefaultB
ContB
ContC
HippocampalAmygdala
TempPar
0.6
0.4
0.2
0
−0.2
−0.4
−0.6

**fMRI PC6**

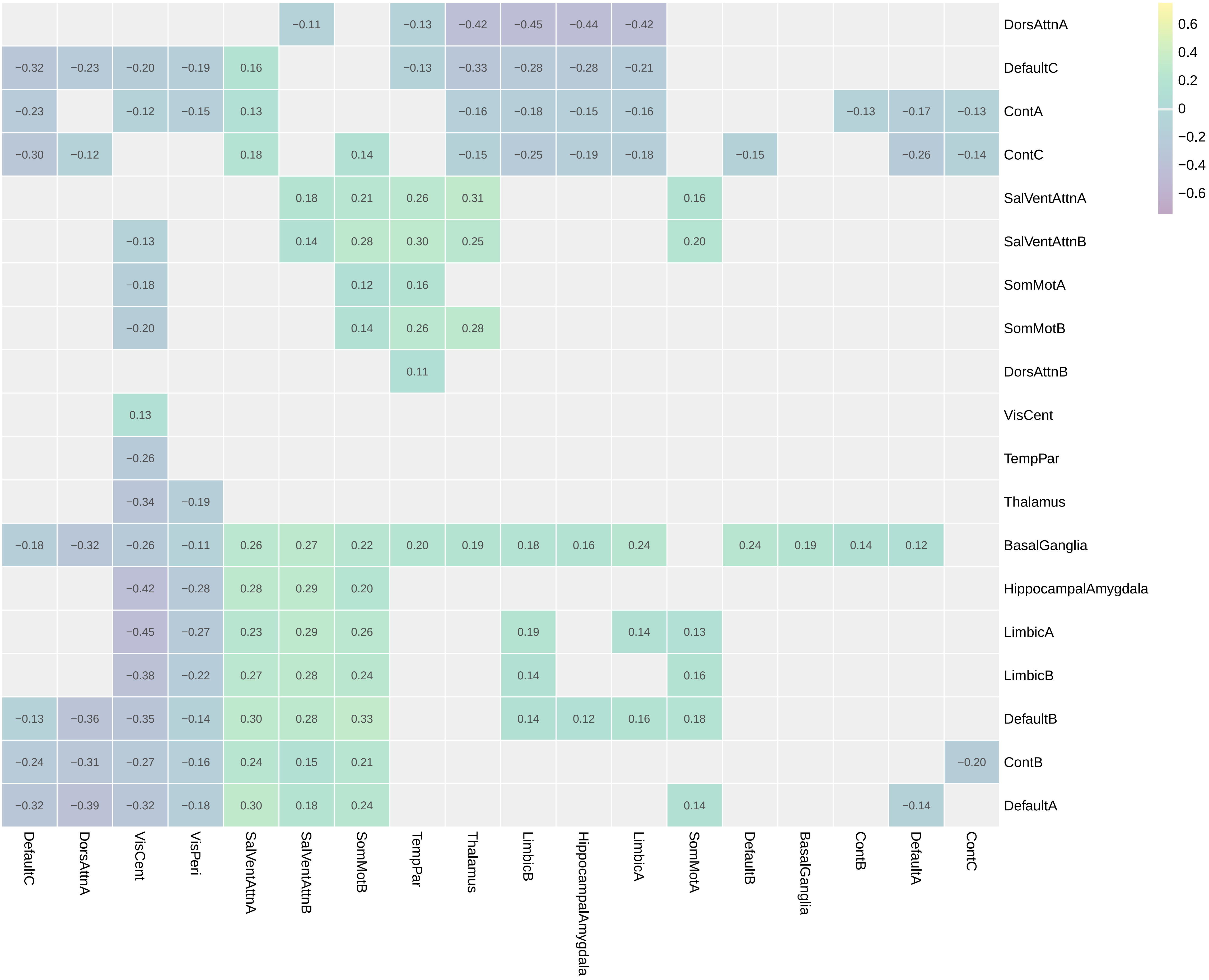
DorsAttnA −0.11 −0.13 −0.42 −0.45 −0.44 −0.42
DefaultC −0.32 −0.23 −0.20 −0.19 0.16 −0.13 −0.33 −0.28 −0.28 −0.21
ContA −0.23 −0.12 −0.15 0.13 −0.16 −0.18 −0.15 −0.16 −0.13 −0.17 −0.13
ContC −0.30 −0.12 0.18 0.14 −0.15 −0.25 −0.19 −0.18 −0.15 −0.26 −0.14
SalVentAttnA 0.18 0.21 0.26 0.31 0.16
SalVentAttnB −0.13 0.14 0.28 0.30 0.25 0.20
SomMotA −0.18 0.12 0.16
SomMotB −0.20 0.14 0.26 0.28
DorsAttnB 0.11
VisCent 0.13
TempPar −0.26
Thalamus −0.34 −0.19
BasalGanglia −0.18 −0.32 −0.26 −0.11 0.26 0.27 0.22 0.20 0.19 0.18 0.16 0.24 0.24 0.19 0.14 0.12
HippocampalAmygdala −0.42 −0.28 0.28 0.29 0.20
LimbicA −0.45 −0.27 0.23 0.29 0.26 0.19 0.14 0.13
LimbicB −0.38 −0.22 0.27 0.28 0.24 0.14 0.16
DefaultB −0.13 −0.36 −0.35 −0.14 0.30 0.28 0.33 0.14 0.12 0.16 0.18
ContB −0.24 −0.31 −0.27 −0.16 0.24 0.15 0.21 −0.20
DefaultA −0.32 −0.39 −0.32 −0.18 0.30 0.18 0.24 0.14 −0.14
DefaultC
DorsAttnA
VisCent
VisPeri
SalVentAttnA
SalVentAttnB
SomMotB
TempPar
Thalamus
LimbicB
HippocampalAmygdala
LimbicA
SomMotA
DefaultB
BasalGanglia
ContB
DefaultA
ContC
0.6
0.4
0.2
0
−0.2
−0.4
−0.6

**fMRI PC7**

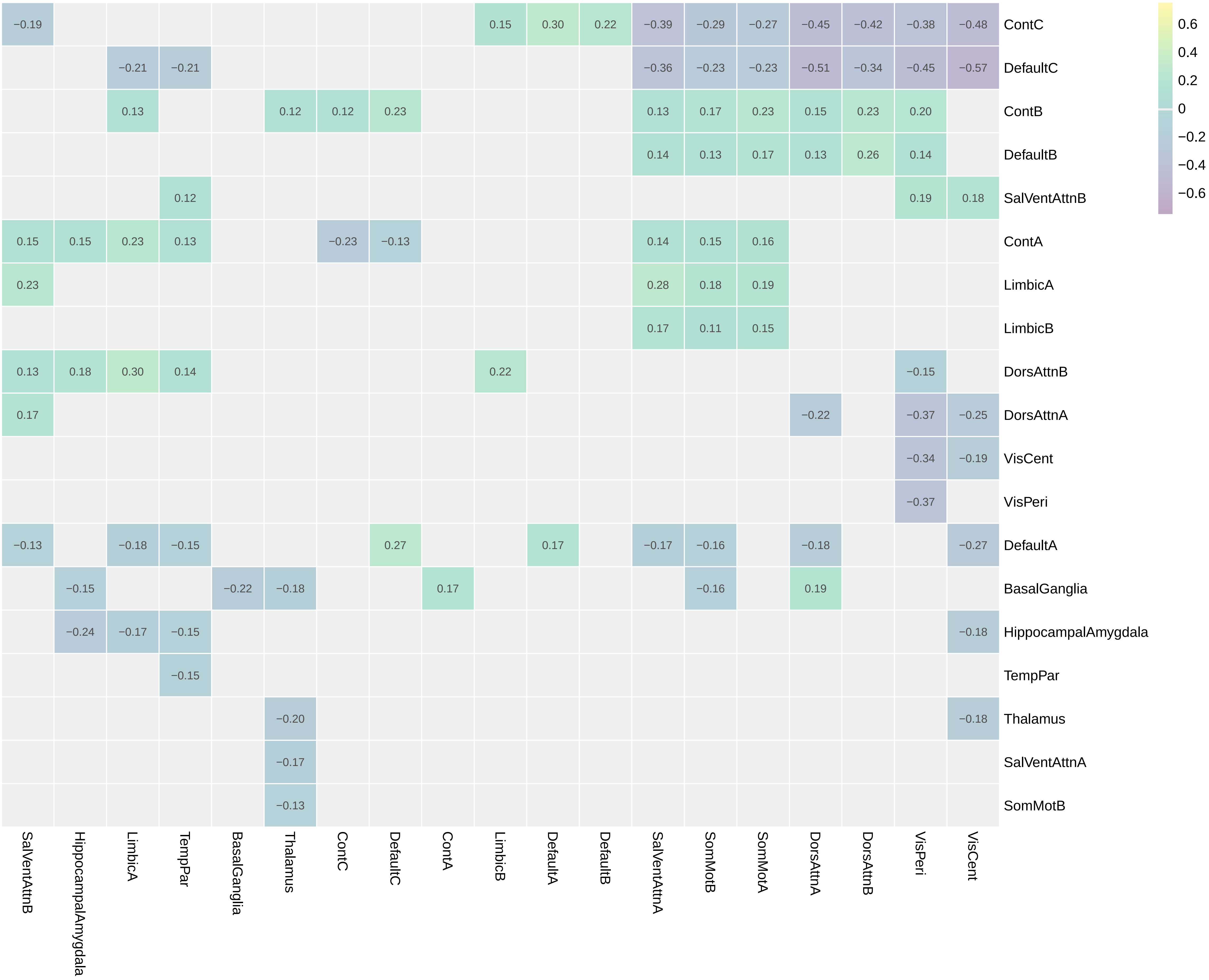
0.6
0.4
0.2
0
−0.2
−0.4
−0.6
ContC
DefaultC
ContB
DefaultB
SalVentAttnB
ContA
LimbicA
LimbicB
DorsAttnB
DorsAttnA
VisCent
VisPeri
DefaultA
BasalGanglia
HippocampalAmygdala
TempPar
Thalamus
SalVentAttnA
SomMotB
SalVentAttnB
HippocampalAmygdala
LimbicA
TempPar
BasalGanglia
Thalamus
ContC
DefaultC
ContA
LimbicB
DefaultA
DefaultB
SalVentAttnA
SomMotB
SomMotA
DorsAttnA
DorsAttnB
VisPeri
VisCent

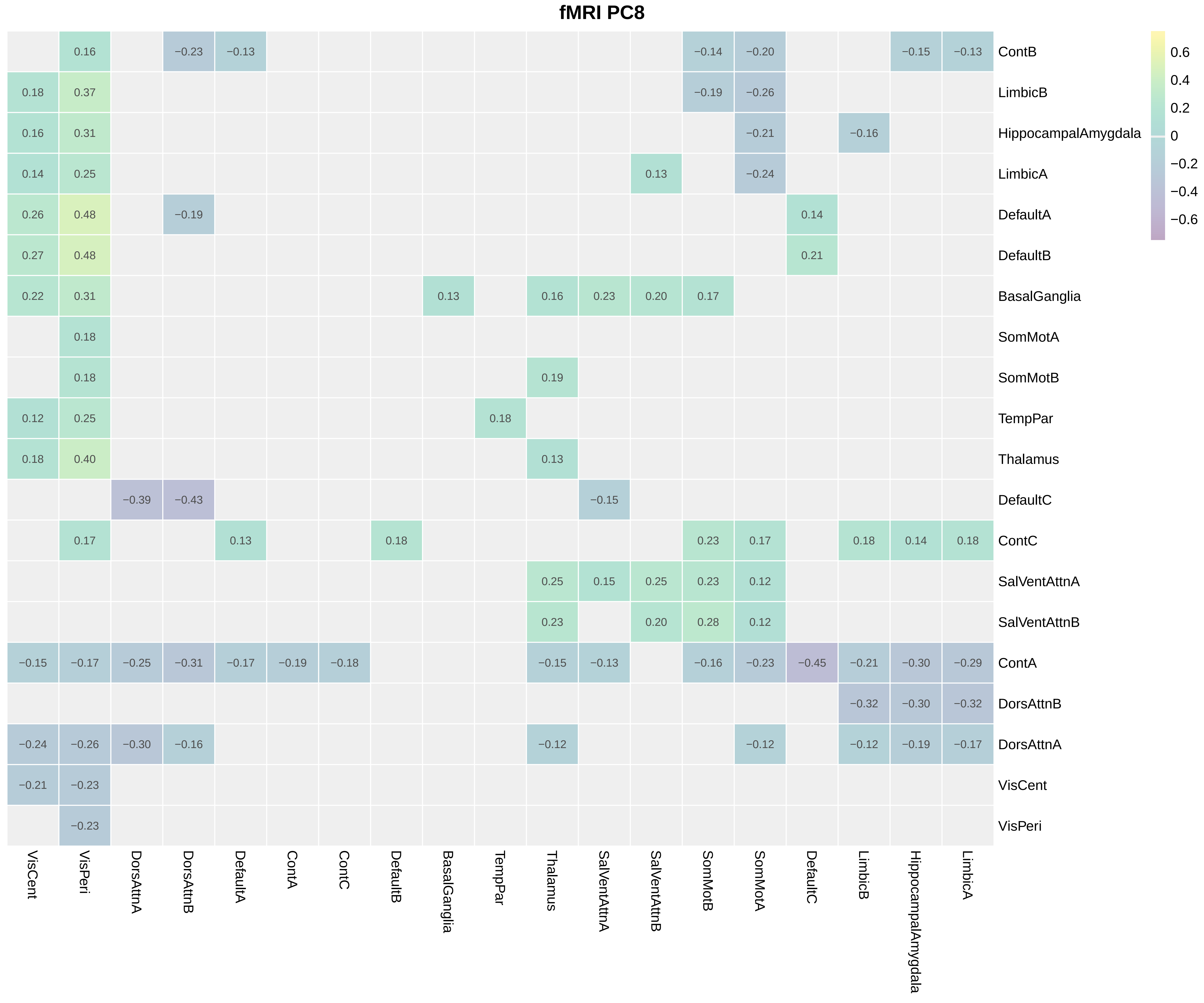
fMRI PC8
ContB
LimbicB
HippocampalAmygdala
LimbicA
DefaultA
DefaultB
BasalGanglia
SomMotA
SomMotB
TempPar
Thalamus
DefaultC
ContC
SalVentAttnA
SalVentAttnB
ContA
DorsAttnB
DorsAttnA
VisCent
VisPeri
VisCent
VisPeri
DorsAttnA
DorsAttnB
DefaultA
ContA
ContC
DefaultB
BasalGanglia
TempPar
Thalamus
SalVentAttnA
SalVentAttnB
SomMotB
SomMotA
DefaultC
LimbicB
HippocampalAmygdala
LimbicA
0.6
0.4
0.2
0
−0.2
−0.4
−0.6

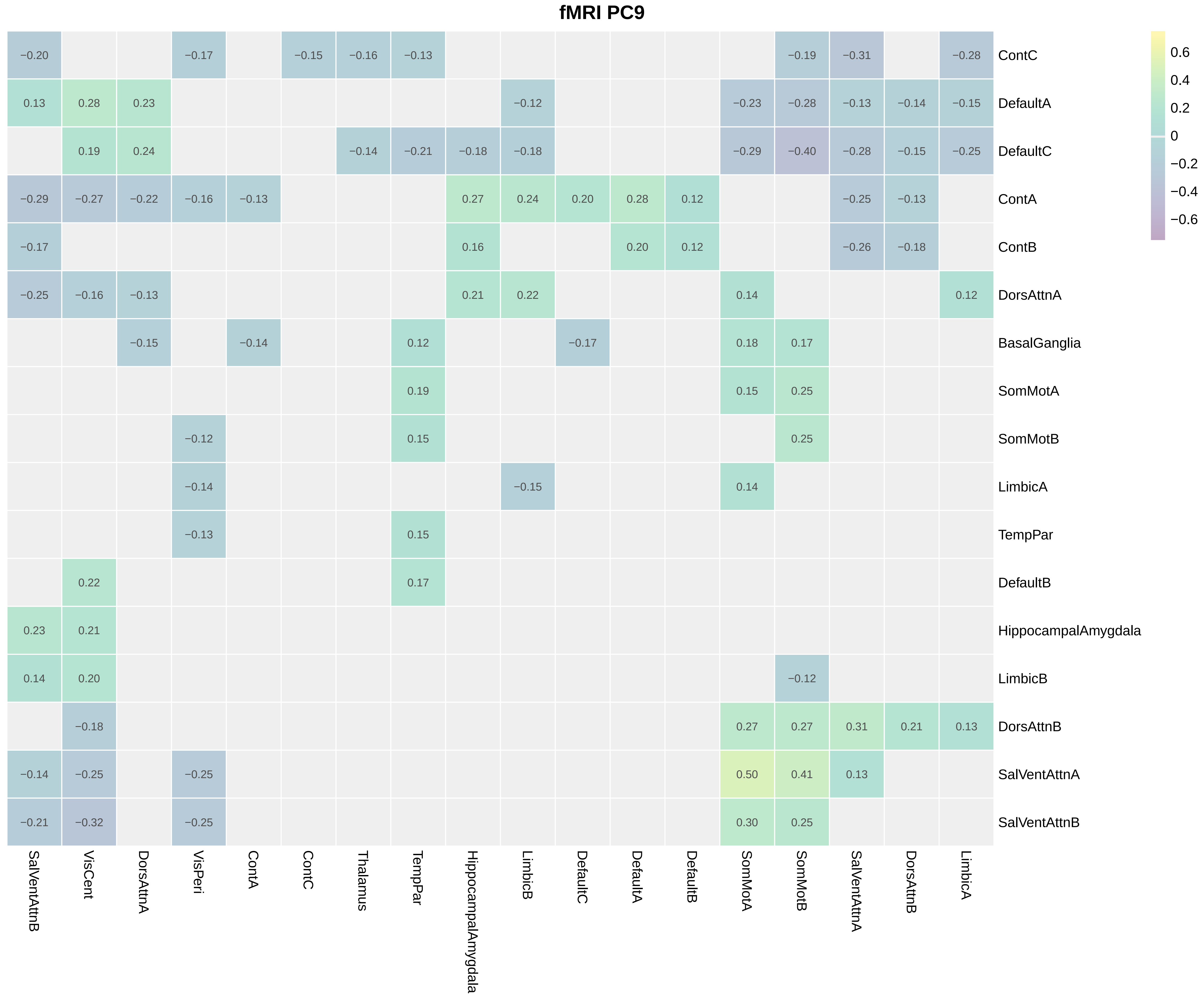

fMRI PC9
ContC
DefaultA
DefaultC
ContA
ContB
DorsAttnA
BasalGanglia
SomMotA
SomMotB
LimbicA
TempPar
DefaultB
HippocampalAmygdala
LimbicB
DorsAttnB
SalVentAttnA
SalVentAttnB
SalVentAttnB
VisCent
DorsAttnA
VisPeri
ContA
ContC
Thalamus
TempPar
HippocampalAmygdala
LimbicB
DefaultC
DefaultA
DefaultB
SomMotA
SomMotB
SalVentAttnA
DorsAttnB
LimbicA
0.6
0.4
0.2
0
−0.2
−0.4
−0.6

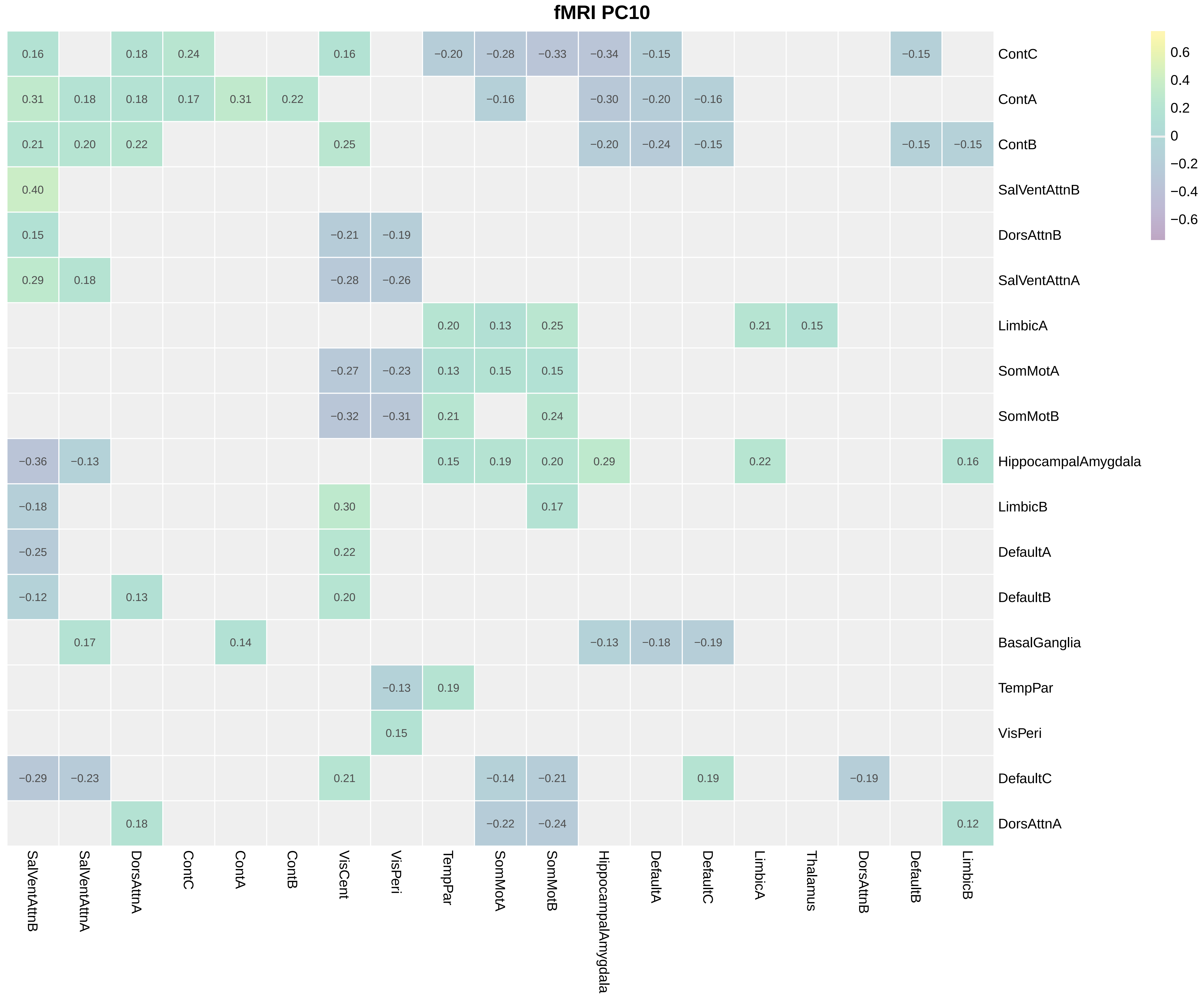
fMRI PC10
ContC
ContA
ContB
SalVentAttnB
DorsAttnB
SalVentAttnA
LimbicA
SomMotA
SomMotB
HippocampalAmygdala
LimbicB
DefaultA
DefaultB
BasalGanglia
TempPar
VisPeri
DefaultC
DorsAttnA
SalVentAttnB
SalVentAttnA
DorsAttnA
ContC
ContA
ContB
VisCent
VisPeri
TempPar
SomMotA
SomMotB
HippocampalAmygdala
DefaultA
DefaultC
LimbicA
Thalamus
DorsAttnB
DefaultB
LimbicB
0.6
0.4
0.2
0
−0.2
−0.4
−0.6

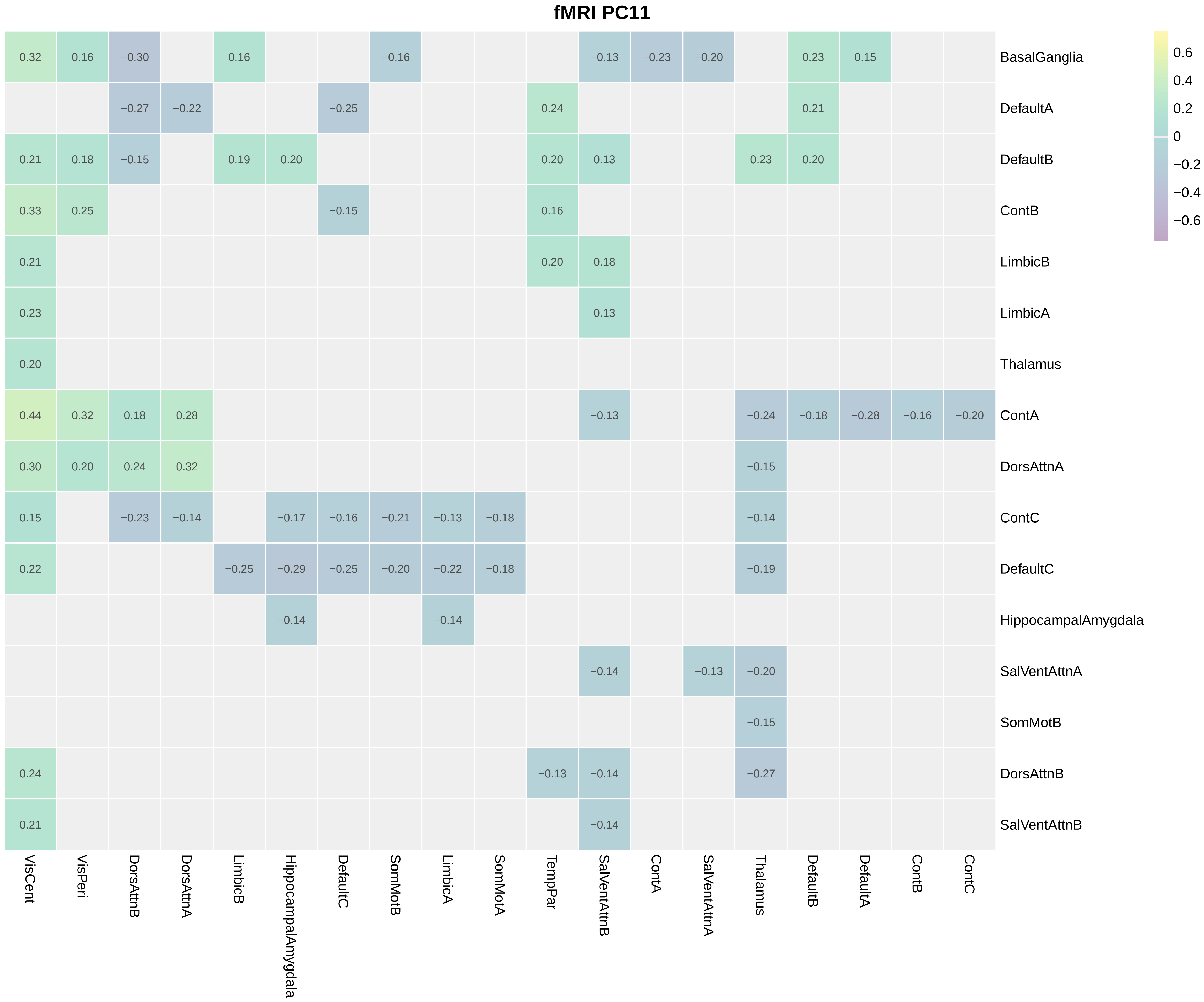
fMRI PC11
0.32 0.16 −0.30 0.16 −0.16 −0.13 −0.23 −0.20 0.23 0.15 BasalGanglia
−0.27 −0.22 −0.25 0.24 0.21 DefaultA
0.21 0.18 −0.15 0.19 0.20 0.20 0.13 0.23 0.20 DefaultB
0.33 0.25 −0.15 0.16 ContB
0.21 0.20 0.18 LimbicB
0.23 0.13 LimbicA
0.20 Thalamus
0.44 0.32 0.18 0.28 −0.13 −0.24 −0.18 −0.28 −0.16 −0.20 ContA
0.30 0.20 0.24 0.32 −0.15 DorsAttnA
0.15 −0.23 −0.14 −0.17 −0.16 −0.21 −0.13 −0.18 −0.14 ContC
0.22 −0.25 −0.29 −0.25 −0.20 −0.22 −0.18 −0.19 DefaultC
−0.14 −0.14 HippocampalAmygdala
−0.14 −0.13 −0.20 SalVentAttnA
−0.15 SomMotB
0.24 −0.13 −0.14 −0.27 DorsAttnB
0.21 −0.14 SalVentAttnB
VisCent
VisPeri
DorsAttnB
DorsAttnA
LimbicB
HippocampalAmygdala
DefaultC
SomMotB
LimbicA
SomMotA
TempPar
SalVentAttnB
ContA
SalVentAttnA
Thalamus
DefaultB
DefaultA
ContB
ContC
0.6
0.4
0.2
0
−0.2
−0.4
−0.6

#### fMRI PC12

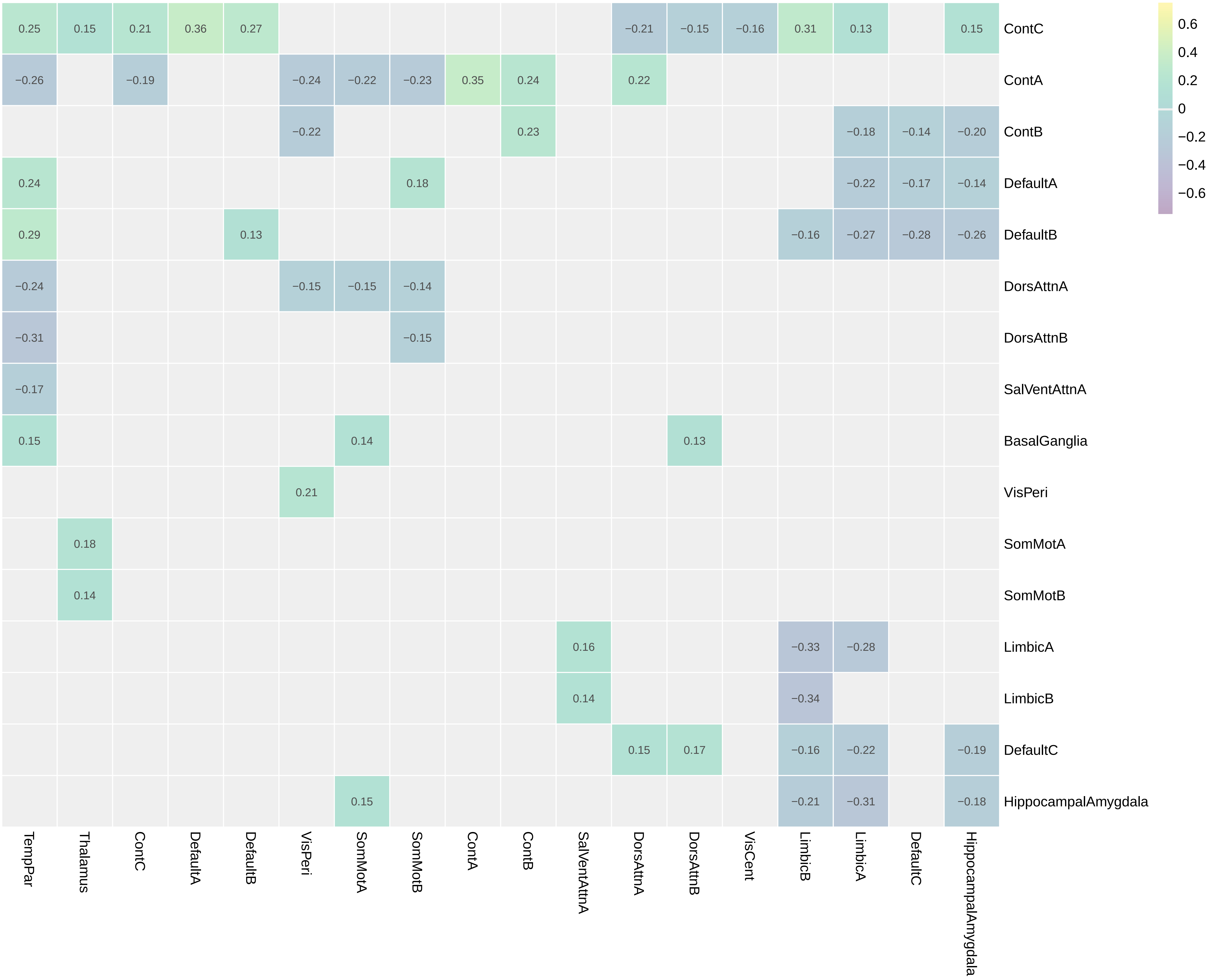
ContC
ContA
ContB
DefaultA
DefaultB
DorsAttnA
DorsAttnB
SalVentAttnA
BasalGanglia
VisPeri
SomMotA
SomMotB
LimbicA
LimbicB
DefaultC
HippocampalAmygdala
TempPar
Thalamus
ContC
DefaultA
DefaultB
VisPeri
SomMotA
SomMotB
ContA
ContB
SalVentAttnA
DorsAttnA
DorsAttnB
VisCent
LimbicB
LimbicA
DefaultC
HippocampalAmygdala
0.6
0.4
0.2
0
−0.2
−0.4
−0.6

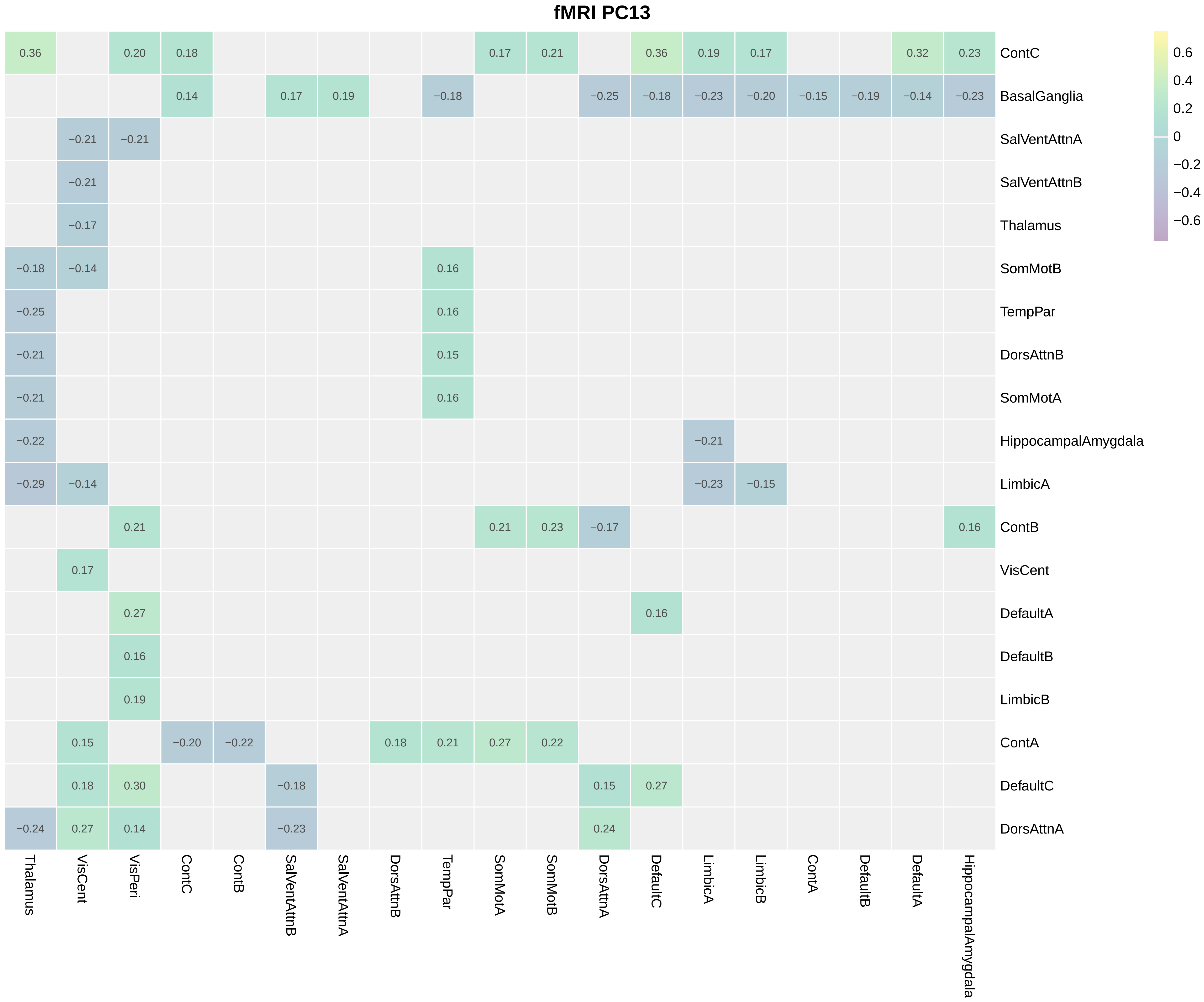
fMRI PC13
0.6
0.4
0.2
0
−0.2
−0.4
−0.6
0.36 0.20 0.18 0.17 0.21 0.36 0.19 0.17 0.32 0.23 ContC
0.14 0.17 0.19 −0.18 −0.25 −0.18 −0.23 −0.20 −0.15 −0.19 −0.14 −0.23 BasalGanglia
−0.21 −0.21 SalVentAttnA
−0.21 SalVentAttnB
−0.17 Thalamus
−0.18 −0.14 0.16 SomMotB
−0.25 0.16 TempPar
−0.21 0.15 DorsAttnB
−0.21 0.16 SomMotA
−0.22 −0.21 HippocampalAmygdala
−0.29 −0.14 −0.23 −0.15 LimbicA
0.21 0.21 0.23 −0.17 0.16 ContB
0.17 VisCent
0.27 0.16 DefaultA
0.16 DefaultB
0.19 LimbicB
0.15 −0.20 −0.22 0.18 0.21 0.27 0.22 ContA
0.18 0.30 −0.18 0.15 0.27 DefaultC
−0.24 0.27 0.14 −0.23 0.24 DorsAttnA
Thalamus
VisCent
VisPeri
ContC
ContB
SalVentAttnB
SalVentAttnA
DorsAttnB
TempPar
SomMotA
SomMotB
DorsAttnA
DefaultC
LimbicA
LimbicB
ContA
DefaultB
DefaultA
HippocampalAmygdala

#### fMRI PC14

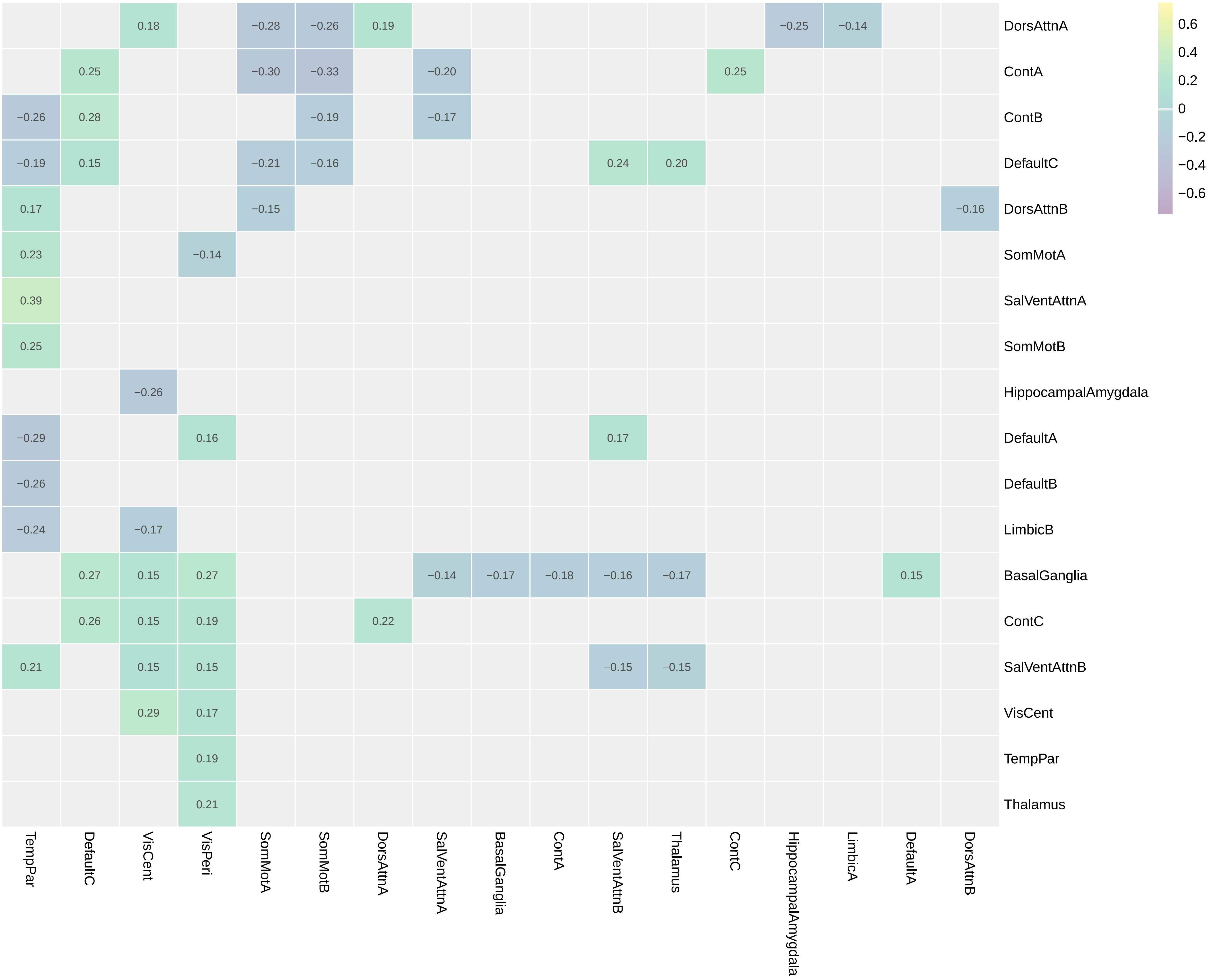
0.18 −0.28 −0.26 0.19 −0.25 −0.14 DorsAttnA
0.25 −0.30 −0.33 −0.20 0.25 ContA
−0.26 0.28 −0.19 −0.17 ContB
−0.19 0.15 −0.21 −0.16 0.24 0.20 DefaultC
0.17 −0.15 −0.16 DorsAttnB
0.23 −0.14 SomMotA
0.39 SalVentAttnA
0.25 SomMotB
−0.26 HippocampalAmygdala
−0.29 0.16 0.17 DefaultA
−0.26 DefaultB
−0.24 −0.17 LimbicB
0.27 0.15 0.27 −0.14 −0.17 −0.18 −0.16 −0.17 0.15 BasalGanglia
0.26 0.15 0.19 0.22 ContC
0.21 0.15 0.15 −0.15 −0.15 SalVentAttnB
0.29 0.17 VisCent
0.19 TempPar
0.21 Thalamus
TempPar
DefaultC
VisCent
VisPeri
SomMotA
SomMotB
DorsAttnA
SalVentAttnA
BasalGanglia
ContA
SalVentAttnB
Thalamus
ContC
HippocampalAmygdala
LimbicA
DefaultA
DorsAttnB
0.6
0.4
0.2
0
−0.2
−0.4
−0.6

#### fMRI PC15

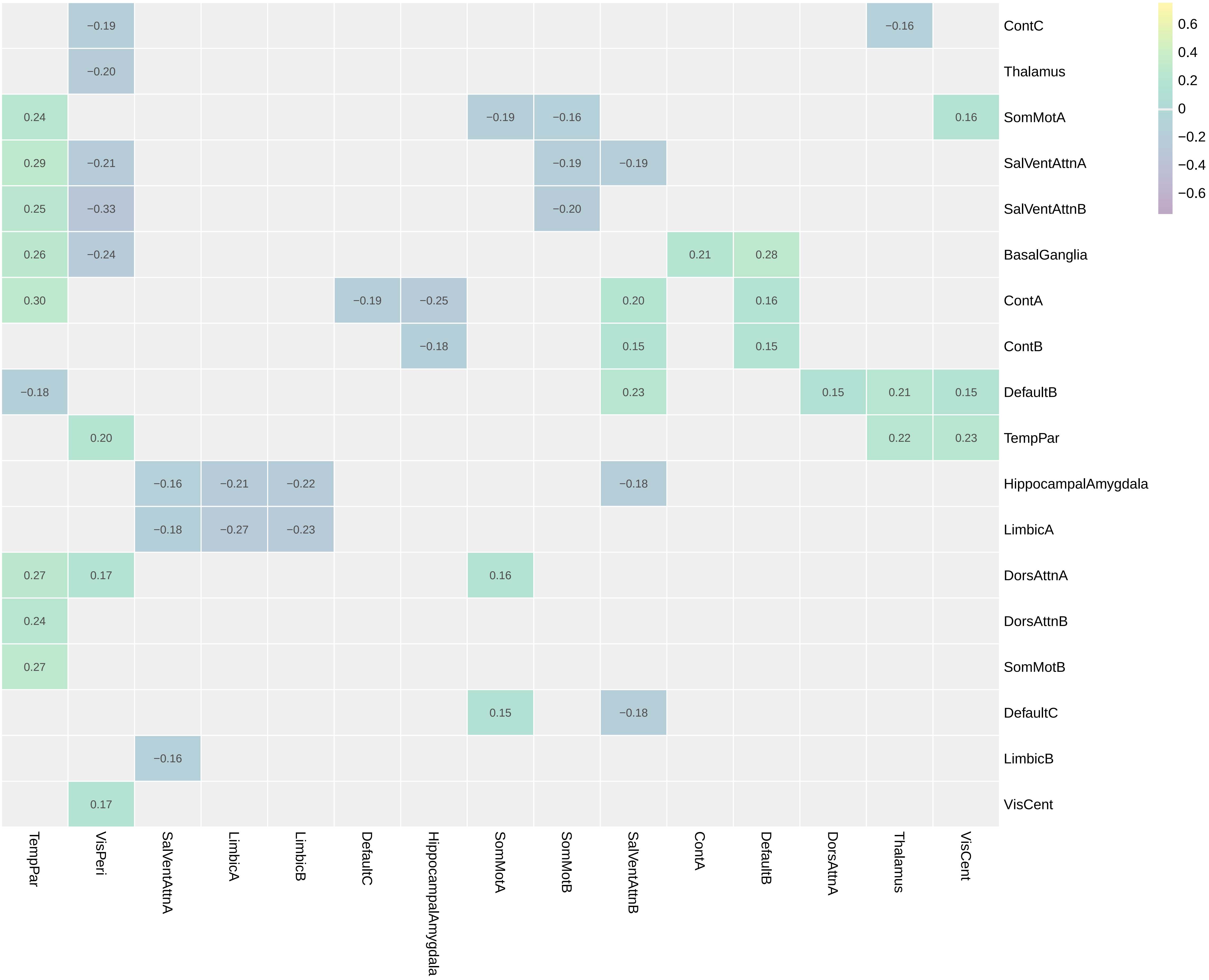

ContC
Thalamus
SomMotA
SalVentAttnA
SalVentAttnB
BasalGanglia
ContA
ContB
DefaultB
TempPar
HippocampalAmygdala
LimbicA
DorsAttnA
DorsAttnB
SomMotB
DefaultC
LimbicB
VisCent
TempPar
VisPeri
SalVentAttnA
LimbicA
LimbicB
DefaultC
HippocampalAmygdala
SomMotA
SomMotB
SalVentAttnB
ContA
DefaultB
DorsAttnA
Thalamus
VisCent
−0.19
−0.16
−0.20
0.24
−0.19
−0.16
0.16
0.29
−0.21
−0.19
−0.19
0.25
−0.33
−0.20
0.26
−0.24
0.21
0.28
0.30
−0.19
−0.25
0.20
0.16
−0.18
0.15
0.15
−0.18
0.23
0.15
0.21
0.15
0.20
0.22
0.23
−0.16
−0.21
−0.22
−0.18
−0.18
−0.27
−0.23
0.27
0.17
0.16
0.24
0.27
0.15
−0.18
−0.16
0.17
0.6
0.4
0.2
0
−0.2
−0.4
−0.6

### fMRI PC16

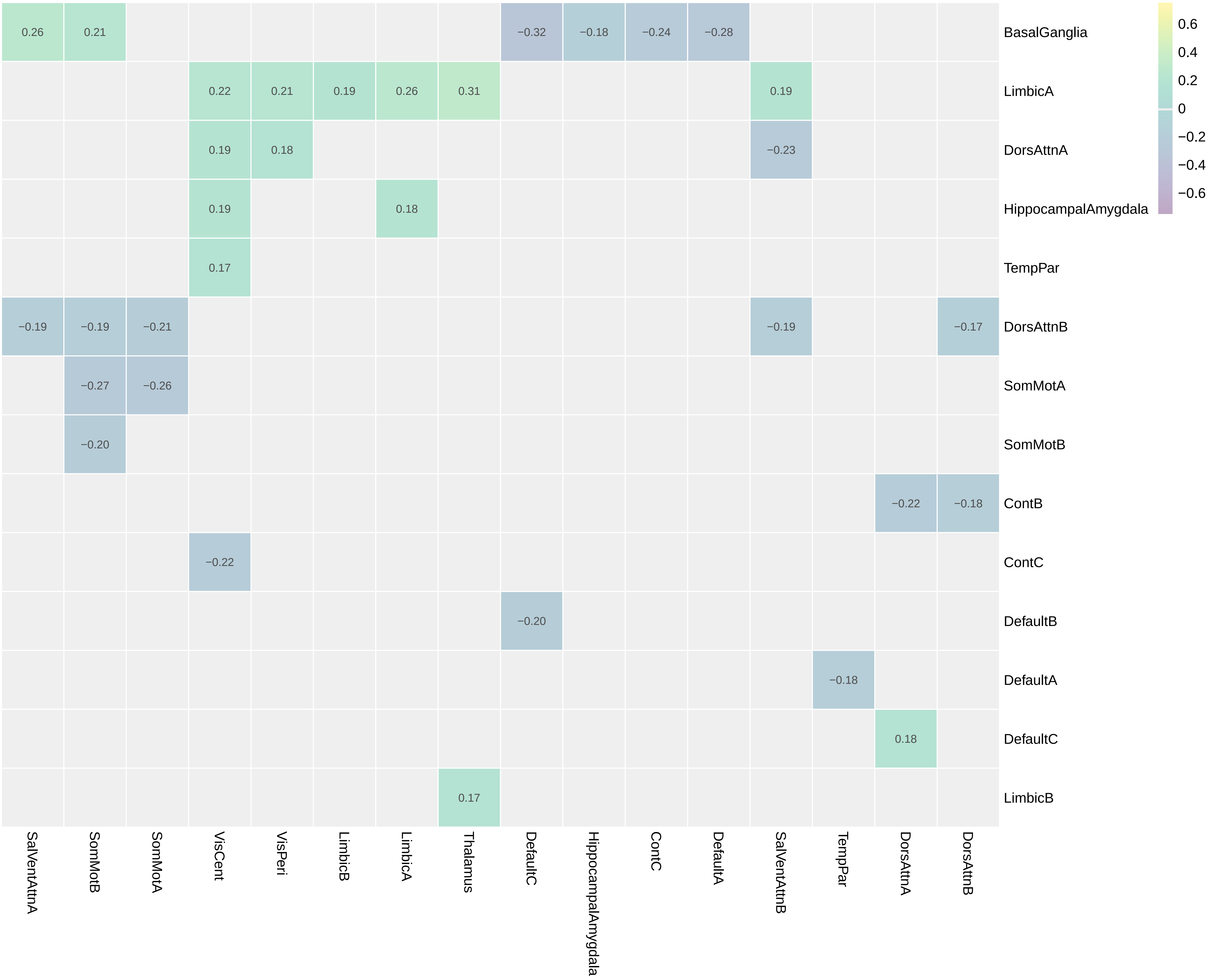

BasalGanglia
LimbicA
DorsAttnA
HippocampalAmygdala
TempPar
DorsAttnB
SomMotA
SomMotB
ContB
ContC
DefaultB
DefaultA
DefaultC
LimbicB
SalVentAttnA
SomMotB
SomMotA
VisCent
VisPeri
LimbicB
LimbicA
Thalamus
DefaultC
HippocampalAmygdala
ContC
DefaultA
SalVentAttnB
TempPar
DorsAttnA
DorsAttnB
0.26
0.21
−0.32
−0.18
−0.24
−0.28
0.22
0.21
0.19
0.26
0.31
0.19
0.19
0.18
−0.23
0.19
0.18
0.17
−0.19
−0.19
−0.21
−0.19
−0.17
−0.27
−0.26
−0.20
−0.22
−0.18
−0.22
−0.20
−0.18
0.18
0.17
0.6
0.4
0.2
0
−0.2
−0.4
−0.6

### fMRI PC17

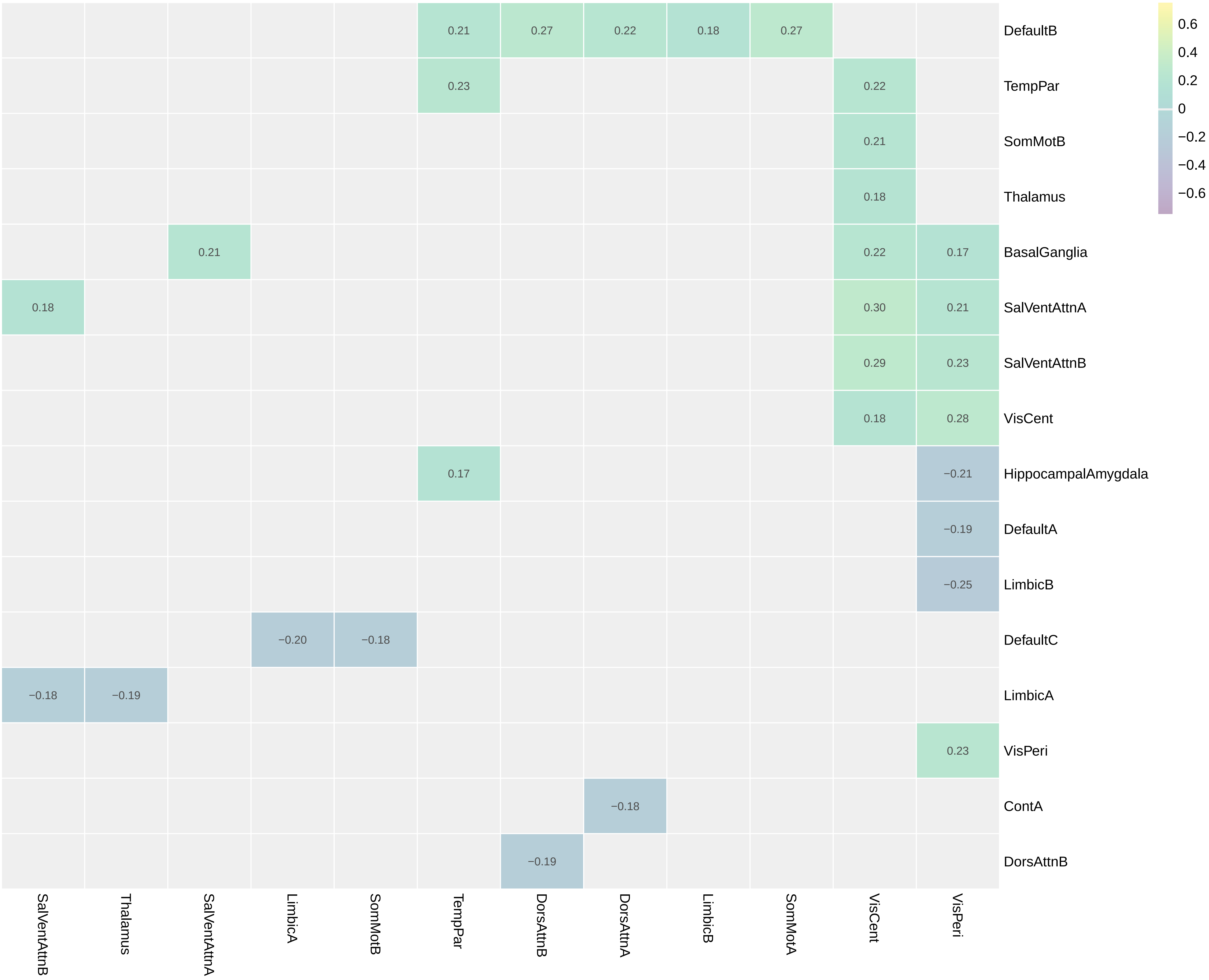

DefaultB
TempPar
SomMotB
Thalamus
BasalGanglia
SalVentAttnA
SalVentAttnB
VisCent
HippocampalAmygdala
DefaultA
LimbicB
DefaultC
LimbicA
VisPeri
ContA
DorsAttnB
SalVentAttnB
Thalamus
SalVentAttnA
LimbicA
SomMotB
TempPar
DorsAttnB
DorsAttnA
LimbicB
SomMotA
VisCent
VisPeri
0.21
0.27
0.22
0.18
0.27
0.23
0.22
0.21
0.18
0.21
0.22
0.17
0.18
0.30
0.21
0.29
0.23
0.18
0.28
0.17
−0.21
−0.19
−0.25
−0.20
−0.18
−0.18
−0.19
0.23
−0.18
−0.19
0.6
0.4
0.2
0
−0.2
−0.4
−0.6

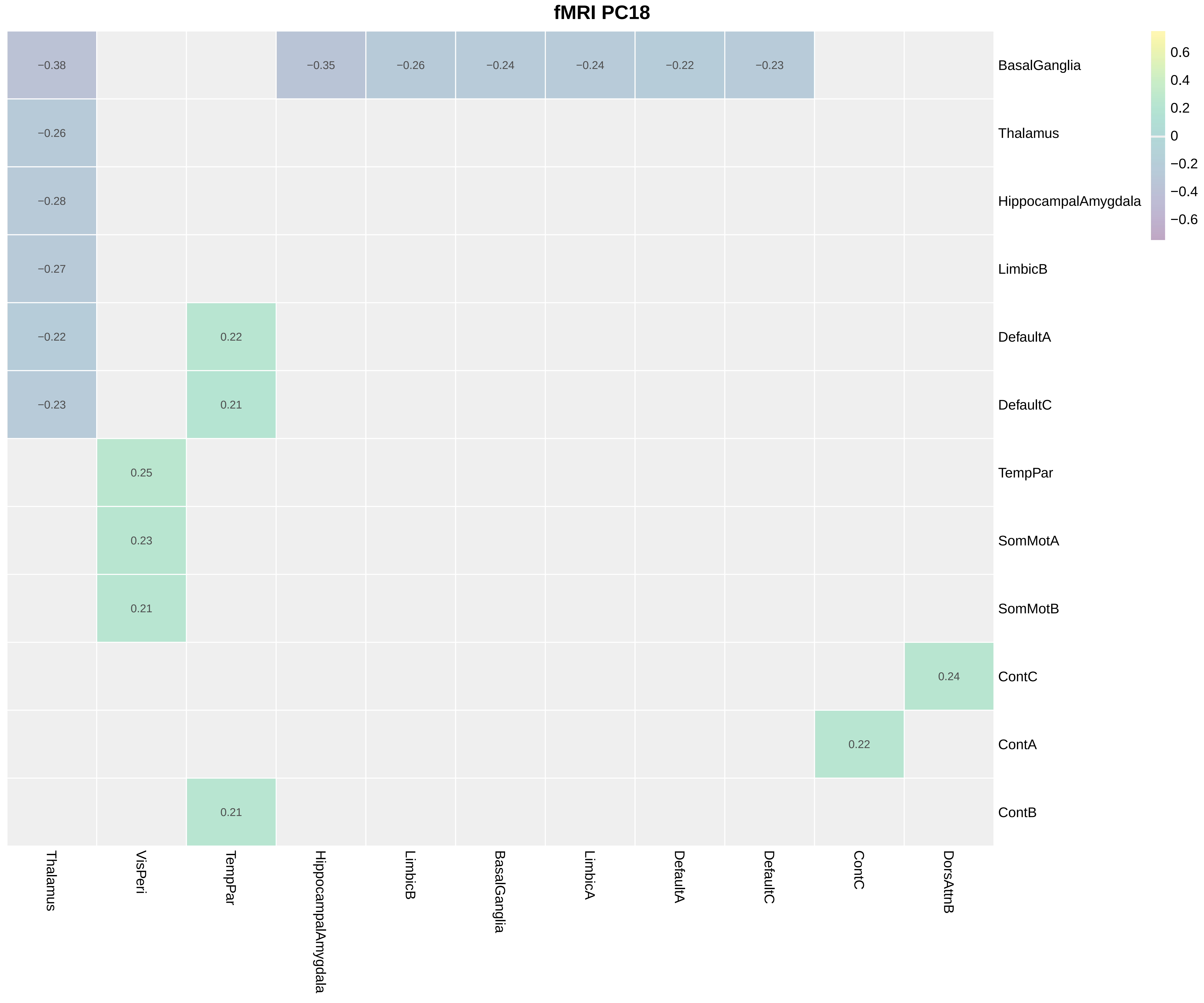
fMRI PC18
BasalGanglia
Thalamus
HippocampalAmygdala
LimbicB
DefaultA
DefaultC
TempPar
SomMotA
SomMotB
ContC
ContA
ContB
Thalamus
VisPeri
TempPar
HippocampalAmygdala
LimbicB
BasalGanglia
LimbicA
DefaultA
DefaultC
ContC
DorsAttnB
−0.38
−0.35
−0.26
−0.24
−0.24
−0.22
−0.23
−0.26
−0.28
−0.27
−0.22
0.22
−0.23
0.21
0.25
0.23
0.21
0.24
0.22
0.21
0.6
0.4
0.2
0
−0.2
−0.4
−0.6

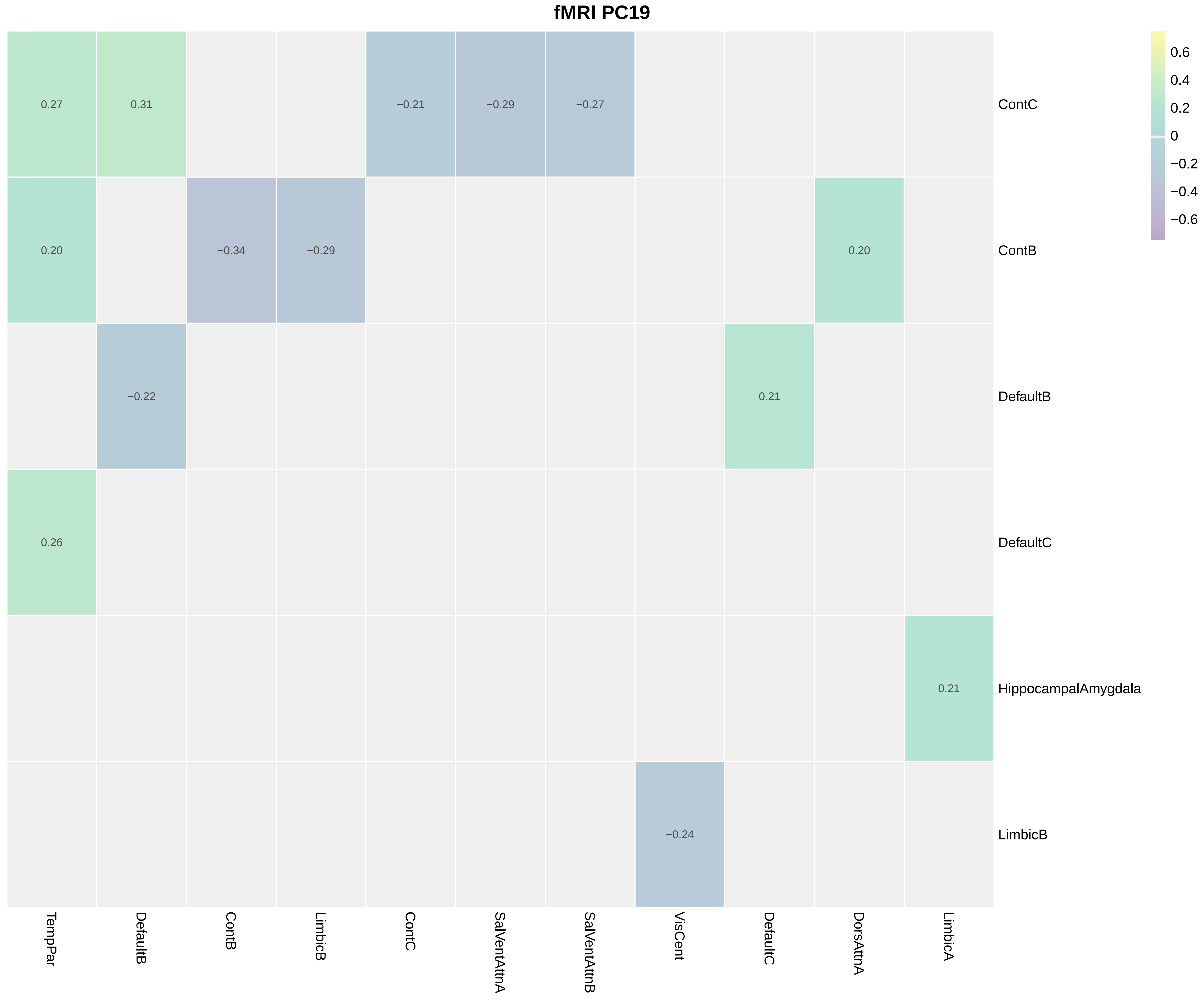
fMRI PC19
0.27
0.31
−0.21
−0.29
−0.27
ContC
0.20
−0.34
−0.29
0.20
ContB
−0.22
0.21
DefaultB
0.26
DefaultC
0.21
HippocampalAmygdala
−0.24
LimbicB
TempPar
DefaultB
ContB
LimbicB
ContC
SalVentAttnA
SalVentAttnB
VisCent
DefaultC
DorsAttnA
LimbicA
0.6
0.4
0.2
0
−0.2
−0.4
−0.6

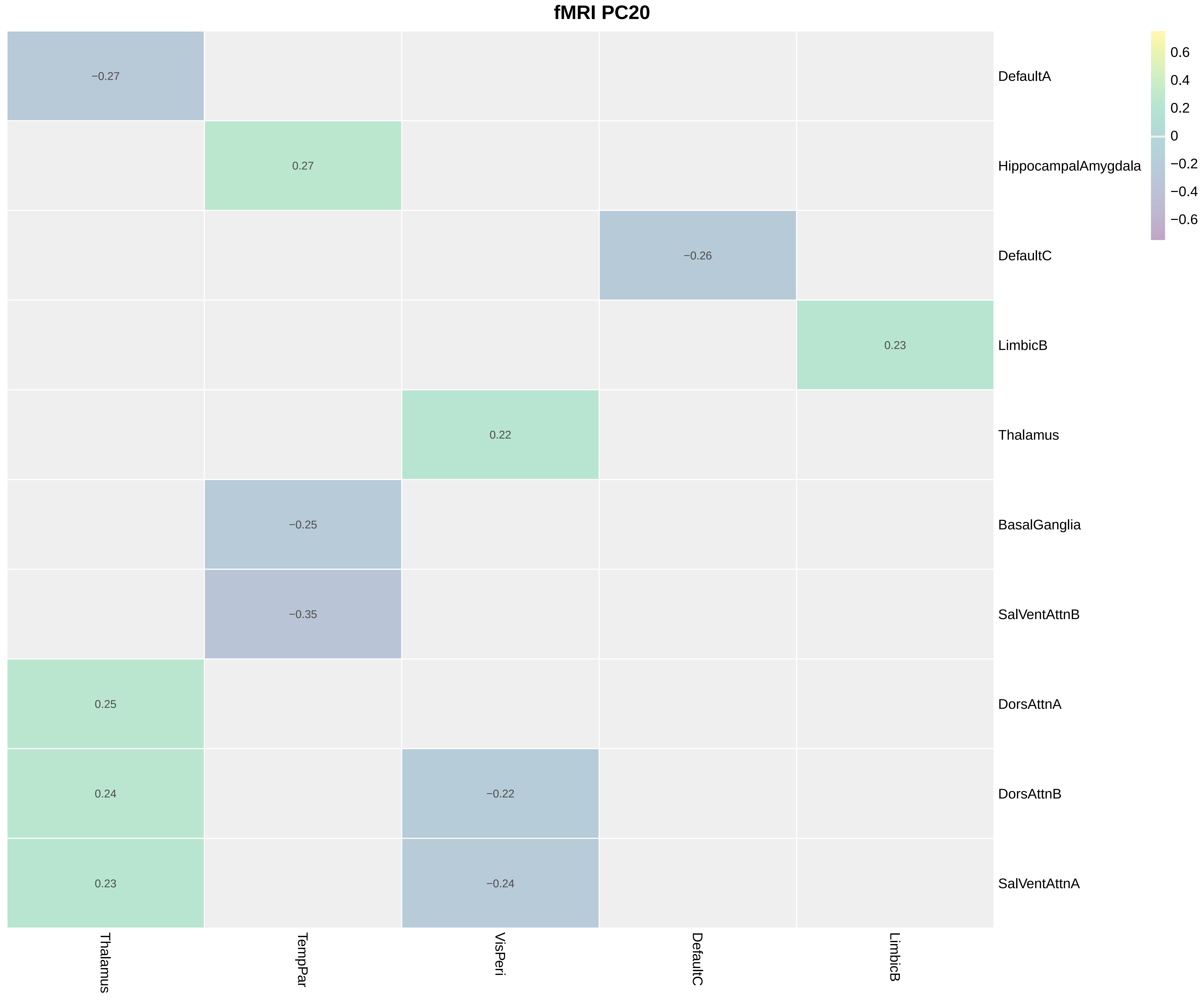
fMRI PC20
−0.27
0.27
−0.26
0.23
0.22
−0.25
−0.35
0.25
0.24
−0.22
0.23
−0.24
DefaultA
HippocampalAmygdala
DefaultC
LimbicB
Thalamus
BasalGanglia
SalVentAttnB
DorsAttnA
DorsAttnB
SalVentAttnA
Thalamus
TempPar
VisPeri
DefaultC
LimbicB
0.6
0.4
0.2
0
−0.2
−0.4
−0.6

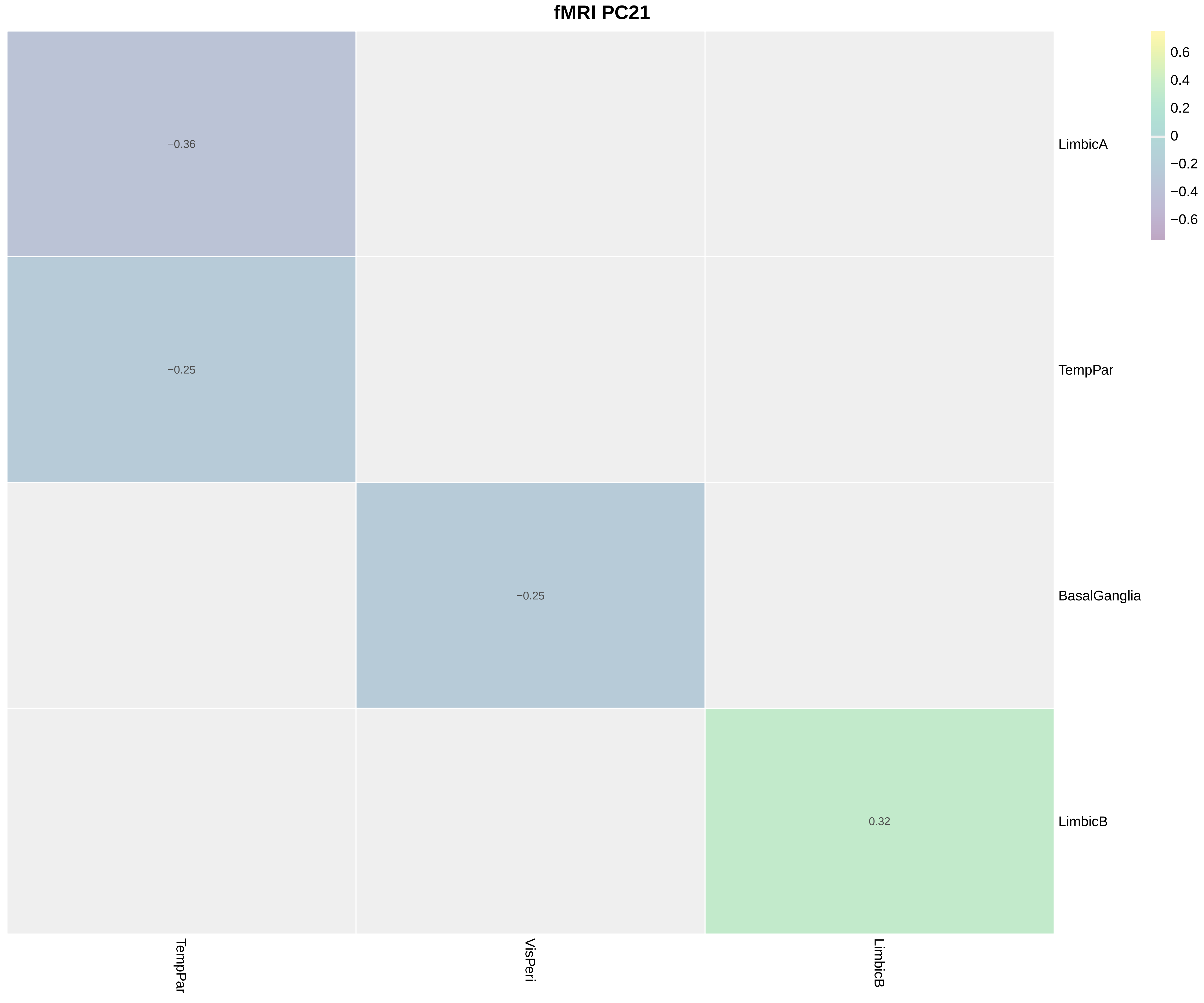
fMRI PC21
−0.36
−0.25
−0.25
0.32
LimbicA
TempPar
BasalGanglia
LimbicB
TempPar
VisPeri
LimbicB
0.6
0.4
0.2
0
−0.2
−0.4
−0.6

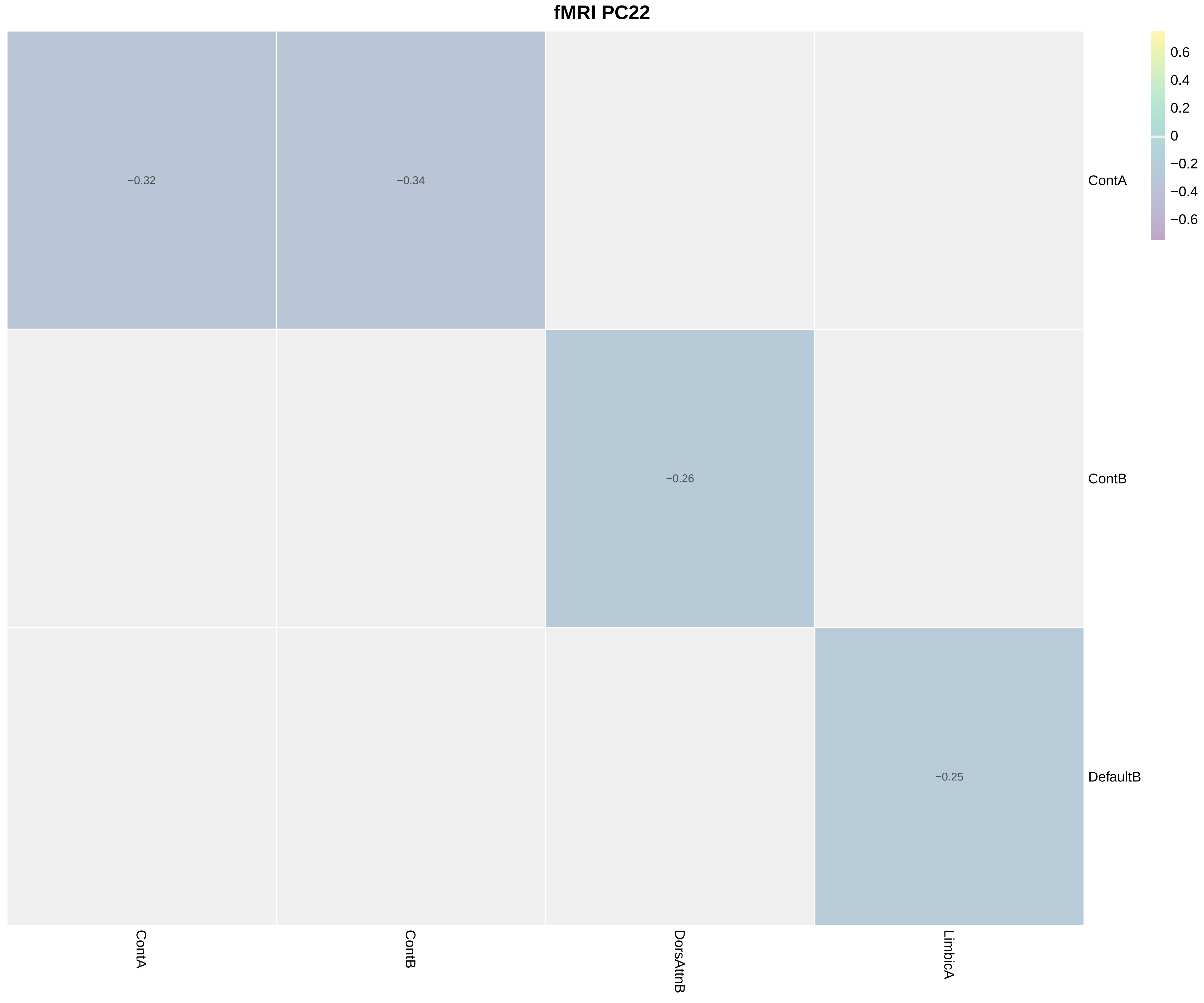
fMRI PC22
−0.32
−0.34
−0.26
−0.25
ContA
ContB
DefaultB
ContA
ContB
DorsAttnB
LimbicA
0.6
0.4
0.2
0
−0.2
−0.4
−0.6
